# Sleep Duration, Quality, and Risk of Peripheral Artery Disease: Multinational Cohort and Mendelian Randomization Studies

**DOI:** 10.1101/2022.06.01.22275864

**Authors:** Shuai Yuan, Michael G. Levin, Olga E Titova, Jie Chen, Yuhao Sun, VA Million Veteran Program, Agneta Åkesson, Xue Li, Scott M. Damrauer, Susanna C. Larsson

## Abstract

**Background:** Sleep duration and quality has been associated with cardiovascular disease, however the effect of sleep on peripheral artery disease (PAD) specifically remains unestablished. We conducted cohort, case-control, and Mendelian randomization (MR) analyses to assess the associations of sleep duration and quality with PAD risk.

**Methods:** Sleep traits were assessed for associations with incident PAD using cohort analysis among 53,416 Swedish adults. Replicated was sought in a case-control study of 28,123 PAD cases and 128,459 controls from the VA Million Veteran Program (MVP) and a cohort study of 452,028 individuals from the UK Biobank study (UKB). Two-sample Mendelian randomization (MR) was used for casual inference-based analyses of sleep-related traits and PAD (31,307 PAD cases 211,753 controls).

**Results:** Observational analyses demonstrated a U-shaped association between sleep duration and PAD risk. In Swedish adults, incident PAD risk was higher in those with short sleep (<5 hours; hazard ratio (HR), 1.74; 95% confidence interval (CI) 1.31-2.31) or long sleep (≥8 hours; HR 1.24; 95% CI 1.08-1.43), compared to individuals with a sleep duration of 7 to <8 hours/night. This finding was supported by case-control analysis in MVP and cohort analysis in UKB. Observational analysis also revealed positive associations between poor sleep quality (HR, 1.81; 95% CI 1.13-2.90) and daytime napping (HR, 1.32; 95% CI 1.18-1.49) with PAD. MR analysis supported an inverse association between sleep duration (odds ratio per hour increase, 0.79, 95% CI, 0.55, 0.89) and PAD, and an association between short sleep and increased PAD (odds ratio, 1.20, 95% CI, 1.04-1.38). MR also found an association between insomnia with PAD (OR, 1.10; 95% CI 1,05-1.15) and a reverse association of PAD on shorter sleep (OR, 1.05; 95% CI 1.01-1.10).

**Conclusions:** Maintenance of healthy sleep habits, especially avoiding habitual short sleep, may prevent PAD.

## 1. Introduction

Sleep plays an important role in health and wellbeing.^1^ Sleep disorders may lead to impaired cardiometabolic profile, activated sympathetic nervous system, systemic inflammation, and accelerated atherosclerosis.^2-6^ Disorders which disrupt consolidated sleep, such as short sleep and insomnia, have been associated with cardiovascular disease in general and coronary artery disease in particular.^7-14^ Apart from inadequate consolidated sleep, sleep-related disorders like sleep apnea, as well as daytime napping (usually regarded as a healthy habit), have also been associated with increased risk of cardiovascular events.^11,15^

Peripheral artery disease (PAD) is the third leading cause of atherosclerotic cardiovascular morbidity, and its prevalence has dramatically increased over recent decades.^16,17^ Although attempts have been made to link sleep-related traits to the development of PAD, the results have been inconsistent^18 19^. Further, while poor sleep quality is a common complaint among individuals suffering from PAD,^20^ prior observational associations between sleep duration and PAD risk are conflicting.^18^ These observational studies may have been limited by small sample size, residual confounding, and reverse causation. Because of these factors, the relationship between sleep quality, sleep duration, and PAD has remained poorly defined.

MR is an analytic framework that utilizes genetic variants as instrumental variables to limit bias from residual confounding and reverse causation. Because genetic variants are randomly allocated at conception, under certain assumptions the MR framework mimics randomized controlled trials and enables more robust causal inference than traditional epidemiologic techniques.^21^ Here, we conducted observational and MR analyses to test the hypotheses that disordered (short or long) sleep duration, poor sleep quality, and daytime napping increase PAD risk. A clear appraisal of these associations can help inform risk mitigation strategies for PAD.

## 2. Methods

### 2.1 Study design

Figure 1 shows the overall design of the present study. We first examined the associations of 3 sleep-related traits (sleep duration, quality, and daytime napping) with incident PAD in a cohort analysis using data from the Swedish Infrastructure for Medical Population-based Life-course and Environmental Research (SIMPLER). The association between sleep duration and PAD was also examined in a case-control analysis in the VA Million Veteran Program (MVP) and in a cohort analysis in the UK Biobank (UKB). Likewise, the association for daytime napping was examined in UKB. Second, we calculated genetic correlations between the sleep-related traits and PAD using genome-wide association data. We then performed nonlinear and two-sample MR analyses to estimate the effects of sleep-related traits on PAD using data from MVP and UKB. The study has been approved by the Swedish Ethical Review Authority (no. 2019-03986) and the United States VA Central IRB. The UK Biobank received ethical permits from the North West Multi-centre Research Ethics Committee, the National Information Governance Board for Health and Social Care in England and Wales, and the Community Health Index Advisory Group in Scotland. All participants have provided informed consent.

**Figure 1.**
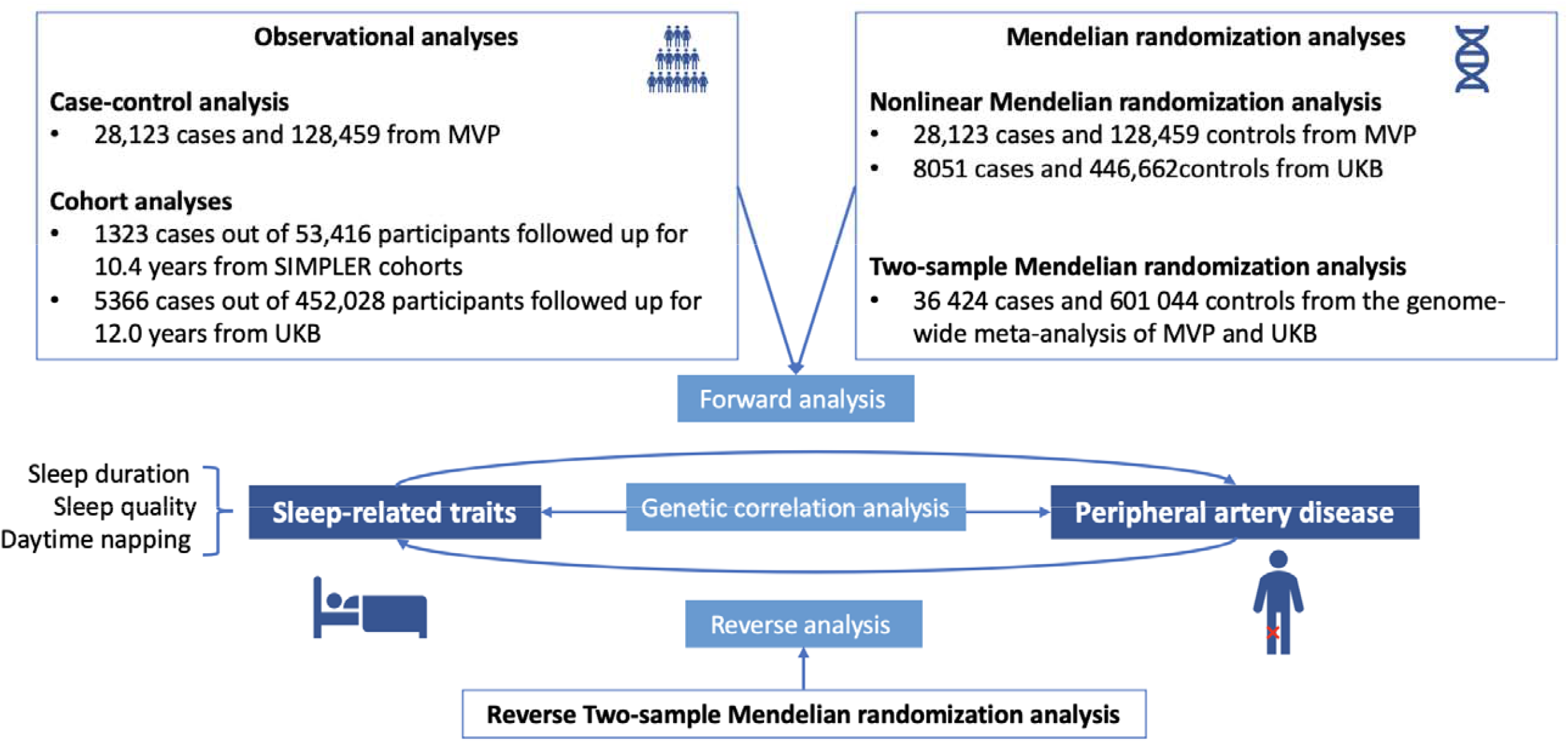
Study design overview. MVP, the VA Million Veteran Program; UKB, UK Biobank.

### 2.2 Prospective cohort analysis in SIMPLER

The cohort analysis used data from SIMPLER, which includes the Swedish Mammography Cohort and the Cohort of Swedish Men. Detailed description of the two cohorts is provided in **Supplementary Methods**. Information on sleep duration and quality, and daytime napping was obtained from participants in the 2008-2009 survey, which included 54,859 middle-aged and older Swedish adults. After removal of individuals who died before 1 July 2009 and those with baseline PAD, 53,416 participants were included in analysis for sleep quality and daytime napping. For sleep duration, data from 49,065 individuals were used after further exclusion of 4351 participants who reported a sleep duration of <2 or >13 hours per night (**Supplementary Figure 1**). Information on sleep duration (in hours/night, open question), 6 sleep measures of disrupted sleep quality, and daytime napping (yes/no) were extracted from the 2008 self-administrated questionnaire. Sleep quality was assessed by a sleep quality score based on six sleep measures and a higher score represents more sleep complaints (**Supplementary Methods**). Covariates included age, sex, education level, smoking, physical activity, and diet quality (**Supplementary Methods**).^22^ We also obtained data on baseline body mass index as well as history of hypertension, hypercholesterolemia, and diabetes, which were regarded as potential mediators. Incident PAD cases were ascertained by clinical diagnosis based on codes from the 9^th^ and 10^th^ revision of the International Classification of Diseases (ICD). Cases were identified by linkage of the cohorts to the Swedish Patient Register, which has a nearly complete coverage of hospital-based inpatient and outpatient care.^23^ Death information was obtained from the Swedish Death Registry. Individuals were followed up from July 1, 2009, until the date of diagnosis of PAD, date of death, or end of follow-up (i.e., 31 December 2019), whichever came first.

### 2.3 Case-control and nonlinear MR analyses in MVP

MVP is a longitudinal precision medicine initiative which recruits subjects who receive care from the nationwide Veterans Affairs healthcare system. Details of MVP have been previously described.^24^ We identified participants of MVP with available genotype, electronic health record, and lifestyle survey data, and included 28,123 PAD cases and 128,459 controls in the analysis. Self-reported sleep duration was extracted from the MVP Lifestyle Survey and was coded as a factor (5 or less, 6, 7, 8, 9, 10 or greater hours) to enable evaluation for a non-linear association between sleep and PAD. Sleep duration of 7 hours was set as the reference level. PAD status was assigned using electronic health record diagnosis codes, as previously described.^25^ Covariates included age, sex, 5 ancestry-specific genetic principal components, self-reported education level, self-reported physical activity, body mass index, hypertension, hemoglobin A1c, smoking history, and statin use at baseline.

### 2.4 Prospective cohort and nonlinear MR analyses in UKB

UKB was initiated by recruiting more than 500,000 participants from 22 assessment centers across the United Kingdom between 2006 and 2010. In this study, we removed individuals with extreme sleep duration per day (≤1% and ≥99% of sleep duration), with non-European descent (to minimize population structure bias), without genetic information, with baseline PAD diagnosis, and with incomplete information on sleep duration and daytime napping. A total of 452,028 individuals was included in the analysis. Information on sleep duration and daytime napping was obtained from the touchscreen questionnaire at baseline. Daytime napping was defined as two groups, yes (sometimes and usually) and no (never/rarely). PAD cases were defined by a primary or secondary diagnosis with data on admissions and diagnoses and corresponding dates from hospital inpatient records using the ICD-9 and ICD-10. Covariates included age, sex, education level, smoking status, physical activity, diet quality, body mass index, and baseline history of hypertension, hypercholesteremia, and diabetes. Death information was obtained from the Death Registry. Individuals were followed up from the baseline (2006-2010), until the date of diagnosis of PAD, date of death, date of loss to follow-up, the last date of hospital admission (i.e., HES and SMR: 31 March 2021, and PEDW: 28 February 2018), whichever came first. Prevalent PAD cases and controls were used in the nonlinear MR analysis.

### 2.5 Two-sample MR analysis

DNA variants with genome-wide significant (p<5x10^−8^) associations with sleep-related traits, including continuous sleep duration, short (<7 hours/night) and long sleep (≥9 hours/night), sleep apnea, snoring, insomnia, and daytime napping, were obtained from the corresponding genome-wide association studies (GWAS) in the public domain. Linkage disequilibrium among these genetic variants for each trait was estimated using 1000 Genomes Phase 3 Version 5 European panel as the reference population.^26^ Independent SNPs (i.e., without linkage disequilibrium, defined by *r*^2^ <0.01 and clump window >10,000 kb) were used as instrumental variables. Detailed information on the genome-wide association studies on sleep-related traits and SNPs included as genetic instruments is presented in **Supplementary Table 1** and **Supplementary Table 2**, respectively.

Summary-level data for PAD corresponding to the sleep-associated variants was obtained from a genome-wide association meta-analysis of MVP and UKB including 31,307 cases and 211,753 controls of European descent (**Supplementary Table 3**).^25^ Cases were defined by having at least two of the ICD-9/10 codes/Current Procedural Terminology codes. For the reverse MR analysis of the association of PAD with sleep traits, we used a genetic instrument for PAD that consisted of 19 independent SNPs associated with PAD at *p*<5×10^−8^ in a multi-ancestry meta-analysis (36,424 PAD cases and 601,044 controls) of genome-wide analyses of MVP (the discovery stage) and UKB (the replication stage).^25^ To minimize bias due to sample overlap,^27^ a complementary set of genetic instruments (n = 18) for PAD was selected based on data from only MVP as the instruments for sleep-related traits were obtained from UK Biobank (**Supplementary Table 4**).

### 2.6 Statistical analysis

For the cohort analyses, we used the Cox proportional hazards regression model to estimate the associations of sleep-related traits with risk of incident PAD with age as the underlying time scale. The assumption of proportionality was satisfied by checking Schoenfeld residuals. The nonlinear association between sleep duration and incident PAD was analyzed using a Cox proportional hazards model where sleep duration was entered in the model as a restricted cubic spline with three knots placed at the 25^th^ (5 hours), 50^th^ (7 hours) and 75^th^ (8 hours) percentiles of sleep duration and 6.9 hours for MVP) and 7.0 hours for UKB was used as a reference. We employed three statistical models: model 1 included age and sex as covariates; model 2 included age, sex, education level, smoking status, physical activity, and diet quality; and model 3 included the aforementioned covariates plus potential mediators, including baseline body mass index and history of hypertension, hypercholesterolemia, and diabetes.

For the case-control analyses, the association between self-reported sleep duration and prevalent peripheral artery disease was assessed using logistic regression. Like the cohort analyses, we used three models: model 1 was adjusted for age, sex, and ancestry-specific genetic principal components; model 2 additionally included education level, self-reported physical activity, and smoking; model 3 further included hemoglobin A1c, body mass index, history of hypertension, and statin use. Analyses were performed separately by ancestry (European or African ancestry) as determined by HARE.^28^ Results were combined using fixed-effects meta-analysis.

Genetic correlation was calculated using LDSC software.^29,30^ For two-sample MR analyses, the inverse-variance weighted method with multiplicative random-effects was used as the primary statistical method and supplemented with three sensitivity analyses, including the weighted median,^31^ MR-Egger^32^ and MR-PRESSO methods, which make different assumptions about the presence of outliers and pleiotropy.^33^ Cochran’s Q value was used to assess the heterogeneity among estimates from SNPs. For nonlinear MR analysis, a weighted polygenic score was constructed based on genetic instruments for sleep duration using plink2. Nonlinear MR analysis was the performed as described by Staley and Burgess, implemented using the nlmr package in R.^34^ Briefly, individuals were stratified by residual sleep duration after conditioning on the effects of the genetic instrument. Then, localized average causal effects within each stratum of sleep duration were estimated using the ratio of coefficients MR method. Finally, nonlinearity of the localized average causal effects was assessed using the fractional polynomial method. Nonlinear MR was performed among the European-ancestry subset of MVP, as the limited number of cases among African-ancestry participants within each stratum of sleep duration was expected to be insufficiently powered to detect differences in localized average causal effects. All statistical tests were two-sided, and the analyses were performed in Stata/SE (version 15.0; StataCorp, Texas, USA) and R software (version 4.0.2). A *p* value below 0.05 was regarded as statistically significant.

## 3. Results

Figure 1 shows the overall study design. We first present the results of observational analyses investigating associations between sleep and PAD in SIMPLER, MVP, and UKB. Then, we present the results of MR analyses which use genetic instruments to estimate causal associations between sleep traits and PAD.

### 3.1 Cohort analysis in SIMPLER

We first examined the association between sleep duration and incident PAD among participants of the SIMPLER cohort. Age-standardized baseline characteristics of SIMPLER participants by sleep duration are presented in **Table 1**. A total of 1,323 incident PAD cases (1,202 cases in the analysis of sleep duration) were identified among cohort participants after a median of 10.4 years and a combined 488,385 person-years of follow-up. A U-shaped association between sleep duration and incident PAD was observed (*p* for nonlinearity <0.001, **Figure 2 A**). Both short and long sleep were associated with an increased PAD risk (**Table 2**). In an analysis adjusted for age, sex, education level, smoking status, physical activity, and diet quality, compared with individuals with sleep duration of 7 to <8 hours per night, the hazard ratios (HR) for incident PAD were 1.74 (95% confidence interval (CI), 1.31, 2.31) for those with <5 hours of sleep and 1.24 (95% CI, 1.08, 1.43) for those with ≥8 hours of sleep (**Table 2**). The associations attenuated slightly in the model 3 with further adjustment for possible mediators (**Table 2**).

**Table 1.** Age-standardized baseline characteristics of 49 065 Swedish adults by sleep duration in SIMPLER.

| No. of individual and characteristics | Sleep duration in hours per night |  |  |  | Total |
| --- | --- | --- | --- | --- | --- |
|  | <5 | ≥5 & <7 | ≥7 & <8 | ≥8 |  |
| No. of Individuals | 1460 | 14 743 | 16 954 | 15 908 | 49 065 |
| Age, mean±SD, years | 72.7±8.7 | 71.3±8.5 | 69.9±8.0 | 71.7±8.0 | 71.0±8.2 |
| Sleep duration, median±IQR, hours | 4±2 | 6±1.5 | 7±0.5 | 8±4 | 7±10 |
| Male, % | 37.1 | 53.2 | 60.2 | 55.1 | 55.7 |
| Body mass index, mean±SD, kg/m<sup>2</sup> | 26.0±4.4 | 25.6±3.6 | 25.3±3.4 | 25.3±3.5 | 25.4±3.5 |
| Post-secondary education, % | 16.3 | 18.7 | 22.9 | 21.5 | 21.1 |
| Hypertension, % | 49.6 | 44.0 | 39.5 | 41.1 | 41.6 |
| Hypercholesterolemia, % | 26.4 | 25.4 | 24.0 | 25.0 | 24.8 |
| Diabetes, % | 14.5 | 9.4 | 7.9 | 9.4 | 9.0 |
| Smoking status, % |  |  |  |  |  |
| Never smoker | 38.9 | 41.9 | 44.0 | 41.4 | 42.4 |
| Past smoker | 29.4 | 32.2 | 32.9 | 32.5 | 32.5 |
| Current smoker | 9.7 | 8.0 | 7.2 | 8.9 | 8.0 |
| Physical activity, % |  |  |  |  |  |
| <10 mins/day | 7.6 | 5.3 | 4.1 | 5.2 | 4.9 |
| 10-30 mins/day | 14.4 | 12.6 | 11.9 | 12.7 | 12.4 |
| 31-60 mins/day | 38.9 | 44.7 | 46.3 | 44.5 | 45.0 |
| >60 mins/day | 22.8 | 24.3 | 26.1 | 25.0 | 25.1 |
| DASH score, mean±SD | 17.3±3.4 | 17.6±3.4 | 17.8±3.4 | 17.6±3.4 | 17.7±3.4 |
| Daytime napping, % | 24.6 | 23.4 | 21.9 | 25.0 | 23.4 |
| Sleep quality (often, mostly, or always), % |  |  |  |  |  |
| Difficulty falling asleep | 55.6 | 31.9 | 12.5 | 9.7 | 18.6 |
| Repeatedly waking up with difficulty falling asleep | 65.7 | 44.7 | 18.0 | 13.0 | 25.7 |
| Premature awakening | 66.1 | 50.9 | 21.2 | 11.6 | 28.2 |
| Disturbed or restless sleep | 56.1 | 37.1 | 16.4 | 13.3 | 22.7 |
| Sleep apnea | 9.8 | 8.4 | 6.6 | 7.1 | 7.4 |
| Disturbing snoring | 21.3 | 23.3 | 22.4 | 23.0 | 22.8 |

DASH, Dietary Approaches to Stop Hypertension; IQR, interquartile range; SD, standard deviation. <sup>a</sup> Missing information was around 17.1% for smoking status, 12.6% for physical activity, 24.5% for sleep medication, 2.2% for napping, 2.5% for difficulty falling asleep, 4.8% for repeatedly waking up with difficulty falling asleep, 5.0% for premature awakening, 6.0% for disturbed or restless sleep, 6.8 % for sleep apnea and 4.7% for disturbing snoring.

**Figure 2.**
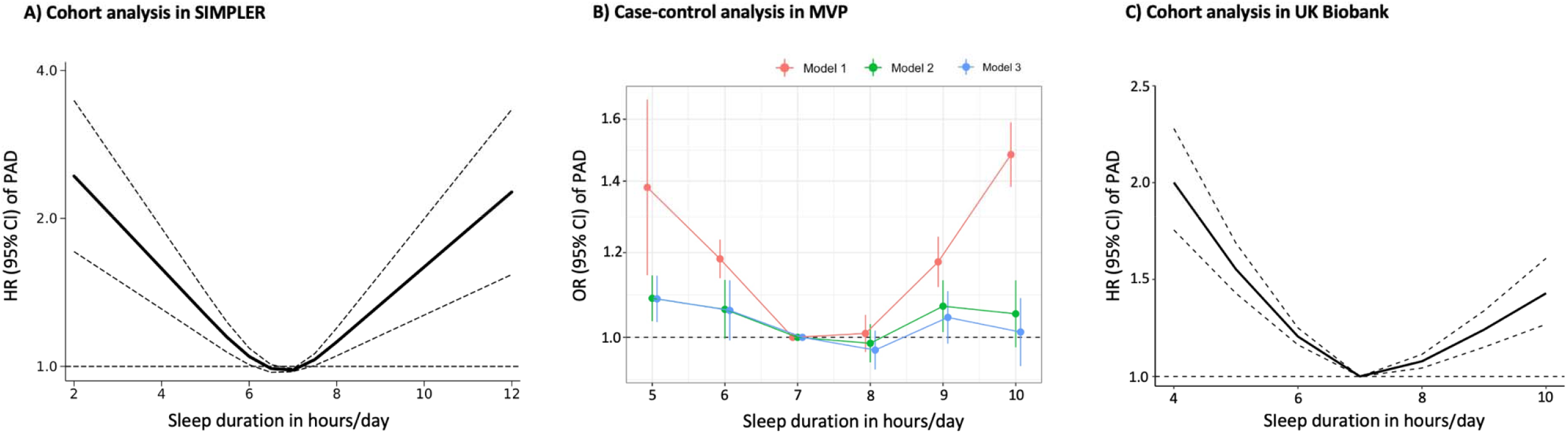
The U-shaped association of sleep duration with risk of peripheral artery disease in cohort and case-control studies. CI, confidence interval; HR, hazard ratio; MVP, the VA Million Veteran Program; PAD, peripheral artery disease; SIMPLER, the Swedish Infrastructure for Medical Population-based Life-course and Environmental Research. The cohort analyses in SIMPLER and UK Biobank were conducted using restricted cubic spline Cox regression and with 6.9 and 7 hours of sleep per day as the reference value, respectively. Estimates were adjusted for age, sex, education level, smoking status, physical activity, and diet quality in SIMPLER. Estimates were adjusted age, sex, education level, smoking status, physical activity, diet quality, body mass index, and baseline history of hypertension, hypercholesteremia, diabetes in UK Biobank. For analysis in MVP, model 1 was adjusted for age, sex, and ancestry-specific genetic principal components; model 2 additionally included education level, self-reported physical activity, and smoking; model 3 further included hemoglobin A1c, body mass index, history of hypertension, and statin use.

**Table 2.** Associations of sleep-related traits with risk of peripheral artery disease in the cohort analysis in SIMPLER.

| Sleep trait | Cases | Total | Person-years | Model 1 |  | Model 2 |  | Model 3 |  |
| --- | --- | --- | --- | --- | --- | --- | --- | --- | --- |
|  |  |  |  | HR (95% CI) | P | HR (95% CI) | P | HR (95% CI) | P |
| Sleep duration (n = 49 065) |  |  |  |  |  |  |  |  |  |
| <5 hours/day | 55 | 1 460 | 12 685 | 1.90 (1.43, 2.53) | <0.001 | 1.74 (1.31, 2.31) | <0.001 | 1.55 (1.16, 2.06) | 0.003 |
| ≥5 & <7 hours/day | 361 | 14 743 | 134 788 | 1.18 (1.02, 1.37) | 0.027 | 1.12 (0.97, 1.30) | 0.124 | 1.09 (0.94, 1.26) | 0.264 |
| ≥7 & <8 hours/day | 344 | 16 954 | 160 072 | 1.00 | Ref | 1.00 | Ref | 1.00 | Ref |
| ≥8 hours/day | 442 | 15 908 | 143 372 | 1.32 (1.15, 1.52) | <0.001 | 1.24 (1.08, 1.43) | 0.003 | 1.22 (1.06, 1.41) | 0.006 |
| Sleep quality score (n = 43 285) |  |  |  |  |  |  |  |  |  |
| 0 | 484 | 19 830 | 182 813 | 1.00 | Ref | 1.00 | Ref | 1.00 | Ref |
| 1 | 198 | 8925 | 83 414 | 0.93 (0.79, 1.10) | 0.422 | 0.89 (0.76, 1.06) | 0.189 | 0.90 (0.76, 1.06) | 0.203 |
| 2 | 115 | 5371 | 50 200 | 0.92 (0.75, 1.13) | 0.446 | 0.86 (0.70, 1.05) | 0.146 | 0.83 (0.68, 1.01) | 0.070 |
| 3 | 96 | 4085 | 38 102 | 1.03 (0.83, 1.28) | 0.789 | 1.01 (0.81, 1.26) | 0.946 | 0.94 (0.75, 1.17) | 0.579 |
| 4 | 101 | 3698 | 34 036 | 1.20 (0.97, 1.49) | 0.099 | 1.14 (0.92, 1.42) | 0.228 | 1.06 (0.85, 1.31) | 0.628 |
| 5 | 23 | 979 | 9084 | 1.09 (0.72, 1.65) | 0.695 | 0.96 (0.63, 1.46) | 0.853 | 0.89 (0.58, 1.35) | 0.570 |
| 6 | 18 | 397 | 3696 | 2.18 (1.36, 3.49) | 0.001 | 1.81 (1.13, 2.90) | 0.014 | 1.54 (0.96, 2.48) | 0.073 |
| Daytime napping (n = 53 416) |  |  |  |  |  |  |  |  |  |
| No | 827 | 39 300 | 370 965 | 1.00 | Ref | 1.00 | Ref | 1.00 | Ref |
| Yes | 449 | 12 394 | 102 787 | 1.43 (1.27, 1.61) | <0.001 | 1.32 (1.18, 1.49) | <0.001 | 1.23 (1.09, 1.39) | 0.001 |

CI, confidence interval; HR, hazard ratio.
The sleep quality score was constructed based on 6 sleep problems, with a higher score representing more problems. Individuals with missing information were classified in a separate missing group in the analysis of daytime napping and were excluded in the analysis of sleep duration and sleep quality.
Model 1 adjusted for age and sex.
Model 2 adjusted for age, sex, education level, smoking status, physical activity, and diet quality.
Model 3 adjusted for the same covariates as in Model 2 and additionally for potential mediators, including body mass index and baseline history of hypertension, hypercholesteremia, and diabetes.

Next, we evaluated the association between self-reported sleep quality and incident PAD. Individuals with 6 sleep quality complaints, compared to those with no sleep quality complaints, had an increased rate of incident PAD (HR 1.81, 95% CI, 1.13, 2.90) after adjusting for confounders (**Table 2**). We did not observe any association of individual sleep quality traits with PAD risk (**Supplementary Table 5**). Compared with individuals without daytime napping, those taking naps were at increased risk of PAD [HR 1.32 (95% CI, 1.18, 1.49)] in the multivariable-adjusted model. The association for daytime napping persisted with further adjustment for sleep duration and sleep quality (HR, 1.41, 95% CI, 1.23, 1.62).

### 3.2 Case-control analysis in MVP

Demographic features of participants of MVP by sleep duration are presented in **Supplementary Table 6**. Like our findings from SIMPLER, a U-shaped association between sleep duration and PAD risk was also observed in MVP (**Figure 2B and Table 3**). Compared to individuals with sleep duration of 7 hours/night, the multivariable-adjusted odds ratios (OR) for PAD were 1.38 (95% CI, 1.14-1.67) and 1.48 (95% CI, 1.38-1.59) for those with sleep duration of 5 and 10 hours/night, respectively (**Table 3**). These associations were attenuated after additional adjustment for potential confounders and mediators (**Figure 2B**).

**Table 3.** Association of sleep duration with peripheral artery disease risk in case-control analysis in MVP.

| Sleep duration<br>in hours/day | Unadjusted model |  |  | Adjusted model |  |  |
| --- | --- | --- | --- | --- | --- | --- |
|  | OR | 95% CI | P | OR | 95% CI | P |
| 5 | 1.40 | 1.14, 1.71 | 0.001 | 1.32 | 1.20, 1.44 | 6.63E-09 |
| 6 | 1.18 | 1.11, 1.26 | 2.96E-07 | 1.13 | 1.09, 1.18 | 6.20E-10 |
| 7 | 1.00 |  |  | Ref |  |  |
| 8 | 1.02 | 0.98, 1.06 | 0.278 | 0.99 | 0.95, 1.03 | 0.679 |
| 9 | 1.17 | 1.12, 1.24 | 6.07E-10 | 1.11 | 1.05, 1.17 | 1.07E-04 |
| 10 | 1.46 | 1.37, 1.56 | 4.97E-31 | 1.27 | 1.19, 1.36 | 1.92E-12 |

CI, confidence interval; MVP, the VA Million Veteran Program; OR, odds ratio.
Estimates were adjusted for age, sex, and ancestry-specific genetic principal components in the unadjusted model and further adjusted for self-reported education level, self-reported physical activity, body mass index, hypertension, hemoglobin A1c, smoking history, and statin use in the adjusted model in the case-control study.

### 3.3 Cohort analysis in UKB

The baseline characteristics of UKB participants by sleep duration are shown in **Supplementary Table 7**. A U-shaped association between sleep duration and incident PAD risk was observed (*p* for nonlinearity <0.001, **Figure 2C**). Compared with individuals with sleep duration of 7 to <8 hours per night, the HR of incident PAD were 1.87 (95% CI, 1.50, 2.34) for those with <5 hours of sleep and 1.14 (95% CI, 1.07, 1.22) for those with ≥8 hours of sleep in the model 2 (**Table 4**). The associations attenuated slightly in the model 3 with further adjustment for possible mediators (**Table 4**). Compared with individuals without daytime napping, the HR of PAD was 1.32 (95% CI, 1.25, 1.39) for those taking daytime naps in the multivariable-adjusted model (**Table 4**).

**Table 4.** Associations of sleep duration and daytime napping with risk of peripheral artery disease in the cohort analysis in UKB.

| Sleep trait | Cases | Total | Person-years | Model 1 |  | Model 2 |  | Model 3 |  |
| --- | --- | --- | --- | --- | --- | --- | --- | --- | --- |
|  |  |  |  | HR (95% CI) | *P* | HR (95% CI) | *P* | HR (95% CI) | *P* |
| Sleep duration |  |  |  |  |  |  |  |  |  |
| <5 hours/day | 83 | 3802 | 43 442 | 2.41 (1.93, 3.00) | <0.001 | 1.87 (1.50, 2.34) | <0.001 | 1.64 (1.32, 2.05) | <0.001 |
| ≥5 & <7 hours/day | 1486 | 104 566 | 1 216 659 | 1.52 (1.41, 1.63) | <0.001 | 1.38 (1.28, 1.48) | <0.001 | 1.31 (1.22, 1.40) | <0.001 |
| ≥7 & <8 hours/day | 1658 | 177 596 | 2 087 039 | 1.00 | Ref | 1.00 | Ref | 1.00 | Ref |
| ≥8 hours/day | 2139 | 166 064 | 1 933 724 | 1.20 (1.13, 1.28) | <0.001 | 1.14 (1.07, 1.22) | <0.001 | 1.11 (1.04, 1.18) | 0.002 |
| Daytime napping |  |  |  |  |  |  |  |  |  |
| No | 2162 | 257197 | 3025548 | 1.00 | Ref | 1.00 | Ref | 1.00 | Ref |
| Yes | 3204 | 194831 | 2255316 | 1.45 (1.37, 1.54) | <0.001 | 1.32 (1.25, 1.39) | <0.001 | 1.22 (1.16, 1.29) | <0.001 |

CI, confidence interval; HR, hazard ratio; UKB, UK Biobank.
Model 1 adjusted for age and sex.
Model 2 adjusted for age, sex, education level, smoking status, physical activity, and diet quality.
Model 3 adjusted for the same covariates as in Model 2 and additionally for potential mediators, including body mass index and baseline history of hypertension, hypercholesteremia, and diabetes.

### 3.4 Genetic correlations

Continuous sleep duration, short and long sleep, insomnia, snoring, and daytime napping were genetically correlated with PAD. Short sleep showed the strongest correlation with PAD (*r*_*g*_ =0.29; *p*=4.59×10^−14^), followed by insomnia (*r*_*g*_=0.26; *p*=2.50×10^−11^), snoring (*r*_*g*_=0.18; *p*=4.77×10^−7^), long sleep (*r*_*g*_=0.17; *p*=5.00×10^−4^), daytime napping (*r*_*g*_=0.15; *p*=9.12×10^−7^) and continuous sleep duration (*r*_*g*_=-0.15; *p*=2.00×10^−4^).

### 3.5 MR analysis

We first evaluated the effects of sleep duration on risk of PAD. In our observational analyses, we detected U-shaped associations between sleep duration and PAD. Despite the prospective nature of our SIMPLER and UKB analyses, these may be biased due to residual confounding. To mitigate potential bias from residual confounding and reverse causality, we performed MR to estimate casual relationships between sleep traits and PAD.

We observed an inverse relationship between sleep duration and risk of PAD (**Figure 3A**). For each one-hour increase in genetically predicted sleep duration, risk of PAD was decreased by 30% (OR 0.70, 95% CI, 0.55, 0.89). Because our observational analyses suggested a U-shaped association between sleep duration and PAD, we performed two complementary MR analyses to evaluate for a non-linear association between these traits. First, we considered the effects of short- (< 7 hours) and long- (≥ 9 hours) sleep duration separately. We found that only short sleep (OR 1.20 per one-unit increase in genetically predicted log-transformed OR of having short sleep, 95% CI, 1,04, 1.38) was associated with increased risk of PAD (**Figure 3A**). Second, we performed nonlinear MR. We did not find evidence support a nonlinear association between genetically predicted sleep duration and PAD risk in either MVP (*p* for nonlinearity = 0.460) or UKB (*p* for nonlinearity = 0.098).

**Figure 3.**
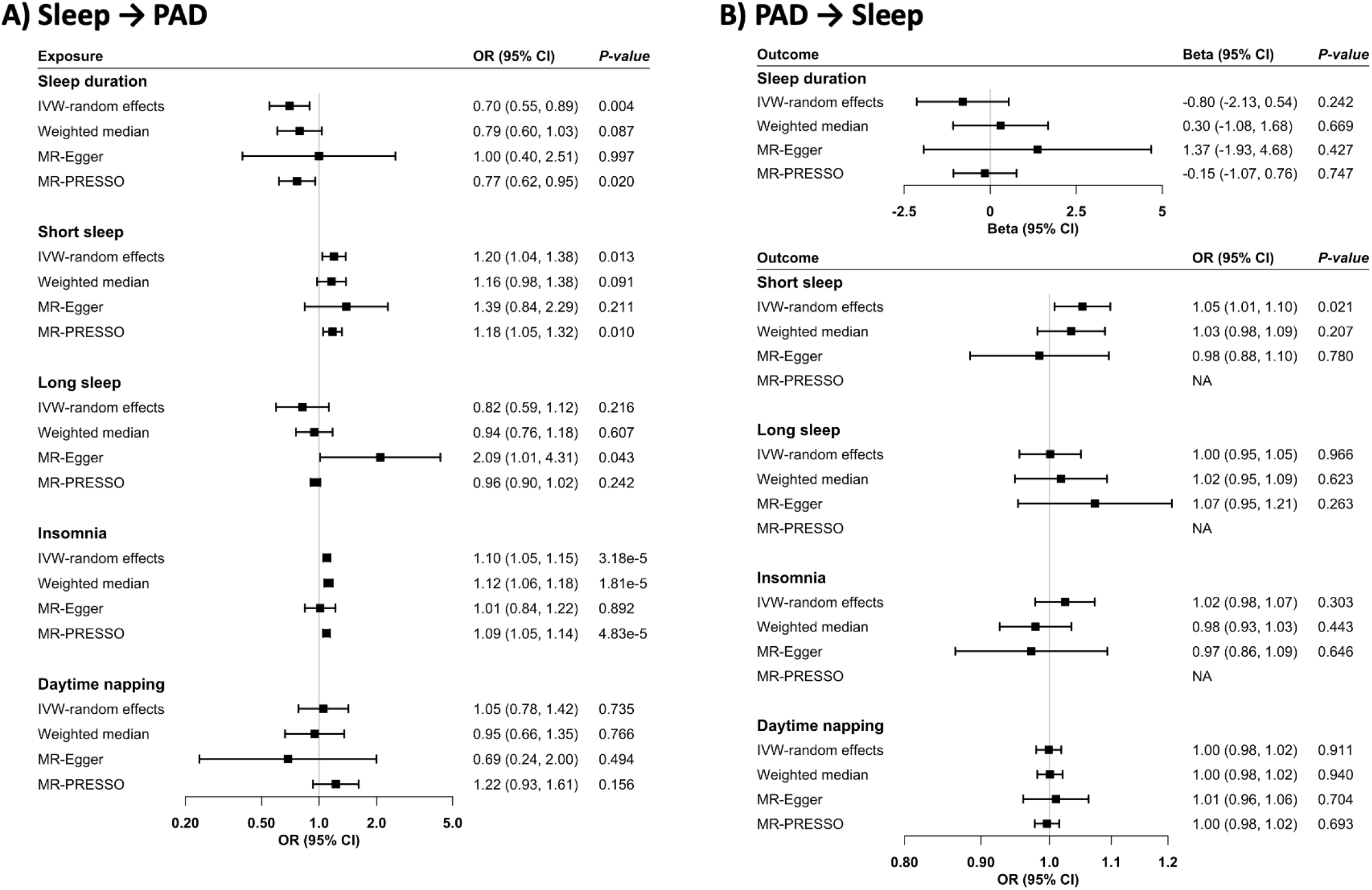
Bidirectional associations between sleep-related traits and risk of peripheral artery disease in Mendelian randomization analysis. Summary-level data for PAD were obtained from the Million Veteran Program, and the corresponding data for sleep-related traits were obtained from published large-scale genome-wide association studies. CI, confidence interval; IVW, inverse variance weighted; OR, odds ratio.

Next, we evaluated the effects of markers of sleep quality on PAD. We detected an association between insomnia and increased risk of PAD (OR 1.10, 95% CI, 1,05, 1.15 per one-unit increase in genetically predicted log-transformed OR of insomnia). No associations were detected between other sleep-related traits and PAD (**Figure 3A and Supplementary Table 9**). Results of sensitivity, heterogeneity, and pleiotropy analyses are presented **Supplementary Table 8**.

Finally, we investigated whether PAD may affect sleep duration or quality. Genetic liability to PAD was associated with an increased risk of having short sleep duration (**Figure 3B**). The OR of having short sleep was 1.05 (95% CI, 1.01, 1.10) per one-unit increase in the log-transformed OR of PAD. This positive association persisted in sensitivity MR analyses and a sensitivity analysis using a PAD genetic instrument from the genome-wide association study excluding UK Biobank participants (**Supplementary Table 10**). We did not observe effects of PAD on other sleep traits (**Supplementary Table 10**).

## 4. Discussion

We performed complementary observational and causal inference analyses among large, diverse cohorts to evaluate the relationship between sleep-related traits and PAD risk. First, in our observational analyses we observed associations between sleep duration and quality with risk of PAD. Next, using genetic variants as instrumental variables to estimate causal effects, we identified significant associations between sleep duration and insomnia with risk of PAD. Our MR analyses largely corroborated our observational findings. Finally, we observed an association between PAD and shorter sleep, suggesting bi-directional relationships between these traits may be important.

Our findings provide clarity to previous studies of sleep duration and PAD risk, which have been limited and conflicting. For example, a prior cross-sectional study including 1,844 participants with a mean age of 68 years found that both short (<6 hours/day) and long (>8 hours/day) sleep were associated with an approximately 2-fold higher prevalence of PAD as compared with those who slept 7 hours per night.^18^ In a cross-sectional study of 3,508 Asian individuals, long but not short sleep was associated with a higher risk of increased arterial stiffness (a risk factor for PAD), but only among male participants.^35^ A recent MR study failed to observe any association between sleep duration and PAD risk in a linear or non-linear fashion in the UK Biobank, but was likely limited by power.^12^ Although at least two well-powered MR studies have demonstrated a clear adverse impact of short sleep on coronary artery disease,^12,36^ effects of cardiovascular risk factors may vary by vascular bed,^37-39^ limiting direct translation of these findings to PAD. Due to the relatively small numbers of participants and PAD cases, these prior studies may have been limited by low statistical power, and observational analyses may have been further limited by residual confounding and reverse causality.

We detected a strong association between sleep duration and PAD across our case-control, cohort, and MR analyses. This triangulation of evidence provides strong support for a causal relationship between short sleep duration and increased risk of PAD. In contrast, our analyses identified discordant effects between increased/long sleep duration and risk of PAD. For example, our cohort and case-control studies found a U-shaped association between sleep duration and increased risk of PAD. However, our MR analyses, which are less susceptible to bias from residual confounding and reverse causality, suggested that the relationship between sleep duration and PAD was linear. Our MR did not support our observational finding that increased sleep duration might be detrimental. Reverse causality, whereby PAD-related pain leads to poor sleep quality but increased time in bed might represent an explanation for the discordance between our observational and MR results. Our reverse-MR analysis provides support for such a bi-directional relationship, where increased liability to PAD leads to shorter actigraphy-assessed sleep duration. These results provide consistent support for short sleep duration as a harmful risk factor for PAD and highlight the possibility of reverse causality leading to distorted effects in observational analyses of sleep and PAD.

Several underlying mechanisms may explain the associations of short sleep duration with increased PAD. Short sleep may activate autonomic nervous system with global sympathetic overactivity, increase oxidative stress, inflammation and endothelial dysfunction and impair metabolic regulation and coagulation system activity and therefore increase PAD risk.^10^ In addition, meta-analyses have found that unfavorable sleep duration, mostly short sleep, were associated with several cardiometabolic factors that are important risk factors for PAD, including obesity, hypertension, and type 2 diabetes.^2^ The specific factors that mediate the effects of sleep traits on PAD requires future study.

Our findings also support a role for sleep quality, in addition to sleep duration, in the pathogenesis of PAD. In our observational analyses, we observed a positive association between poor sleep quality based on a composite of 6 sleep features and increased PAD risk, which is a novel finding and needs conformation. Daytime napping has been associated with an increased risk of cardiovascular disease and mortality in recent years,^40^ and a prior MR study has suggested daytime napping might unfavorably influence cardiometabolic factors such as blood pressure and waist circumference.^41^ Although we detected an elevated risk of incident PAD related to daytime napping in our cohort study, this association was not verified by MR. Our MR analysis provided support for insomnia as a risk factor for PAD, which was also observed in one of our previous studies based on data from the UK Biobank study.^13^ Although sleep apnea has been identified as a risk factor for PAD,^19,42^ this association was not confirmed in our cohort or MR analyses. This result is consistent with a prior meta-analysis, which did not find a clear association of sleep apnea with coronary artery disease.^43^ We studied five other sleep features in relation to incident PAD and found no strong associations. The mechanism for the positive association between poor sleep quality and PAD may be related to fragmentation, which may accelerate atherosclerosis and elevate the risk of cardiovascular disease.^44,45^

Our findings have potential clinical implications for diagnosis and management of both PAD and sleep disorders. We observe a bidirectional relationship between sleep duration and PAD, suggesting feed-forward mechanisms may accelerate both sets of diseases. Although we did not directly evaluate rates of comorbid sleep disorders and PAD, our findings raise the possibility that screening for PAD among those with poor sleep, and for poor sleep among those with PAD merits future study. Similarly, our findings suggest that interventions to improve sleep may have downstream effects on PAD, and likewise interventions to treat PAD may improve sleep. Although the optimal interventions for interrupting the sleep-PAD link is not yet known, the American Heart Association has identified sleep research as an important priority for improving cardiometabolic health.^2^

There are several strengths of the present study. We used complementary case-control, prospective cohort, and MR study designs to provide convergent evidence and increase confidence in results. For the cohort studies, a large number of incident cases, the availability of data on important confounders, and objective diagnostic information for PAD from a national register made our observational findings reliable. Further, although latitude (and resultant differences in daily sunlight exposure) may influence sleep,^46^ our observational cohorts spanned the northern hemisphere, suggesting these differences in sunlight exposure are unlikely to explain the link between sleep and PAD. With regard to MR analysis, we selected instrumental variables for sleep-related traits from the largest available genome-wide association studies^41,47-50^ and used summary-level data for PAD from a GWAS including nearly 30,000 more PAD cases compared to a prior MR study of sleep traits and PAD^12^, which increased our statistical power to detect meaningful associations.

Some limitations should be considered when interpreting our findings. In our cohort studies, data on sleep habits and covariates were obtained from self-administrated questionnaires completed by participants at start of follow-up. Lifestyle habits are prone to changes, and it is conceivable that our estimates were affected by some degree of measurement errors. However, any misclassification of participants into the wrong exposure or covariate category would be expected to attenuate the observed associations toward the null as participants were free of

PAD at the time of exposure measurement. Our cohort analyses were based on two cohorts of middle-aged and older individuals. Whether the definition on short and long sleep and corresponding associations with PAD can be generalized to younger populations is unknown. Similarly, the influence of sleep across the lifespan on PAD risk requires further study. For our MR analyses, we considered individuals of diverse ancestries, although there was over-representation of individuals of European descent (77% European participants in the Million Veteran Program). Thus, whether our findings are applicable to other populations warrants further study. Although we could not completely rule out pleiotropic effects our in MR analysis, our findings were robust across MR methods that make different assumptions about pleiotropy. Genetic instruments for sleep-related traits were based on measurements from the UK Biobank, which includes relatively healthy participants, and this may also limit the generalizability of our findings.

In conclusion, across several complementary observational and MR analyses, we find sleep duration and quality play an important role in the development of PAD and may represent an opportunity for improved evaluation and treatment.

## Data Availability

De-identified SIMPLER data are available for researchers upon application (http://www.simpler4health.se/). Summary data from the MVP GWAS of PAD can be obtained via dbGaP Study Ascession: phs001672. Access to the UK Biobank data can be obtained upon application (https://www.ukbiobank.ac.uk/). Data used in two-sample Mendelian randomization analysis are available in Supplementary materials.

## Additional information

## Acknowledgement

We want to acknowledge the participants and investigators of SIMPLER for provisioning of facilities and experimental support. SIMPLER receives funding through the Swedish Research Council under grant number 2017-00644. The computations were performed on resources provided by SNIC through Uppsala Multidisciplinary Center for Advanced Computational Science (UPPMAX) under Project simpl2020002. We also want to acknowledge the participants and investigators of the VA Million Veteran Program. This research was conducted using the UK Biobank study under Application Number 66354.

## Sources of funding

Funding for this study came from the Karolinska Institutet’s Research Foundation Grants (Grant number 2020-01842), the Swedish Research Council (Vetenskapsrådet; Grant Number 2019-00977), the Swedish Research Council for Health, Working Life and Welfare (Forte; 2018-00123) and the Swedish Heart-Lung Foundation (Hjärt-Lungfonden; Grant number 20190247). M.G.L. is supported by the Institute for Translational Medicine and Therapeutics of the Perelman School of Medicine at the University of Pennsylvania and the NIH/NHLBI National Research Service Award postdoctoral fellowship (T32HL007843). S.M.D. is supported by IK2-CX001780. This publication does not represent the views of the Department of Veterans Affairs or the United States Government.

## Disclosures

All authors declare no conflict of interest.

## Author contributions

S.Y., M.G.L., S.M.D, and S.C.L. conceived and designed the study. S.Y., M.G.L., Y.S., and X.L. undertook the statistical analyses. S.Y. and M.G.L. wrote the first draft of the manuscript. All authors provided important comments to the manuscript and approved the final version of the manuscript.

## Notes

### Competing Interest Statement

The authors have declared no competing interest.

### Funding Statement

Funding for this study came from the Karolinska Institutet Research Foundation Grants (Grant number 2020-01842), the Swedish Research Council (Grant Number 2019-00977), the Swedish Research Council for Health, Working Life and Welfare (2018-00123) and the Swedish Heart-Lung Foundation (Grant number 20190247). M.G.L. is supported by the Institute for Translational Medicine and Therapeutics of the Perelman School of Medicine at the University of Pennsylvania and the NIH/NHLBI National Research Service Award postdoctoral fellowship (T32HL007843). S.M.D. is supported by IK2-CX001780. This publication does not represent the views of the Department of Veterans Affairs or the United States Government.

### Author Declarations

The study has been approved by the Swedish Ethical Review Authority (no. 2019-03986) and the United States VA Central IRB. The UK Biobank received ethical permits from the North West Multi-centre Research Ethics Committee, the National Information Governance Board for Health and Social Care in England and Wales, and the Community Health Index Advisory Group in Scotland. All participants have provided informed consent.

