## Supplementary Table for "Sleep Duration, Quality, and Risk of Peripheral Artery Disease: Multinational Cohort and Mendelian Randomization Studies": Supporting information.docx

### **Supplementary methods**

**Description of two cohorts in SIMPLER**

The cohort analysis used data from SIMPLER (<https://www.simpler4health.se/>), which includes the Swedish Mammography Cohort and the Cohort of Swedish Men. The Swedish Mammography Cohort was initiated in 1987 and three sequent surveys were conducted in 1997, 2008-2009, and 2019. The Cohort of Swedish Men was established in 1997. Likewise, participants in this cohort were revisited in 2008-2009 and 2019. Participants were asked to fill in questionnaires on health status and lifestyle factors with a few sex-specific questions between two cohorts. Incident PAD cases were ascertained by the clinical diagnosis based on codes from the 9th (440.0, 440.2, 440.3, 440.4, 440.9, 443.9) and 10th (I70.0, I70.2, I70.3, I70.4, I70.5, I70.6, I70.7, I70.9, I703.9) revision of the International Classification of Diseases (ICD).

**Sleep quality measurement in SIMPLER**

Sleep quality was assessed by a sleep quality score based on six sleep measures. The six sleep measures included “Difficulty falling asleep”, “Repeatedly waking up with difficulty falling asleep”, “Premature awakening”, “Disturbed or restless sleep”, “Sleep apnea”, and “Disturbing snoring”. Participants were asked to define the frequency (never, seldom, often, mostly, and always) of each sleep problem. We constructed a score including these 6 sleep complaints to represent overall sleep quality. For each sleep problem, the participant was assigned one point when they reported often, mostly, or always for the specified sleep problem. Otherwise, they got zero. The sum of sleep quality score ranged from 0 to 6, with a higher score representing more sleep complaints.

**Covariates in SIMPLER**

Covariates included age (continuous in year), sex, education level (≤9, 10 to 12, >12 years), smoking (never smoker, past smoker with <20, 20-39 and ≥ 40 pack-years and current smoker with <20, 20-39 and ≥ 40 pack-years), physical activity (0-10, 11-30 and 31-60 and >60 minutes per day of walking or cycling) and diet quality as obtained from the 2008 and 2009 questionnaires. Diet quality was assessed by a modified version of the Dietary Approaches to Stop Hypertension diet. We also obtained data on baseline body weight and height and calculated body mass index (height divided by weight squared) as well as data on history of hypertension, hypercholesterolemia, and diabetes, which were regarded as potential mediators.

**Modified version of the Dietary Approaches to Stop Hypertension diet measurement in SIMPLER**

Diet quality was assessed by a modified version of the Dietary Approaches to Stop Hypertension diet. This dietary quality score included fruits, vegetables, nuts and legumes, whole grains, and low‐fat dairy products as healthy components and red and processed meat and sweetened beverages as unhealthy components.^1^ Individuals were assigned a score from 1 to 5 according to the quintiles of consumption of each food and the scores were summed to create a diet score (7 to 35). A high score indicates a high adherence to the modified Dietary Approaches to Stop Hypertension diet pattern.

1.	Yuan S, Bruzelius M, Håkansson N, Åkesson A, Larsson SC. Lifestyle factors and venous thromboembolism in two cohort studies. *Thrombosis Research*. 2021. doi: <https://doi.org/10.1016/j.thromres.2021.03.024>

### **Supplementary Table 1. Information on used studies in the Mendelian randomization analyses**

| **Exposure or outcome** | **Definition** | **Participants included in analysis** | **Adjustments** | **Identified SNPs** | **Instrument variables** |
| --- | --- | --- | --- | --- | --- |
| Continuous sleep duration^a^ | Hours | 446 118 individuals from UK Biobank | Age, sex, 10 principal components of ancestry, genotyping array, and genetic correlation matrix | 78 | 73 |
| Short sleep^a^ | < 7 hours | 106 192 cases and 305 742 controls from UK Biobank | Age, sex, 10 principal components of ancestry, genotyping array, and genetic correlation matrix | 27 | 27 |
| Long sleep^a^ | ≥ 9 hours | 34 184 cases and 305 742 controls from UK Biobank | Age, sex, 10 principal components of ancestry, genotyping array, and genetic correlation matrix | 8 | 8 |
| Sleep apnea^b^ | By either self-reported diagnostic item, a self-reported diagnosis or a general practitioner diagnosis based on ICD10 codes | 25 062 cases and 485 422 controls of European ancestry | Age, sex, batch (where relevant), and genetic ancestry principal components | 37 | 35 |
| Snoring^c^ | Self-reported information on snoring | 15 200 cases and 256 00 controls from UK Biobank | Age, sex, genotyping array, and the first 20 genetic principal components | 28 | 27 |
| Insomnia^d^ | Self-reported information on insomnia | 397,959 cases and 933,051 controls from UK Biobank and 23andMe | Age, sex, genotyping array, and genetic principal components | 248 | 208 |
| Daytime napping^e^ | Do you have a nap during the day? never/rarely, sometimes, usually, prefer not to answer (treated as a continuous variable) | 452 633 individuals from UK Biobank and 541 333 individuals from 23andMe | Age, sex, 10 principal components of ancestry, genotyping array, and genetic correlation matrix | 123 | 105 |
| Peripheral artery disease (instrument)^f^ | By at least two of the ICD-9/10 codes/Current Procedural Terminology codes | 36 424 cases and 601 044 controls (>77% of European ancestry) | Age, sex and five principal components | 19 | 19 |
| Peripheral artery disease^f^ | By at least two of the ICD-9/10 codes/Current Procedural Terminology codes | 31 307 cases and 211 753 controls of multi-ancestries (77% of European ancestry) | Age, sex and five principal components | - | - |

SNPs, single nucleotide polymorphisms. Instrument variable was selected by setting *r*^2^<0.01 and clumping window > 10 000 kb.

^a^ Dashti HS, Jones SE, Wood AR, Lane JM, van Hees VT, Wang H, Rhodes JA, Song Y, Patel K, Anderson SG, Beaumont RN, Bechtold DA, Bowden J, Cade BE, Garaulet M, Kyle SD, Little MA, Loudon AS, Luik AI, Scheer F, Spiegelhalder K, Tyrrell J, Gottlieb DJ, Tiemeier H, Ray DW, Purcell SM, Frayling TM, Redline S, Lawlor DA, Rutter MK, Weedon MN and Saxena R. Genome-wide association study identifies genetic loci for self-reported habitual sleep duration supported by accelerometer-derived estimates. Nat Commun. 2019;10:1100.

^b^ Campos AI, Ingold N, Huang Y, Kho P-F, Han X, Ong J-S, García-Marín LM, , Law MH, Martin NG, Dong X, Cuellar-Partida G, MacGregor S, Aslibekyan S and Rentería ME. Genome-wide analyses in 1,987,836 participants identify 39 genetic loci associated with sleep apnoea. medRxiv. 2020. doi: <https://doi.org/10.1101/2020.09.29.20199893>.

^c^ Campos AI, García-Marín LM, Byrne EM, Martin NG, Cuéllar-Partida G, Rentería ME. Insights into the aetiology of snoring from observational and genetic investigations in the UK Biobank. Nat Commun. 2020 Feb 14;11(1):817.

^d^ Jansen PR, Watanabe K, Stringer S, Skene N, Bryois J, Hammerschlag AR, de Leeuw CA, Benjamins JS, Muñoz-Manchado AB, Nagel M, Savage JE, Tiemeier H, White T; 23andMe Research Team, Tung JY, Hinds DA, Vacic V, Wang X, Sullivan PF, van der Sluis S, Polderman TJC, Smit AB, Hjerling-Leffler J, Van Someren EJW, Posthuma D. Genome-wide analysis of insomnia in 1,331,010 individuals identifies new risk loci and functional pathways. Nat Genet. 2019;51(3):394-403.

^e^ Dashti HS, Daghlas I, Lane JM, Huang Y, Udler MS, Wang H, Ollila HM, Jones SE, Kim J, Wood AR, Weedon MN, Aslibekyan S, Garaulet M and Saxena R. Genetic determinants of daytime napping and effects on cardiometabolic health. Nat Commun. 2021;12:900.

^f^ Klarin D, Lynch J, Aragam K, Chaffin M, Assimes TL, Huang J, Lee KM, Shao Q, Huffman JE, Natarajan P, Arya S, Small A, Sun YV, Vujkovic M, Freiberg MS, Wang L, Chen J, Saleheen D, Lee JS, Miller DR, Reaven P, Alba PR, Patterson OV, DuVall SL, Boden WE, Beckman JA, Gaziano JM, Concato J, Rader DJ, Cho K, Chang KM, Wilson PWF, O'Donnell CJ, Kathiresan S, Tsao PS and Damrauer SM. Genome-wide association study of peripheral artery disease in the Million Veteran Program. Nat Med. 2019;25:1274-1279.

### **Supplementary Table 2. Instrumental variables for sleep-related traits and their associations with peripheral artery disease**

| **Trait** | **SNP** | **Chr** | **Position** | **Nearby gene** | **EA** | **NEA** | **EAF** | **Sleep-related habit** | | | **PAD** | | |
| --- | --- | --- | --- | --- | --- | --- | --- | --- | --- | --- | --- | --- | --- |
|  |  |  |  |  |  |  |  | **Beta** | **SE** | **P** | **Beta** | **SE** | **P** |
| Napping | rs12031519 | 1 | 162876200 | *CCDC190, RGS4* | A | G | 0.89 | 0.011 | 0.002 | 5.50E-09 | 0.036 | 0.016 | 0.021 |
| Napping | rs12140153 | 1 | 62579891 | *PATJ* | G | T | 0.92 | 0.025 | 0.002 | 2.40E-31 | 0.043 | 0.020 | 0.036 |
| Napping | rs1843815 | 1 | 98589715 | *DPYD, SNX7* | T | A | 0.49 | 0.007 | 0.001 | 3.30E-09 | 0.011 | 0.010 | 0.238 |
| Napping | rs1931175 | 1 | 96928273 | *PTBP2* | G | C | 0.37 | 0.008 | 0.001 | 4.60E-10 | 0.012 | 0.010 | 0.239 |
| Napping | rs2250377 | 1 | 201860626 | *SHISA4* | A | G | 0.31 | 0.013 | 0.001 | 1.40E-24 | -0.020 | 0.010 | 0.042 |
| Napping | rs2786547 | 1 | 33342304 | *FNDC5, HPCA* | C | T | 0.83 | 0.011 | 0.002 | 1.00E-11 | 0.019 | 0.013 | 0.145 |
| Napping | rs2893323 | 1 | 95783037 | *RWDD3* | A | G | 0.33 | 0.007 | 0.001 | 3.30E-08 | -0.003 | 0.010 | 0.779 |
| Napping | rs11125776 | 2 | 59478517 | *LINC01793* | T | G | 0.85 | 0.012 | 0.002 | 1.50E-11 | -0.002 | 0.013 | 0.880 |
| Napping | rs12614085 | 2 | 151501545 | *RND3, RBM43* | C | T | 0.09 | 0.012 | 0.002 | 2.00E-08 | -0.008 | 0.016 | 0.612 |
| Napping | rs13023587 | 2 | 49413860 | *FSHR, STON1-GTF2A1L* | C | G | 0.47 | 0.007 | 0.001 | 6.50E-10 | 0.005 | 0.009 | 0.554 |
| Napping | rs13033444 | 2 | 23611438 | *KLHL29* | G | A | 0.26 | 0.010 | 0.001 | 3.60E-13 | 0.011 | 0.011 | 0.318 |
| Napping | rs17049683 | 2 | 58905715 | *FANCL* | G | A | 0.31 | 0.008 | 0.001 | 4.10E-10 | -0.016 | 0.011 | 0.137 |
| Napping | rs350785 | 2 | 52934600 | *GPR75-ASB3, ASB3* | T | C | 0.11 | 0.013 | 0.002 | 1.50E-11 | -0.033 | 0.015 | 0.028 |
| Napping | rs35144585 | 2 | 169092428 | *STK39* | T | A | 0.87 | 0.010 | 0.002 | 1.20E-08 | -0.009 | 0.015 | 0.544 |
| Napping | rs3732085 | 2 | 206831860 | *INO80D, NDUFS1, NRP2* | C | A | 0.66 | 0.007 | 0.001 | 1.50E-08 | -0.017 | 0.010 | 0.082 |
| Napping | rs56180058 | 2 | 25337155 | *EFR3B* | C | T | 0.86 | 0.009 | 0.002 | 1.30E-08 | -0.010 | 0.015 | 0.506 |
| Napping | rs62189006 | 2 | 162573586 | *SLC4A10* | A | G | 0.91 | 0.012 | 0.002 | 8.30E-09 | 0.011 | 0.017 | 0.498 |
| Napping | rs7422655 | 2 | 208941004 | *CRYGD, PLEKHM3* | C | T | 0.34 | 0.008 | 0.001 | 1.10E-08 | -0.005 | 0.010 | 0.600 |
| Napping | rs80163246 | 2 | 164574344 | *FIGN* | C | T | 0.10 | 0.012 | 0.002 | 1.40E-10 | -0.002 | 0.016 | 0.896 |
| Napping | rs9309116 | 2 | 44557919 | *PREPL* | T | C | 0.67 | 0.007 | 0.001 | 3.90E-09 | 0.020 | 0.010 | 0.048 |
| Napping | rs1001817 | 3 | 183995341 | *ECE2* | C | T | 0.48 | 0.008 | 0.001 | 1.70E-10 | -0.003 | 0.009 | 0.715 |
| Napping | rs1601440 | 3 | 84665393 | *CADM2* | C | T | 0.28 | 0.009 | 0.001 | 1.90E-11 | 0.000 | 0.010 | 0.992 |
| Napping | rs253666 | 3 | 138115770 | *MRAS* | A | G | 0.78 | 0.008 | 0.001 | 1.80E-08 | -0.026 | 0.011 | 0.024 |
| Napping | rs2699869 | 3 | 133032892 | *TMEM108* | A | C | 0.45 | 0.007 | 0.001 | 3.10E-08 | -0.011 | 0.010 | 0.270 |
| Napping | rs76824303 | 3 | 62459819 | *CADPS* | A | C | 0.91 | 0.012 | 0.002 | 1.70E-08 | 0.047 | 0.018 | 0.009 |
| Napping | rs77154532 | 3 | 19377311 | *KCNH8* | A | G | 0.65 | 0.008 | 0.001 | 1.40E-09 | 0.000 | 0.010 | 0.968 |
| Napping | rs936944 | 3 | 44385814 | *TCAIM* | G | A | 0.15 | 0.009 | 0.002 | 3.20E-08 | 0.017 | 0.013 | 0.207 |
| Napping | rs9883093 | 3 | 82774831 | *GBE1* | G | T | 0.59 | 0.007 | 0.001 | 4.50E-09 | 0.005 | 0.009 | 0.575 |
| Napping | rs4356873 | 4 | 164216739 | *NPY1R, NAF1* | C | T | 0.26 | 0.008 | 0.001 | 4.50E-08 | 0.003 | 0.011 | 0.818 |
| Napping | rs4692709 | 4 | 170228549 | *SH3RF1, NEK1* | C | T | 0.49 | 0.007 | 0.001 | 2.40E-09 | -0.002 | 0.009 | 0.861 |
| Napping | rs9998136 | 4 | 79550425 | *ANXA3, BMP2K* | G | C | 0.71 | 0.009 | 0.001 | 4.40E-10 | -0.002 | 0.010 | 0.852 |
| Napping | rs10875606 | 5 | 143769614 | *KCTD16* | C | A | 0.35 | 0.007 | 0.001 | 1.30E-08 | -0.004 | 0.010 | 0.660 |
| Napping | rs10875622 | 5 | 146693062 | *STK32A* | A | G | 0.57 | 0.010 | 0.001 | 1.30E-17 | 0.012 | 0.009 | 0.188 |
| Napping | rs12657723 | 5 | 62760828 | *HTR1A, RNF180* | T | C | 0.30 | 0.008 | 0.001 | 2.00E-10 | 0.020 | 0.011 | 0.057 |
| Napping | rs2099810 | 5 | 112030173 | *APC, REEP5* | A | G | 0.53 | 0.008 | 0.001 | 2.80E-10 | 0.003 | 0.010 | 0.738 |
| Napping | rs2195272 | 5 | 102295504 | *PAM* | T | G | 0.30 | 0.010 | 0.001 | 6.70E-15 | -0.008 | 0.010 | 0.403 |
| Napping | rs2431108 | 5 | 103947968 | *RP11-6N13.1* | C | T | 0.31 | 0.013 | 0.001 | 7.70E-24 | -0.013 | 0.010 | 0.214 |
| Napping | rs2943023 | 5 | 89597733 | *CETN3* | C | T | 0.58 | 0.007 | 0.001 | 7.40E-09 | 0.026 | 0.009 | 0.005 |
| Napping | rs388016 | 5 | 106861892 | *EFNA5* | A | G | 0.63 | 0.007 | 0.001 | 6.10E-09 | -0.007 | 0.009 | 0.476 |
| Napping | rs4357022 | 5 | 106311039 | *EFNA5* | G | T | 0.47 | 0.007 | 0.001 | 1.40E-08 | 0.000 | 0.009 | 0.972 |
| Napping | rs6452787 | 5 | 87712831 | *TMEM161B* | A | G | 0.48 | 0.008 | 0.001 | 2.00E-10 | 0.007 | 0.009 | 0.470 |
| Napping | rs73817091 | 5 | 159042652 | *IL12B, ADRA1B* | T | C | 0.03 | 0.017 | 0.003 | 3.80E-08 | 0.065 | 0.031 | 0.033 |
| Napping | rs11967137 | 6 | 28199764 | *ZSCAN9* | A | G | 0.79 | 0.009 | 0.002 | 1.70E-09 | -0.029 | 0.011 | 0.009 |
| Napping | rs140506252 | 6 | 155129280 | *SCAF8* | A | T | 0.98 | 0.023 | 0.004 | 4.30E-08 | -0.058 | 0.035 | 0.096 |
| Napping | rs2143792 | 6 | 62182204 | *MTRNR2L9* | G | A | 0.58 | 0.007 | 0.001 | 9.10E-09 | 0.004 | 0.011 | 0.710 |
| Napping | rs2653349 | 6 | 55142337 | *HCRTR2* | A | G | 0.19 | 0.017 | 0.001 | 3.40E-29 | 0.019 | 0.011 | 0.103 |
| Napping | rs34262487 | 6 | 36224315 | *PNPLA1* | C | A | 0.93 | 0.015 | 0.002 | 1.20E-09 | -0.007 | 0.019 | 0.727 |
| Napping | rs4236060 | 6 | 38470087 | *BTBD9* | T | C | 0.26 | 0.009 | 0.001 | 8.80E-11 | 0.007 | 0.011 | 0.503 |
| Napping | rs614987 | 6 | 124920871 | *NKAIN2* | C | A | 0.55 | 0.011 | 0.001 | 7.50E-19 | -0.011 | 0.009 | 0.240 |
| Napping | rs6919087 | 6 | 37699156 | *MDGA1, ZFAND3* | T | G | 0.69 | 0.011 | 0.001 | 1.00E-16 | -0.002 | 0.010 | 0.838 |
| Napping | rs9389556 | 6 | 100079774 | *PRDM13, MCHR2, CCNC* | G | C | 0.27 | 0.008 | 0.001 | 9.70E-10 | 0.016 | 0.010 | 0.133 |
| Napping | rs9460110 | 6 | 170621249 | *FAM120B* | C | T | 0.34 | 0.007 | 0.001 | 8.40E-09 | -0.003 | 0.010 | 0.766 |
| Napping | rs10257273 | 7 | 99188589 | *GS1-259H13.13* | A | T | 0.71 | 0.011 | 0.002 | 8.90E-10 | 0.009 | 0.012 | 0.474 |
| Napping | rs35851551 | 7 | 31330785 | *NEUROD6, ADCYAP1R1* | A | G | 0.92 | 0.011 | 0.002 | 3.50E-08 | -0.011 | 0.018 | 0.530 |
| Napping | rs13263535 | 8 | 1175131 | *DLGAP2, ERICH1* | G | T | 0.48 | 0.007 | 0.001 | 1.60E-08 | 0.010 | 0.009 | 0.281 |
| Napping | rs2059639 | 8 | 67381858 | *ADHFE1, C8orf46* | C | T | 0.43 | 0.007 | 0.001 | 2.40E-08 | -0.011 | 0.009 | 0.229 |
| Napping | rs285815 | 8 | 106108303 | *ZFPM2, LRP12* | T | A | 0.41 | 0.008 | 0.001 | 5.80E-10 | -0.002 | 0.009 | 0.849 |
| Napping | rs351776 | 8 | 28191306 | *PNOC* | C | A | 0.50 | 0.008 | 0.001 | 8.40E-10 | 0.008 | 0.009 | 0.384 |
| Napping | rs7814873 | 8 | 142213997 | *SLC45A4, DENND3* | C | T | 0.36 | 0.007 | 0.001 | 1.50E-08 | -0.017 | 0.010 | 0.092 |
| Napping | rs10811438 | 9 | 20962282 | *FOCAD* | G | C | 0.62 | 0.007 | 0.001 | 3.20E-09 | 0.000 | 0.009 | 0.979 |
| Napping | rs12346996 | 9 | 131840856 | *DOLPP1, FAM73B* | T | C | 0.25 | 0.008 | 0.001 | 5.00E-09 | 0.004 | 0.011 | 0.753 |
| Napping | rs13284688 | 9 | 81727018 | *TLE4, PSAT1* | C | T | 0.20 | 0.015 | 0.001 | 1.70E-23 | -0.006 | 0.011 | 0.571 |
| Napping | rs1415218 | 9 | 73504992 | *TRPM3* | T | C | 0.28 | 0.008 | 0.001 | 1.20E-09 | 0.005 | 0.010 | 0.658 |
| Napping | rs17502738 | 9 | 37441650 | *ZBTB5* | T | C | 0.82 | 0.009 | 0.002 | 2.00E-08 | 0.031 | 0.012 | 0.011 |
| Napping | rs4604518 | 9 | 120555845 | *TLR4* | G | A | 0.54 | 0.007 | 0.001 | 1.40E-08 | -0.005 | 0.010 | 0.655 |
| Napping | rs62560863 | 9 | 34081331 | *DCAF12, UBAP2* | T | C | 0.11 | 0.011 | 0.002 | 2.50E-08 | 0.005 | 0.016 | 0.737 |
| Napping | rs971415 | 9 | 108846445 | *TMEM38B, FKTN* | A | G | 0.88 | 0.011 | 0.002 | 1.20E-09 | 0.016 | 0.014 | 0.277 |
| Napping | rs11258652 | 10 | 13868855 | *FRMD4A* | C | A | 0.78 | 0.010 | 0.001 | 3.70E-13 | 0.001 | 0.011 | 0.916 |
| Napping | rs224111 | 10 | 64552010 | *ADO, EGR2, ZNF365* | G | A | 0.61 | 0.008 | 0.001 | 1.60E-10 | -0.013 | 0.009 | 0.175 |
| Napping | rs10835420 | 11 | 28866669 | *METTL15* | T | A | 0.75 | 0.009 | 0.001 | 1.70E-10 | 0.016 | 0.010 | 0.118 |
| Napping | rs11224896 | 11 | 101479583 | *TRPC6, ANGPTL5* | T | C | 0.90 | 0.011 | 0.002 | 1.10E-08 | 0.003 | 0.016 | 0.848 |
| Napping | rs174541 | 11 | 61565908 | *FADS2, FEN1* | C | T | 0.33 | 0.010 | 0.001 | 4.40E-15 | -0.053 | 0.010 | 0.000 |
| Napping | rs271057 | 11 | 92769748 | *MTNR1B, SLC36A4* | C | T | 0.33 | 0.009 | 0.001 | 6.50E-10 | 0.001 | 0.010 | 0.905 |
| Napping | rs11615756 | 12 | 117942076 | *KSR2* | T | C | 0.38 | 0.018 | 0.001 | 0.00E+00 | -0.003 | 0.010 | 0.751 |
| Napping | rs2417268 | 12 | 13493906 | *EMPI1, C12orf36* | T | A | 0.43 | 0.008 | 0.001 | 1.20E-09 | 0.011 | 0.009 | 0.251 |
| Napping | rs35011311 | 12 | 38616581 | *ALG10B* | G | T | 0.89 | 0.009 | 0.001 | 4.40E-11 | 0.052 | 0.041 | 0.203 |
| Napping | rs60222088 | 12 | 108334477 | *ASCL4, PRDM4* | C | A | 0.82 | 0.011 | 0.002 | 6.40E-11 | 0.003 | 0.013 | 0.821 |
| Napping | rs2769916 | 13 | 107820389 | *FAM155A* | A | G | 0.71 | 0.009 | 0.001 | 1.70E-11 | 0.002 | 0.010 | 0.886 |
| Napping | rs10149986 | 14 | 29721044 | *PRKD1, FOXG1* | G | T | 0.19 | 0.011 | 0.002 | 4.40E-12 | -0.001 | 0.012 | 0.912 |
| Napping | rs2370926 | 14 | 79602542 | *NRXN3* | T | C | 0.64 | 0.008 | 0.001 | 1.70E-10 | -0.010 | 0.009 | 0.279 |
| Napping | rs4983329 | 14 | 29218473 | *FOXG1* | A | C | 0.54 | 0.008 | 0.001 | 3.10E-10 | 0.007 | 0.009 | 0.468 |
| Napping | rs10152428 | 15 | 93510243 | *CHD2* | C | G | 0.66 | 0.008 | 0.001 | 1.10E-08 | 0.015 | 0.010 | 0.150 |
| Napping | rs11071755 | 15 | 63793936 | *USP3, CA12* | G | A | 0.60 | 0.007 | 0.001 | 5.20E-09 | 0.007 | 0.011 | 0.538 |
| Napping | rs17158413 | 15 | 83235408 | *CPEB1* | A | G | 0.23 | 0.009 | 0.001 | 4.40E-11 | -0.002 | 0.011 | 0.853 |
| Napping | rs1592544 | 16 | 56129244 | *GNAO1, CES5A* | C | T | 0.51 | 0.009 | 0.001 | 8.50E-13 | 0.012 | 0.009 | 0.188 |
| Napping | rs3986805 | 16 | 28350059 | *NPIPB6* | G | A | 0.43 | 0.007 | 0.001 | 8.70E-09 | 0.009 | 0.009 | 0.343 |
| Napping | rs528301822 | 16 | 69403012 | *TERF2* | T | A | 0.30 | 0.008 | 0.001 | 2.80E-09 | 0.001 | 0.010 | 0.957 |
| Napping | rs60920123 | 16 | 10134637 | *GRIN2A* | G | A | 0.58 | 0.008 | 0.001 | 4.50E-10 | -0.010 | 0.009 | 0.274 |
| Napping | rs9939355 | 16 | 23837048 | *PRKCB, CHP2* | C | T | 0.49 | 0.007 | 0.001 | 4.50E-09 | 0.005 | 0.009 | 0.593 |
| Napping | rs112520848 | 17 | 64308310 | *PRKCA* | C | G | 0.38 | 0.007 | 0.001 | 1.80E-08 | -0.002 | 0.009 | 0.862 |
| Napping | rs12451365 | 17 | 35595368 | *ACACA* | C | T | 0.31 | 0.011 | 0.002 | 1.50E-12 | 0.015 | 0.011 | 0.157 |
| Napping | rs385199 | 17 | 43686419 | *CRHR1, PLEKHM1* | A | C | 0.78 | 0.021 | 0.001 | 0.00E+00 | -0.036 | 0.011 | 0.001 |
| Napping | rs3935190 | 17 | 79084367 | *BAIAP2* | A | G | 0.52 | 0.008 | 0.001 | 5.40E-11 | -0.027 | 0.009 | 0.004 |
| Napping | rs1941182 | 18 | 25569743 | *CDH2* | A | C | 0.22 | 0.007 | 0.001 | 4.10E-08 | 0.027 | 0.026 | 0.287 |
| Napping | rs2861805 | 18 | 36105007 | *CELF4* | A | G | 0.57 | 0.009 | 0.001 | 2.00E-14 | 0.006 | 0.010 | 0.561 |
| Napping | rs34728579 | 18 | 31610848 | *NOL4* | C | T | 0.20 | 0.008 | 0.002 | 3.70E-08 | -0.002 | 0.012 | 0.877 |
| Napping | rs9965170 | 18 | 44788274 | *SKOR2, SMAD2* | G | A | 0.60 | 0.014 | 0.001 | 7.80E-29 | 0.019 | 0.009 | 0.034 |
| Napping | rs17265513 | 20 | 39832628 | *ZHX3* | C | T | 0.19 | 0.009 | 0.002 | 2.00E-09 | 0.028 | 0.012 | 0.020 |
| Napping | rs3810484 | 20 | 62194103 | *HELZ2* | A | G | 0.57 | 0.007 | 0.001 | 2.20E-08 | 0.014 | 0.009 | 0.111 |
| Napping | rs910187 | 20 | 45841052 | *ZMYND8* | G | A | 0.85 | 0.007 | 0.001 | 4.90E-09 | 0.019 | 0.035 | 0.597 |
| Napping | rs1883048 | 21 | 47397586 | *COL6A1, PCBP3* | C | T | 0.50 | 0.008 | 0.001 | 1.80E-10 | -0.011 | 0.010 | 0.252 |
| Napping | rs2284015 | 22 | 37096573 | *CACNG2* | G | C | 0.27 | 0.008 | 0.001 | 4.10E-08 | -0.007 | 0.010 | 0.494 |
| Sleep duration | rs12567114 | 1 | 98527951 | *DPYD* | A | G | 0.28 | 0.015 | 0.003 | 4.30E-09 | -0.002 | 0.012 | 0.883 |
| Sleep duration | rs269054 | 1 | 57864304 | *DAB1* | A | T | 0.42 | 0.014 | 0.002 | 2.10E-09 | 0.022 | 0.009 | 0.016 |
| Sleep duration | rs61796569 | 1 | 66476437 | *PDE4B* | T | C | 0.27 | 0.015 | 0.003 | 1.50E-09 | -0.006 | 0.011 | 0.606 |
| Sleep duration | rs915416 | 1 | 34731984 | *CSMD2* | C | G | 0.29 | 0.019 | 0.002 | 9.90E-15 | -0.006 | 0.010 | 0.580 |
| Sleep duration | rs10173260 | 2 | 210377845 | *MAP2* | C | T | 0.61 | 0.013 | 0.002 | 2.90E-08 | -0.011 | 0.009 | 0.245 |
| Sleep duration | rs11885663 | 2 | 166944004 | *SCN1A* | T | C | 0.25 | 0.016 | 0.003 | 8.60E-10 | 0.022 | 0.011 | 0.038 |
| Sleep duration | rs374153 | 2 | 40382712 | *SLC8A1* | C | T | 0.16 | 0.018 | 0.003 | 9.10E-09 | 0.006 | 0.012 | 0.617 |
| Sleep duration | rs4128364 | 2 | 147612734 | *PABPCP2* | C | T | 0.34 | 0.015 | 0.002 | 1.40E-09 | -0.010 | 0.009 | 0.310 |
| Sleep duration | rs4538155 | 2 | 157040773 | *NR4A2* | T | C | 0.65 | 0.013 | 0.002 | 3.60E-08 | 0.010 | 0.010 | 0.284 |
| Sleep duration | rs62120041 | 2 | 9185564 | *MBOAT2* | T | C | 0.93 | 0.026 | 0.005 | 9.60E-09 | -0.015 | 0.020 | 0.471 |
| Sleep duration | rs72804080 | 2 | 59358659 | *LINC01122* | G | A | 0.15 | 0.018 | 0.003 | 2.90E-08 | -0.017 | 0.015 | 0.266 |
| Sleep duration | rs75539574 | 2 | 58871658 | *VRK2* | C | A | 0.09 | 0.036 | 0.004 | 6.90E-19 | 0.020 | 0.018 | 0.277 |
| Sleep duration | rs7556815 | 2 | 114085785 | *PAX8* | A | G | 0.22 | 0.041 | 0.003 | 1.00E-200 | -0.007 | 0.011 | 0.557 |
| Sleep duration | rs112230981 | 3 | 55879269 | *ERC2* | A | G | 0.95 | 0.032 | 0.005 | 2.20E-09 | -0.066 | 0.087 | 0.451 |
| Sleep duration | rs17732997 | 3 | 70470834 | *FOXP1* | C | G | 0.57 | 0.013 | 0.002 | 1.20E-08 | -0.009 | 0.010 | 0.341 |
| Sleep duration | rs7616632 | 3 | 137031237 | *IL20RB* | T | G | 0.52 | 0.013 | 0.002 | 4.30E-09 | 0.011 | 0.009 | 0.231 |
| Sleep duration | rs7644809 | 3 | 107564459 | *BBX* | T | C | 0.42 | 0.013 | 0.002 | 1.60E-08 | -0.005 | 0.010 | 0.604 |
| Sleep duration | rs13109404 | 4 | 102896591 | *BANK1* | T | G | 0.93 | 0.031 | 0.004 | 1.40E-12 | -0.060 | 0.019 | 0.001 |
| Sleep duration | rs17427571 | 4 | 82254908 | *PRKG2* | A | G | 0.68 | 0.014 | 0.002 | 1.30E-08 | -0.007 | 0.010 | 0.452 |
| Sleep duration | rs2192528 | 4 | 18327896 | *LCORL* | A | G | 0.48 | 0.013 | 0.002 | 2.70E-09 | -0.005 | 0.010 | 0.608 |
| Sleep duration | rs35531607 | 4 | 92533225 | *CCSER1* | C | T | 0.47 | 0.013 | 0.002 | 1.50E-08 | -0.024 | 0.009 | 0.008 |
| Sleep duration | rs11567976 | 5 | 137654218 | *CDC25C* | T | C | 0.57 | 0.013 | 0.002 | 2.10E-08 | -0.021 | 0.009 | 0.025 |
| Sleep duration | rs151014368 | 5 | 176751059 | *LMAN2* | A | G | 0.21 | 0.016 | 0.003 | 9.10E-09 | -0.021 | 0.012 | 0.086 |
| Sleep duration | rs365663 | 5 | 1428883 | *SLC6A3* | A | G | 0.55 | 0.015 | 0.002 | 1.00E-10 | -0.017 | 0.010 | 0.104 |
| Sleep duration | rs460692 | 5 | 3126584 | *LINC01377* | C | T | 0.14 | 0.021 | 0.003 | 3.60E-10 | 0.005 | 0.015 | 0.754 |
| Sleep duration | rs56372231 | 5 | 102321905 | *PAM* | T | C | 0.33 | 0.017 | 0.002 | 2.20E-12 | -0.012 | 0.010 | 0.208 |
| Sleep duration | rs113113059 | 6 | 43160375 | *CUL9* | T | C | 0.78 | 0.016 | 0.003 | 8.40E-09 | -0.027 | 0.011 | 0.010 |
| Sleep duration | rs2231265 | 6 | 89790201 | *PNRC1* | G | A | 0.77 | 0.015 | 0.003 | 2.70E-08 | -0.018 | 0.011 | 0.122 |
| Sleep duration | rs34556183 | 6 | 28584775 | *ZBED9* | A | G | 0.72 | 0.017 | 0.003 | 2.30E-11 | -0.018 | 0.010 | 0.069 |
| Sleep duration | rs80193650 | 6 | 33464363 | *ZBTB9* | G | A | 0.16 | 0.017 | 0.003 | 4.10E-08 | 0.007 | 0.013 | 0.608 |
| Sleep duration | rs9345234 | 6 | 93162639 | *LOC100129847* | C | A | 0.58 | 0.013 | 0.002 | 1.80E-08 | 0.009 | 0.009 | 0.311 |
| Sleep duration | rs9382445 | 6 | 54937974 | *FAM83B* | T | C | 0.62 | 0.015 | 0.002 | 4.80E-10 | -0.011 | 0.010 | 0.256 |
| Sleep duration | rs2079070 | 7 | 114126432 | *FOXP2* | C | G | 0.26 | 0.018 | 0.003 | 7.50E-12 | -0.022 | 0.011 | 0.047 |
| Sleep duration | rs34731055 | 7 | 2106928 | *MAD1L1* | T | C | 0.18 | 0.019 | 0.003 | 3.70E-11 | -0.024 | 0.012 | 0.052 |
| Sleep duration | rs7806045 | 7 | 132610266 | *CHCHD3* | T | C | 0.75 | 0.015 | 0.003 | 1.40E-08 | -0.017 | 0.010 | 0.100 |
| Sleep duration | rs330088 | 8 | 9149746 | *PPP1R3B* | C | T | 0.55 | 0.014 | 0.002 | 2.70E-10 | 0.019 | 0.010 | 0.056 |
| Sleep duration | rs73219758 | 8 | 14279446 | *SGCZ* | G | A | 0.71 | 0.016 | 0.002 | 5.60E-11 | 0.009 | 0.011 | 0.370 |
| Sleep duration | rs10973207 | 9 | 37100525 | *EBLN3* | T | G | 0.16 | 0.020 | 0.003 | 6.00E-11 | -0.001 | 0.013 | 0.963 |
| Sleep duration | rs1776776 | 9 | 140497072 | *ARRDC1* | T | C | 0.87 | 0.020 | 0.003 | 4.90E-09 | 0.006 | 0.012 | 0.634 |
| Sleep duration | rs10761674 | 10 | 64618340 | *EGR2* | C | T | 0.48 | 0.012 | 0.002 | 4.20E-08 | 0.015 | 0.009 | 0.109 |
| Sleep duration | rs11190970 | 10 | 103128332 | *BTRC* | G | A | 0.80 | 0.015 | 0.003 | 4.60E-08 | 0.016 | 0.012 | 0.175 |
| Sleep duration | rs12246842 | 10 | 21830580 | *MLLT10* | A | G | 0.46 | 0.013 | 0.002 | 3.90E-09 | -0.019 | 0.010 | 0.065 |
| Sleep duration | rs7915425 | 10 | 125016501 | *BUB3* | T | C | 0.17 | 0.019 | 0.003 | 2.00E-10 | -0.001 | 0.012 | 0.960 |
| Sleep duration | rs1057703 | 11 | 122830251 | *BSX* | G | T | 0.15 | 0.019 | 0.003 | 1.10E-09 | 0.001 | 0.012 | 0.966 |
| Sleep duration | rs11602180 | 11 | 48162453 | *PTPRJ* | C | T | 0.84 | 0.018 | 0.003 | 2.30E-09 | -0.021 | 0.012 | 0.077 |
| Sleep duration | rs1263056 | 11 | 116576415 | *BUD13* | A | G | 0.52 | 0.013 | 0.002 | 2.00E-08 | -0.031 | 0.009 | 0.001 |
| Sleep duration | rs12791153 | 11 | 80685181 | *LOC729790* | T | A | 0.08 | 0.024 | 0.004 | 1.90E-08 | -0.026 | 0.020 | 0.198 |
| Sleep duration | rs1517572 | 11 | 28829882 | *METT5D1* | C | A | 0.58 | 0.015 | 0.002 | 1.50E-10 | -0.013 | 0.010 | 0.187 |
| Sleep duration | rs1553132 | 11 | 88297740 | *GRM5* | G | A | 0.26 | 0.015 | 0.003 | 2.50E-08 | -0.017 | 0.010 | 0.100 |
| Sleep duration | rs174560 | 11 | 61581764 | *FADS1* | C | T | 0.31 | 0.014 | 0.002 | 2.80E-08 | -0.045 | 0.010 | 0.000 |
| Sleep duration | rs1939455 | 11 | 101520886 | *TRPC6* | G | T | 0.88 | 0.020 | 0.004 | 1.20E-08 | -0.003 | 0.014 | 0.856 |
| Sleep duration | rs4592416 | 11 | 43800474 | *HSD17B12* | G | A | 0.46 | 0.015 | 0.002 | 9.30E-11 | -0.010 | 0.009 | 0.273 |
| Sleep duration | rs7115226 | 11 | 113408518 | *DRD2* | A | C | 0.07 | 0.027 | 0.004 | 1.70E-09 | 0.023 | 0.016 | 0.144 |
| Sleep duration | rs7951019 | 11 | 118358027 | *KMT2A* | G | T | 0.03 | 0.037 | 0.007 | 1.20E-08 | -0.037 | 0.034 | 0.266 |
| Sleep duration | rs34354917 | 12 | 38764559 | *ALG10B* | C | A | 0.71 | 0.014 | 0.003 | 3.90E-08 | 0.008 | 0.012 | 0.499 |
| Sleep duration | rs4767550 | 12 | 117951150 | *KSR2* | G | A | 0.41 | 0.014 | 0.002 | 6.30E-10 | -0.004 | 0.010 | 0.670 |
| Sleep duration | rs10483350 | 14 | 29816155 | *MIR548AI* | G | A | 0.19 | 0.017 | 0.003 | 1.50E-09 | 0.003 | 0.012 | 0.798 |
| Sleep duration | rs11621908 | 14 | 78495761 | *ADCK1* | C | T | 0.92 | 0.024 | 0.004 | 5.60E-09 | 0.039 | 0.016 | 0.019 |
| Sleep duration | rs55658675 | 14 | 65554638 | *MAX* | C | T | 0.64 | 0.013 | 0.002 | 2.00E-08 | -0.007 | 0.010 | 0.516 |
| Sleep duration | rs61985058 | 14 | 60233841 | *RTN1* | T | C | 0.14 | 0.019 | 0.003 | 1.30E-08 | 0.003 | 0.014 | 0.824 |
| Sleep duration | rs6575005 | 14 | 26954078 | *NOVA1* | T | C | 0.76 | 0.016 | 0.003 | 4.40E-09 | 0.015 | 0.011 | 0.157 |
| Sleep duration | rs8038326 | 15 | 47989799 | *SEMA6D* | A | G | 0.73 | 0.016 | 0.003 | 2.80E-10 | 0.003 | 0.011 | 0.763 |
| Sleep duration | rs11643715 | 16 | 23909538 | *PRKCB* | G | C | 0.29 | 0.014 | 0.002 | 3.20E-08 | 0.013 | 0.011 | 0.215 |
| Sleep duration | rs3095508 | 16 | 6550400 | *RBFOX1* | C | A | 0.59 | 0.015 | 0.002 | 3.10E-11 | 0.004 | 0.009 | 0.681 |
| Sleep duration | rs8050478 | 16 | 56120461 | *GNAO1* | G | A | 0.50 | 0.016 | 0.002 | 1.70E-12 | 0.013 | 0.009 | 0.154 |
| Sleep duration | rs9940646 | 16 | 53800629 | *FTO* | C | G | 0.58 | 0.017 | 0.002 | 1.20E-13 | -0.051 | 0.009 | 0.000 |
| Sleep duration | rs1991556 | 17 | 44083402 | *MAPT* | G | A | 0.77 | 0.017 | 0.003 | 1.00E-09 | -0.038 | 0.012 | 0.001 |
| Sleep duration | rs205024 | 17 | 11227352 | *SHISA6* | T | C | 0.38 | 0.014 | 0.002 | 3.90E-09 | -0.004 | 0.009 | 0.656 |
| Sleep duration | rs7503199 | 17 | 8134275 | *PER1* | C | T | 0.73 | 0.015 | 0.003 | 1.00E-08 | -0.002 | 0.011 | 0.829 |
| Sleep duration | rs9903973 | 17 | 50571227 | *CA10* | C | T | 0.47 | 0.013 | 0.002 | 2.60E-08 | -0.001 | 0.010 | 0.939 |
| Sleep duration | rs12607679 | 18 | 53059748 | *TCF4* | T | C | 0.74 | 0.020 | 0.003 | 8.30E-15 | -0.006 | 0.011 | 0.610 |
| Sleep duration | rs10421649 | 19 | 9942262 | *FBXL12* | A | T | 0.56 | 0.013 | 0.002 | 6.90E-09 | 0.015 | 0.009 | 0.118 |
| Sleep duration | rs2072727 | 20 | 43538733 | *YWHAB* | T | C | 0.44 | 0.013 | 0.002 | 7.90E-09 | -0.031 | 0.009 | 0.001 |
| Sleep_long | rs7534398 | 1 | 7767464 | *CAMTA1* | A | T | 0.19 | 0.047 | 0.012 | 2.10E-08 | -0.005 | 0.012 | 0.678 |
| Sleep_long | rs6737318 | 2 | 114083120 | *PAX8* | G | A | 0.21 | 0.076 | 0.011 | 3.40E-13 | -0.007 | 0.012 | 0.563 |
| Sleep_long | rs10899257 | 11 | 76415209 | *GUCY2E* | A | G | 0.13 | 0.068 | 0.013 | 4.60E-08 | 0.000 | 0.013 | 0.999 |
| Sleep_long | rs3751046 | 11 | 122828342 | *BSX* | G | A | 0.17 | 0.070 | 0.014 | 2.00E-08 | 0.001 | 0.012 | 0.938 |
| Sleep_long | rs75458655 | 11 | 118115331 | *MPZL2* | T | C | 0.02 | 0.185 | 0.029 | 5.40E-12 | 0.086 | 0.046 | 0.060 |
| Sleep_long | rs17817288 | 16 | 53807764 | *FTO* | A | G | 0.53 | 0.039 | 0.009 | 8.90E-09 | -0.042 | 0.009 | 0.000 |
| Sleep_long | rs17688916 | 17 | 43778680 | *CRHR1* | T | A | 0.80 | 0.071 | 0.013 | 1.10E-11 | -0.039 | 0.012 | 0.001 |
| Sleep_short | rs12567114 | 1 | 98527951 | *DPYD* | G | A | 0.75 | 0.036 | 0.006 | 4.10E-09 | 0.002 | 0.012 | 0.883 |
| Sleep_short | rs2186122 | 1 | 66470206 | *PDE4B* | T | A | 0.57 | 0.024 | 0.006 | 4.80E-09 | -0.007 | 0.009 | 0.445 |
| Sleep_short | rs2820313 | 1 | 201870221 | *LMOD1* | G | A | 0.33 | 0.031 | 0.006 | 2.30E-09 | -0.022 | 0.010 | 0.024 |
| Sleep_short | rs7524118 | 1 | 34736052 | *CSMD2* | C | T | 0.67 | 0.030 | 0.006 | 4.90E-08 | 0.006 | 0.010 | 0.562 |
| Sleep_short | rs1380703 | 2 | 57941287 | *VRK2* | G | A | 0.36 | 0.035 | 0.005 | 1.60E-11 | 0.006 | 0.010 | 0.562 |
| Sleep_short | rs2863957 | 2 | 114089551 | *PAX8* | C | A | 0.79 | 0.054 | 0.007 | 2.60E-18 | 0.005 | 0.011 | 0.631 |
| Sleep_short | rs75539574 | 2 | 58871658 | *LINC01122* | A | C | 0.92 | 0.045 | 0.011 | 8.40E-11 | -0.020 | 0.018 | 0.277 |
| Sleep_short | rs2014830 | 3 | 50172397 | *RBM5* | C | T | 0.70 | 0.030 | 0.006 | 2.70E-08 | -0.009 | 0.010 | 0.393 |
| Sleep_short | rs13107325 | 4 | 103188709 | *SLC39A8* | T | C | 0.08 | 0.075 | 0.011 | 2.50E-13 | 0.067 | 0.017 | 0.000 |
| Sleep_short | rs17005118 | 4 | 82288564 | *RASGEF1B* | A | G | 0.28 | 0.030 | 0.006 | 2.50E-09 | 0.019 | 0.010 | 0.066 |
| Sleep_short | rs12518468 | 5 | 7249696 | *ADCY2* | C | T | 0.31 | 0.031 | 0.006 | 8.50E-09 | 0.001 | 0.010 | 0.911 |
| Sleep_short | rs3776864 | 5 | 102327868 | *PAM* | A | C | 0.70 | 0.031 | 0.006 | 1.70E-08 | 0.009 | 0.010 | 0.353 |
| Sleep_short | rs4585442 | 5 | 135508381 | *SMAD5* | G | A | 0.35 | 0.031 | 0.006 | 8.10E-10 | 0.009 | 0.010 | 0.386 |
| Sleep_short | rs12661667 | 6 | 41792545 | *USP49* | T | C | 0.24 | 0.028 | 0.006 | 2.80E-08 | -0.006 | 0.011 | 0.552 |
| Sleep_short | rs142180737 | 6 | 28344731 | *ZSCAN12* | C | T | 0.01 | 0.154 | 0.032 | 4.40E-09 | -0.012 | 0.056 | 0.836 |
| Sleep_short | rs9321171 | 6 | 129848635 | *LAMA2* | C | T | 0.56 | 0.031 | 0.006 | 4.20E-08 | 0.009 | 0.009 | 0.324 |
| Sleep_short | rs9367621 | 6 | 55040290 | *HCRTR2* | T | A | 0.41 | 0.024 | 0.006 | 1.60E-08 | 0.007 | 0.009 | 0.445 |
| Sleep_short | rs11763750 | 7 | 2080114 | *MAD1L1* | G | A | 0.81 | 0.035 | 0.008 | 5.10E-09 | 0.021 | 0.012 | 0.087 |
| Sleep_short | rs1229762 | 7 | 114218582 | *FOXP2* | T | C | 0.69 | 0.037 | 0.006 | 1.10E-12 | 0.020 | 0.010 | 0.043 |
| Sleep_short | rs60882754 | 8 | 52886619 | *PCMTD1* | A | T | 0.87 | 0.055 | 0.012 | 1.80E-08 | 0.002 | 0.016 | 0.917 |
| Sleep_short | rs1607227 | 11 | 28808617 | *METT5D1* | G | T | 0.72 | 0.031 | 0.006 | 1.50E-09 | 0.013 | 0.011 | 0.217 |
| Sleep_short | rs7939345 | 11 | 47980568 | *PTPRJ* | T | G | 0.21 | 0.035 | 0.007 | 4.00E-08 | 0.023 | 0.012 | 0.058 |
| Sleep_short | rs17388803 | 15 | 48027204 | *SEMA6D* | C | A | 0.12 | 0.053 | 0.010 | 6.50E-10 | -0.019 | 0.016 | 0.222 |
| Sleep_short | rs59779556 | 16 | 56227965 | *GNAO1* | T | G | 0.54 | 0.025 | 0.006 | 2.00E-08 | 0.007 | 0.009 | 0.462 |
| Sleep_short | rs205024 | 17 | 11227352 | *SHISA6* | C | T | 0.62 | 0.031 | 0.006 | 2.70E-08 | 0.004 | 0.009 | 0.656 |
| Sleep_short | rs12963463 | 18 | 53099093 | *TCF4* | C | T | 0.36 | 0.029 | 0.006 | 1.90E-11 | 0.014 | 0.010 | 0.174 |
| Sleep_short | rs5757675 | 22 | 39838892 | *MGAT3* | G | T | 0.31 | 0.034 | 0.007 | 2.70E-09 | 0.003 | 0.010 | 0.802 |
| Insomnia | rs10800992 | 1 | 190900576 | *NA* | T | C | NA | 0.042 | 0.006 | 3.84E-12 | 0.017 | 0.009 | 0.073 |
| Insomnia | rs11119409 | 1 | 210293333 | *-* | T | C | NA | -0.035 | 0.006 | 1.19E-08 | 0.034 | 0.010 | 0.001 |
| Insomnia | rs11588755 | 1 | 57819204 | *-* | A | G | NA | -0.035 | 0.006 | 5.14E-09 | 0.023 | 0.009 | 0.010 |
| Insomnia | rs11803128 | 1 | 190060095 | *-* | A | G | NA | -0.041 | 0.006 | 6.85E-11 | -0.007 | 0.010 | 0.471 |
| Insomnia | rs12030482 | 1 | 96961268 | *-* | A | T | NA | 0.041 | 0.007 | 8.16E-09 | 0.020 | 0.012 | 0.075 |
| Insomnia | rs1289939 | 1 | 117944435 | *-* | T | C | NA | -0.041 | 0.007 | 6.00E-09 | -0.023 | 0.011 | 0.033 |
| Insomnia | rs1620977 | 1 | 72729142 | *-* | A | G | NA | 0.052 | 0.007 | 2.27E-14 | -0.002 | 0.011 | 0.877 |
| Insomnia | rs2089358 | 1 | 37194103 | *-* | T | C | NA | -0.041 | 0.007 | 2.75E-10 | -0.006 | 0.010 | 0.582 |
| Insomnia | rs5877 | 1 | 173878862 | *-* | T | C | NA | 0.036 | 0.006 | 1.23E-08 | 0.011 | 0.010 | 0.252 |
| Insomnia | rs623025 | 1 | 201765094 | *-* | T | C | NA | -0.038 | 0.007 | 3.16E-08 | 0.010 | 0.010 | 0.344 |
| Insomnia | rs6702604 | 1 | 107190062 | *-* | A | G | NA | -0.037 | 0.006 | 1.30E-09 | -0.011 | 0.009 | 0.215 |
| Insomnia | rs699844 | 1 | 74878253 | *-* | A | G | NA | 0.060 | 0.011 | 4.11E-08 | 0.016 | 0.016 | 0.334 |
| Insomnia | rs10928256 | 2 | 146458738 | *-* | T | C | NA | 0.034 | 0.006 | 1.61E-08 | 0.015 | 0.010 | 0.147 |
| Insomnia | rs113851554 | 2 | 66750564 | *-* | T | G | NA | 0.206 | 0.014 | 1.00E-200 | 0.009 | 0.024 | 0.722 |
| Insomnia | rs116466468 | 2 | 159137557 | *-* | T | C | NA | 0.044 | 0.007 | 2.11E-10 | -0.023 | 0.012 | 0.047 |
| Insomnia | rs11679943 | 2 | 77724624 | *-* | A | G | NA | 0.037 | 0.006 | 3.16E-09 | 0.004 | 0.009 | 0.642 |
| Insomnia | rs12991815 | 2 | 68071990 | *-* | C | G | NA | 0.040 | 0.006 | 3.02E-11 | 0.004 | 0.010 | 0.656 |
| Insomnia | rs13010288 | 2 | 51824512 | *-* | T | G | NA | -0.060 | 0.009 | 9.26E-12 | -0.014 | 0.015 | 0.357 |
| Insomnia | rs1519102 | 2 | 66677816 | *-* | C | G | NA | -0.037 | 0.006 | 1.90E-08 | -0.009 | 0.011 | 0.402 |
| Insomnia | rs1530938 | 2 | 236900633 | *-* | A | G | NA | 0.036 | 0.006 | 8.82E-10 | 0.002 | 0.009 | 0.807 |
| Insomnia | rs1861412 | 2 | 58893065 | *-* | A | G | NA | 0.038 | 0.006 | 1.67E-10 | -0.020 | 0.009 | 0.024 |
| Insomnia | rs34967082 | 2 | 215382654 | *-* | A | G | NA | 0.035 | 0.006 | 4.34E-09 | -0.009 | 0.010 | 0.354 |
| Insomnia | rs4664299 | 2 | 160570033 | *-* | T | C | NA | -0.041 | 0.007 | 4.95E-09 | 0.001 | 0.010 | 0.928 |
| Insomnia | rs55772859 | 2 | 208042581 | *-* | A | C | NA | 0.042 | 0.006 | 4.82E-11 | -0.019 | 0.010 | 0.061 |
| Insomnia | rs56097173 | 2 | 44262449 | *-* | T | C | NA | 0.040 | 0.006 | 2.69E-10 | 0.015 | 0.010 | 0.109 |
| Insomnia | rs62158170 | 2 | 114082175 | *-* | A | G | NA | 0.066 | 0.007 | 1.20E-19 | 0.006 | 0.012 | 0.620 |
| Insomnia | rs62213452 | 2 | 210380152 | *-* | T | G | NA | 0.037 | 0.007 | 2.39E-08 | 0.015 | 0.011 | 0.175 |
| Insomnia | rs6545798 | 2 | 60521311 | *-* | A | T | NA | -0.041 | 0.006 | 1.19E-11 | 0.014 | 0.010 | 0.187 |
| Insomnia | rs6734957 | 2 | 42813247 | *-* | T | G | NA | -0.042 | 0.007 | 1.82E-09 | 0.013 | 0.013 | 0.308 |
| Insomnia | rs6756610 | 2 | 147480394 | *-* | C | G | NA | 0.037 | 0.006 | 1.14E-09 | 0.020 | 0.010 | 0.038 |
| Insomnia | rs72820274 | 2 | 104412924 | *-* | A | G | NA | 0.034 | 0.006 | 1.28E-08 | 0.015 | 0.010 | 0.130 |
| Insomnia | rs75452188 | 2 | 67134426 | *-* | A | G | NA | 0.052 | 0.009 | 1.58E-08 | 0.014 | 0.016 | 0.383 |
| Insomnia | rs7571486 | 2 | 176473295 | *-* | A | G | NA | -0.039 | 0.007 | 1.40E-08 | -0.016 | 0.010 | 0.130 |
| Insomnia | rs7599697 | 2 | 239231477 | *-* | T | C | NA | -0.037 | 0.006 | 5.00E-09 | 0.000 | 0.010 | 0.986 |
| Insomnia | rs823247 | 2 | 2850540 | *-* | T | C | NA | -0.037 | 0.006 | 5.25E-10 | -0.008 | 0.009 | 0.358 |
| Insomnia | rs10865954 | 3 | 49211989 | *-* | T | C | NA | 0.042 | 0.006 | 1.92E-11 | -0.014 | 0.009 | 0.147 |
| Insomnia | rs1567084 | 3 | 71435955 | *-* | A | G | NA | 0.033 | 0.006 | 2.14E-08 | 0.015 | 0.009 | 0.122 |
| Insomnia | rs1580173 | 3 | 107955515 | *-* | A | G | NA | 0.033 | 0.006 | 2.28E-08 | 0.014 | 0.009 | 0.113 |
| Insomnia | rs17025198 | 3 | 88001713 | *-* | A | G | NA | 0.041 | 0.007 | 2.19E-08 | -0.001 | 0.011 | 0.914 |
| Insomnia | rs2216427 | 3 | 180785697 | *-* | C | G | NA | 0.035 | 0.006 | 1.60E-08 | 0.009 | 0.010 | 0.361 |
| Insomnia | rs2364921 | 3 | 158522463 | *-* | T | C | NA | -0.034 | 0.006 | 2.13E-08 | 0.016 | 0.009 | 0.086 |
| Insomnia | rs35110063 | 3 | 43066558 | *-* | A | G | NA | 0.039 | 0.006 | 8.82E-11 | 0.002 | 0.009 | 0.830 |
| Insomnia | rs3774751 | 3 | 50209053 | *-* | T | G | NA | -0.041 | 0.006 | 7.32E-12 | 0.014 | 0.009 | 0.122 |
| Insomnia | rs4260410 | 3 | 178469932 | *-* | T | C | NA | 0.034 | 0.006 | 4.87E-08 | 0.010 | 0.010 | 0.287 |
| Insomnia | rs4858708 | 3 | 25154112 | *-* | A | T | NA | -0.034 | 0.006 | 1.23E-08 | 0.001 | 0.009 | 0.948 |
| Insomnia | rs492858 | 3 | 155432229 | *-* | T | C | NA | -0.066 | 0.011 | 3.46E-09 | 0.001 | 0.017 | 0.945 |
| Insomnia | rs62264767 | 3 | 117642005 | *-* | A | C | NA | 0.065 | 0.008 | 1.63E-14 | 0.006 | 0.013 | 0.633 |
| Insomnia | rs6808140 | 3 | 10581380 | *-* | T | C | NA | 0.039 | 0.006 | 5.35E-11 | -0.005 | 0.009 | 0.615 |
| Insomnia | rs694786 | 3 | 173112907 | *-* | T | C | NA | -0.044 | 0.006 | 1.97E-13 | -0.009 | 0.009 | 0.300 |
| Insomnia | rs7615602 | 3 | 18718055 | *-* | C | G | NA | -0.040 | 0.007 | 2.59E-09 | -0.045 | 0.011 | 0.000 |
| Insomnia | rs7625896 | 3 | 44062561 | *-* | A | G | NA | 0.036 | 0.006 | 5.28E-09 | 0.002 | 0.010 | 0.860 |
| Insomnia | rs11722569 | 4 | 112822731 | *-* | T | C | NA | 0.034 | 0.006 | 2.91E-08 | 0.008 | 0.010 | 0.427 |
| Insomnia | rs13135092 | 4 | 103198082 | *-* | A | G | NA | -0.089 | 0.011 | 2.53E-16 | -0.062 | 0.017 | 0.000 |
| Insomnia | rs13138995 | 4 | 148987430 | *-* | A | G | NA | 0.034 | 0.006 | 1.97E-08 | 0.005 | 0.010 | 0.606 |
| Insomnia | rs16990210 | 4 | 34720226 | *-* | T | C | NA | -0.046 | 0.008 | 1.97E-08 | 0.017 | 0.012 | 0.170 |
| Insomnia | rs17005118 | 4 | 82288564 | *-* | A | G | NA | 0.042 | 0.007 | 6.13E-10 | 0.019 | 0.010 | 0.066 |
| Insomnia | rs4699157 | 4 | 106055212 | *-* | T | C | NA | -0.081 | 0.015 | 3.98E-08 | -0.017 | 0.027 | 0.515 |
| Insomnia | rs62301574 | 4 | 22050165 | *-* | C | G | NA | -0.042 | 0.007 | 1.37E-08 | -0.001 | 0.011 | 0.964 |
| Insomnia | rs72657797 | 4 | 90820809 | *-* | T | C | NA | -0.056 | 0.008 | 1.52E-12 | -0.010 | 0.013 | 0.425 |
| Insomnia | rs12187443 | 5 | 102660400 | *-* | T | C | NA | 0.040 | 0.006 | 1.64E-10 | 0.009 | 0.010 | 0.395 |
| Insomnia | rs12520974 | 5 | 61514611 | *-* | T | C | NA | -0.036 | 0.006 | 1.69E-09 | 0.006 | 0.009 | 0.518 |
| Insomnia | rs152555 | 5 | 106849674 | *-* | A | G | NA | -0.052 | 0.008 | 4.83E-10 | -0.043 | 0.065 | 0.508 |
| Insomnia | rs16903122 | 5 | 87693561 | *-* | T | C | NA | 0.055 | 0.007 | 9.04E-16 | -0.006 | 0.011 | 0.587 |
| Insomnia | rs17083297 | 5 | 92995477 | *-* | A | C | NA | -0.044 | 0.008 | 1.60E-08 | 0.009 | 0.012 | 0.448 |
| Insomnia | rs17223714 | 5 | 50492629 | *-* | A | G | NA | 0.046 | 0.007 | 2.44E-10 | 0.000 | 0.012 | 0.994 |
| Insomnia | rs17367725 | 5 | 107112116 | *-* | T | C | NA | -0.036 | 0.006 | 9.29E-09 | 0.005 | 0.010 | 0.651 |
| Insomnia | rs2431108 | 5 | 103947968 | *-* | T | C | NA | -0.053 | 0.006 | 7.83E-17 | 0.013 | 0.010 | 0.214 |
| Insomnia | rs35539975 | 5 | 91607148 | *-* | A | G | NA | 0.042 | 0.007 | 4.49E-09 | 0.006 | 0.011 | 0.601 |
| Insomnia | rs4502882 | 5 | 153093998 | *-* | T | C | NA | -0.039 | 0.006 | 7.96E-10 | -0.004 | 0.010 | 0.727 |
| Insomnia | rs55972276 | 5 | 135653737 | *-* | A | C | NA | 0.073 | 0.009 | 4.19E-17 | -0.002 | 0.014 | 0.902 |
| Insomnia | rs62383308 | 5 | 165460085 | *-* | A | G | NA | -0.060 | 0.011 | 3.98E-08 | 0.029 | 0.020 | 0.140 |
| Insomnia | rs6601080 | 5 | 179511043 | *-* | A | G | NA | 0.035 | 0.006 | 2.21E-08 | 0.013 | 0.010 | 0.204 |
| Insomnia | rs6888135 | 5 | 141254063 | *-* | A | C | NA | 0.038 | 0.006 | 1.21E-10 | -0.012 | 0.010 | 0.245 |
| Insomnia | rs701394 | 5 | 80296487 | *-* | A | G | NA | -0.036 | 0.006 | 6.83E-09 | 0.003 | 0.011 | 0.801 |
| Insomnia | rs8180457 | 5 | 107209814 | *-* | T | C | NA | -0.056 | 0.008 | 1.12E-11 | 0.002 | 0.013 | 0.902 |
| Insomnia | rs10944696 | 6 | 94498850 | *-* | A | G | NA | -0.038 | 0.007 | 7.99E-09 | 0.007 | 0.011 | 0.540 |
| Insomnia | rs10947428 | 6 | 33647058 | *-* | T | C | NA | -0.068 | 0.007 | 9.06E-21 | -0.018 | 0.011 | 0.105 |
| Insomnia | rs10947690 | 6 | 37631768 | *-* | A | G | NA | -0.047 | 0.007 | 4.04E-12 | -0.014 | 0.011 | 0.197 |
| Insomnia | rs10947987 | 6 | 41754370 | *-* | T | C | NA | -0.033 | 0.006 | 4.08E-08 | -0.016 | 0.009 | 0.082 |
| Insomnia | rs1147852 | 6 | 147980909 | *-* | A | G | NA | 0.039 | 0.006 | 9.94E-10 | 0.005 | 0.010 | 0.583 |
| Insomnia | rs117152417 | 6 | 166411281 | *-* | A | G | NA | -0.147 | 0.026 | 2.82E-08 | 0.026 | 0.052 | 0.611 |
| Insomnia | rs11756035 | 6 | 18843810 | *-* | C | G | NA | 0.051 | 0.009 | 1.29E-08 | -0.005 | 0.017 | 0.785 |
| Insomnia | rs138678612 | 6 | 30932223 | *-* | A | G | NA | -0.117 | 0.020 | 1.41E-08 | 0.028 | 0.030 | 0.343 |
| Insomnia | rs2388840 | 6 | 99598756 | *-* | A | G | NA | -0.037 | 0.006 | 1.37E-09 | -0.009 | 0.009 | 0.311 |
| Insomnia | rs3131638 | 6 | 31475127 | *-* | A | G | NA | -0.044 | 0.007 | 7.88E-10 | -0.026 | 0.011 | 0.017 |
| Insomnia | rs314281 | 6 | 105400605 | *-* | T | C | NA | -0.043 | 0.006 | 6.03E-13 | 0.003 | 0.009 | 0.722 |
| Insomnia | rs4709655 | 6 | 163280204 | *-* | T | C | NA | -0.054 | 0.009 | 3.09E-09 | -0.022 | 0.015 | 0.141 |
| Insomnia | rs62429521 | 6 | 140324582 | *-* | A | C | NA | 0.051 | 0.008 | 1.78E-09 | 0.013 | 0.013 | 0.310 |
| Insomnia | rs6457796 | 6 | 34828553 | *-* | T | C | NA | -0.039 | 0.007 | 1.12E-08 | -0.038 | 0.010 | 0.000 |
| Insomnia | rs728017 | 6 | 124292594 | *-* | A | G | NA | -0.035 | 0.006 | 9.51E-09 | 0.009 | 0.009 | 0.348 |
| Insomnia | rs9373590 | 6 | 101212001 | *-* | A | T | NA | 0.040 | 0.006 | 2.18E-11 | 0.011 | 0.009 | 0.223 |
| Insomnia | rs9394502 | 6 | 38452503 | *-* | T | C | NA | -0.054 | 0.006 | 7.76E-18 | 0.004 | 0.010 | 0.650 |
| Insomnia | rs12666306 | 7 | 115082406 | *-* | A | G | NA | 0.042 | 0.006 | 2.24E-12 | 0.013 | 0.011 | 0.233 |
| Insomnia | rs1731951 | 7 | 137075847 | *-* | A | T | NA | -0.035 | 0.006 | 1.36E-08 | 0.007 | 0.011 | 0.538 |
| Insomnia | rs17520265 | 7 | 119674508 | *-* | A | G | NA | -0.091 | 0.016 | 2.87E-08 | 0.004 | 0.030 | 0.901 |
| Insomnia | rs190073 | 7 | 10985188 | *-* | A | G | NA | -0.034 | 0.006 | 2.86E-08 | -0.017 | 0.010 | 0.071 |
| Insomnia | rs2030672 | 7 | 21687925 | *-* | C | G | NA | 0.034 | 0.006 | 1.10E-08 | -0.005 | 0.009 | 0.568 |
| Insomnia | rs2598293 | 7 | 133989882 | *-* | T | C | NA | 0.035 | 0.006 | 2.48E-09 | 0.011 | 0.009 | 0.201 |
| Insomnia | rs521484 | 7 | 49894349 | *-* | A | G | NA | -0.040 | 0.007 | 1.53E-08 | -0.003 | 0.012 | 0.799 |
| Insomnia | rs6465151 | 7 | 88310899 | *-* | T | C | NA | 0.056 | 0.009 | 1.90E-09 | -0.031 | 0.016 | 0.049 |
| Insomnia | rs670501 | 7 | 108625185 | *-* | T | C | NA | 0.053 | 0.007 | 7.40E-13 | 0.005 | 0.011 | 0.651 |
| Insomnia | rs6967168 | 7 | 132672192 | *-* | T | G | NA | -0.044 | 0.007 | 1.39E-10 | -0.014 | 0.011 | 0.187 |
| Insomnia | rs6978112 | 7 | 1966841 | *-* | T | C | NA | 0.034 | 0.006 | 2.11E-08 | 0.016 | 0.010 | 0.095 |
| Insomnia | rs73671843 | 7 | 3520024 | *-* | A | G | NA | -0.056 | 0.009 | 5.49E-10 | -0.022 | 0.013 | 0.089 |
| Insomnia | rs75932578 | 7 | 106844694 | *-* | T | C | NA | -0.040 | 0.007 | 4.15E-08 | 0.016 | 0.012 | 0.179 |
| Insomnia | rs8180817 | 7 | 114047542 | *-* | C | G | NA | -0.049 | 0.006 | 1.83E-16 | -0.012 | 0.009 | 0.193 |
| Insomnia | rs10955647 | 8 | 114154187 | *-* | T | G | NA | 0.033 | 0.006 | 1.84E-08 | 0.012 | 0.009 | 0.208 |
| Insomnia | rs17643634 | 8 | 91650818 | *-* | T | C | NA | -0.060 | 0.008 | 1.34E-13 | -0.009 | 0.014 | 0.525 |
| Insomnia | rs2737240 | 8 | 116657235 | *-* | A | G | NA | 0.036 | 0.007 | 3.37E-08 | 0.005 | 0.010 | 0.615 |
| Insomnia | rs28552587 | 8 | 103356226 | *-* | A | G | NA | 0.033 | 0.006 | 3.30E-08 | 0.008 | 0.010 | 0.394 |
| Insomnia | rs28611339 | 8 | 10170037 | *-* | T | G | NA | 0.058 | 0.009 | 8.46E-11 | -0.033 | 0.013 | 0.011 |
| Insomnia | rs4588900 | 8 | 73890425 | *-* | A | G | NA | 0.033 | 0.006 | 1.57E-08 | 0.011 | 0.009 | 0.235 |
| Insomnia | rs671985 | 8 | 60914783 | *-* | A | G | NA | -0.038 | 0.006 | 2.79E-10 | 0.000 | 0.009 | 0.991 |
| Insomnia | rs871994 | 8 | 35190619 | *-* | A | C | NA | 0.035 | 0.006 | 5.50E-09 | 0.001 | 0.009 | 0.926 |
| Insomnia | rs874168 | 8 | 30849450 | *-* | T | C | NA | 0.034 | 0.006 | 7.95E-09 | 0.026 | 0.010 | 0.009 |
| Insomnia | rs10756571 | 9 | 14534505 | *-* | T | C | NA | 0.036 | 0.006 | 1.80E-08 | 0.009 | 0.011 | 0.406 |
| Insomnia | rs10758593 | 9 | 4292083 | *-* | A | G | NA | -0.036 | 0.006 | 4.90E-09 | -0.007 | 0.009 | 0.430 |
| Insomnia | rs10761240 | 9 | 96361922 | *-* | A | G | NA | -0.043 | 0.006 | 2.12E-12 | 0.007 | 0.009 | 0.459 |
| Insomnia | rs118166957 | 9 | 8858043 | *-* | T | C | NA | 0.068 | 0.008 | 1.95E-16 | -0.011 | 0.013 | 0.417 |
| Insomnia | rs1927902 | 9 | 120518991 | *-* | T | C | NA | 0.053 | 0.007 | 1.15E-14 | 0.026 | 0.010 | 0.010 |
| Insomnia | rs2792990 | 9 | 125621610 | *-* | C | G | NA | 0.054 | 0.008 | 1.15E-10 | 0.015 | 0.012 | 0.215 |
| Insomnia | rs4090240 | 9 | 77118987 | *-* | T | C | NA | -0.039 | 0.007 | 8.46E-09 | -0.002 | 0.011 | 0.883 |
| Insomnia | rs6597649 | 9 | 133786652 | *-* | T | C | NA | 0.033 | 0.006 | 3.05E-08 | 0.010 | 0.009 | 0.286 |
| Insomnia | rs7040224 | 9 | 134886837 | *-* | A | G | NA | 0.037 | 0.006 | 4.24E-09 | 0.008 | 0.009 | 0.402 |
| Insomnia | rs7044885 | 9 | 81739348 | *-* | C | G | NA | -0.041 | 0.006 | 5.67E-12 | 0.009 | 0.010 | 0.367 |
| Insomnia | rs72773790 | 9 | 139109080 | *-* | T | C | NA | 0.037 | 0.006 | 3.71E-09 | -0.004 | 0.010 | 0.670 |
| Insomnia | rs77641763 | 9 | 140265782 | *-* | T | C | NA | 0.071 | 0.009 | 6.53E-15 | -0.017 | 0.016 | 0.289 |
| Insomnia | rs10825503 | 10 | 57177470 | *-* | T | G | NA | 0.033 | 0.006 | 1.43E-08 | 0.010 | 0.009 | 0.256 |
| Insomnia | rs11001276 | 10 | 76825638 | *-* | A | T | NA | -0.038 | 0.007 | 2.52E-08 | -0.033 | 0.010 | 0.001 |
| Insomnia | rs12251016 | 10 | 21821918 | *-* | A | T | NA | -0.039 | 0.006 | 3.89E-10 | -0.012 | 0.009 | 0.191 |
| Insomnia | rs224029 | 10 | 64519299 | *-* | T | C | NA | -0.039 | 0.006 | 2.51E-10 | 0.011 | 0.009 | 0.236 |
| Insomnia | rs7475916 | 10 | 77771194 | *-* | C | G | NA | -0.037 | 0.006 | 6.70E-09 | 0.016 | 0.012 | 0.168 |
| Insomnia | rs1064939 | 11 | 118396331 | *-* | A | T | NA | 0.130 | 0.020 | 2.16E-10 | 0.016 | 0.041 | 0.702 |
| Insomnia | rs10898940 | 11 | 73455292 | *-* | A | C | NA | 0.034 | 0.006 | 8.09E-09 | 0.090 | 0.048 | 0.057 |
| Insomnia | rs11605348 | 11 | 47606483 | *-* | A | G | NA | -0.045 | 0.006 | 7.01E-13 | -0.008 | 0.010 | 0.436 |
| Insomnia | rs12790660 | 11 | 57667222 | *-* | T | C | NA | -0.040 | 0.006 | 4.49E-10 | 0.010 | 0.011 | 0.357 |
| Insomnia | rs214934 | 11 | 17193475 | *-* | A | T | NA | -0.038 | 0.006 | 3.16E-09 | 0.016 | 0.010 | 0.127 |
| Insomnia | rs2221119 | 11 | 88598444 | *-* | C | G | NA | 0.036 | 0.006 | 2.00E-09 | 0.016 | 0.009 | 0.090 |
| Insomnia | rs4592425 | 11 | 62697813 | *-* | T | G | NA | 0.040 | 0.006 | 4.31E-10 | 0.007 | 0.011 | 0.511 |
| Insomnia | rs524859 | 11 | 66041079 | *-* | A | G | NA | -0.044 | 0.006 | 1.48E-12 | -0.007 | 0.010 | 0.489 |
| Insomnia | rs56133505 | 11 | 72348039 | *-* | A | G | NA | 0.041 | 0.006 | 5.59E-12 | -0.010 | 0.009 | 0.311 |
| Insomnia | rs566673 | 11 | 66401373 | *-* | T | G | NA | -0.039 | 0.006 | 1.18E-10 | -0.005 | 0.010 | 0.649 |
| Insomnia | rs647905 | 11 | 121534938 | *-* | T | C | NA | 0.033 | 0.006 | 2.87E-08 | -0.013 | 0.009 | 0.151 |
| Insomnia | rs6589988 | 11 | 99126016 | *-* | A | G | NA | -0.038 | 0.006 | 4.70E-09 | 0.002 | 0.010 | 0.838 |
| Insomnia | rs667730 | 11 | 83277325 | *-* | T | C | NA | 0.033 | 0.006 | 2.26E-08 | 0.003 | 0.009 | 0.753 |
| Insomnia | rs72899452 | 11 | 45415577 | *-* | T | C | NA | 0.074 | 0.012 | 1.00E-09 | 0.002 | 0.018 | 0.905 |
| Insomnia | rs1167132 | 12 | 43484487 | *-* | T | C | NA | 0.035 | 0.006 | 8.73E-09 | 0.005 | 0.009 | 0.553 |
| Insomnia | rs12310246 | 12 | 84700945 | *-* | A | G | NA | 0.045 | 0.007 | 4.74E-11 | 0.010 | 0.010 | 0.316 |
| Insomnia | rs2286729 | 12 | 6873818 | *-* | A | G | NA | 0.070 | 0.011 | 5.37E-11 | 0.018 | 0.018 | 0.296 |
| Insomnia | rs28582096 | 12 | 123856998 | *-* | A | G | NA | -0.054 | 0.007 | 1.74E-13 | 0.000 | 0.011 | 0.990 |
| Insomnia | rs324017 | 12 | 57487814 | *-* | A | C | NA | 0.039 | 0.007 | 1.61E-09 | 0.007 | 0.010 | 0.504 |
| Insomnia | rs4767645 | 12 | 118385788 | *-* | T | G | NA | -0.037 | 0.006 | 6.47E-10 | -0.007 | 0.009 | 0.442 |
| Insomnia | rs61921611 | 12 | 66367726 | *-* | T | C | NA | -0.044 | 0.006 | 7.84E-12 | -0.021 | 0.010 | 0.033 |
| Insomnia | rs6606731 | 12 | 109982578 | *-* | A | T | NA | 0.043 | 0.008 | 1.51E-08 | 0.022 | 0.011 | 0.046 |
| Insomnia | rs1031654 | 13 | 54382035 | *-* | A | C | NA | -0.051 | 0.007 | 3.88E-12 | 0.008 | 0.011 | 0.451 |
| Insomnia | rs11149313 | 13 | 85294881 | *-* | A | G | NA | 0.040 | 0.007 | 2.38E-09 | -0.010 | 0.010 | 0.341 |
| Insomnia | rs1536053 | 13 | 111982291 | *-* | T | C | NA | -0.038 | 0.006 | 6.04E-09 | -0.024 | 0.010 | 0.015 |
| Insomnia | rs2389631 | 13 | 96932868 | *-* | A | C | NA | -0.040 | 0.006 | 2.03E-10 | -0.018 | 0.009 | 0.059 |
| Insomnia | rs6562066 | 13 | 60532796 | *-* | T | C | NA | 0.039 | 0.006 | 1.38E-10 | -0.018 | 0.010 | 0.068 |
| Insomnia | rs7992992 | 13 | 54721699 | *-* | A | G | NA | 0.051 | 0.009 | 1.15E-08 | 0.026 | 0.012 | 0.032 |
| Insomnia | rs9527083 | 13 | 53991125 | *-* | A | G | NA | -0.076 | 0.006 | 1.61E-32 | 0.007 | 0.010 | 0.496 |
| Insomnia | rs9540729 | 13 | 66947124 | *-* | A | T | NA | 0.036 | 0.006 | 1.40E-09 | 0.014 | 0.009 | 0.141 |
| Insomnia | rs9563886 | 13 | 61720066 | *-* | T | C | NA | -0.034 | 0.006 | 3.08E-08 | -0.002 | 0.009 | 0.825 |
| Insomnia | rs4981170 | 14 | 33412996 | *-* | A | G | NA | -0.054 | 0.008 | 7.33E-13 | 0.018 | 0.012 | 0.127 |
| Insomnia | rs1038093 | 15 | 74012409 | *-* | T | C | NA | 0.039 | 0.006 | 2.47E-10 | 0.007 | 0.010 | 0.477 |
| Insomnia | rs12912299 | 15 | 38897857 | *-* | T | C | NA | -0.043 | 0.006 | 4.42E-13 | -0.009 | 0.009 | 0.332 |
| Insomnia | rs12917449 | 15 | 74331659 | *-* | A | C | NA | -0.042 | 0.008 | 2.97E-08 | -0.030 | 0.012 | 0.011 |
| Insomnia | rs176644 | 15 | 89913632 | *-* | T | G | NA | 0.035 | 0.006 | 9.49E-09 | 0.016 | 0.010 | 0.117 |
| Insomnia | rs4702 | 15 | 91426560 | *-* | A | G | NA | -0.048 | 0.006 | 6.78E-16 | 0.002 | 0.009 | 0.835 |
| Insomnia | rs715338 | 15 | 57215867 | *-* | A | G | NA | 0.041 | 0.006 | 7.85E-12 | -0.018 | 0.028 | 0.518 |
| Insomnia | rs7168238 | 15 | 66709386 | *-* | C | G | NA | 0.064 | 0.011 | 1.80E-08 | 0.054 | 0.018 | 0.003 |
| Insomnia | rs7402939 | 15 | 99183876 | *-* | T | C | NA | -0.036 | 0.006 | 5.19E-09 | -0.020 | 0.010 | 0.052 |
| Insomnia | rs1015438 | 16 | 51177517 | *-* | A | G | NA | 0.058 | 0.008 | 2.51E-14 | 0.013 | 0.011 | 0.256 |
| Insomnia | rs12924275 | 16 | 9191790 | *-* | T | C | NA | 0.038 | 0.007 | 1.93E-08 | 0.005 | 0.010 | 0.651 |
| Insomnia | rs3184470 | 16 | 715164 | *-* | A | G | NA | -0.038 | 0.006 | 9.73E-10 | -0.009 | 0.010 | 0.363 |
| Insomnia | rs34214423 | 16 | 52303107 | *-* | A | C | NA | 0.045 | 0.008 | 3.18E-09 | 0.009 | 0.012 | 0.451 |
| Insomnia | rs35322724 | 16 | 77137324 | *-* | A | C | NA | 0.049 | 0.006 | 3.75E-16 | 0.008 | 0.009 | 0.404 |
| Insomnia | rs3902952 | 16 | 61647589 | *-* | T | C | NA | 0.048 | 0.008 | 2.55E-10 | -0.011 | 0.012 | 0.353 |
| Insomnia | rs4238755 | 16 | 52746089 | *-* | A | C | NA | -0.043 | 0.007 | 2.30E-10 | 0.016 | 0.011 | 0.130 |
| Insomnia | rs4788203 | 16 | 29978827 | *-* | A | G | NA | -0.035 | 0.006 | 6.32E-09 | 0.018 | 0.010 | 0.065 |
| Insomnia | rs66674044 | 16 | 19904344 | *-* | A | T | NA | -0.060 | 0.009 | 2.18E-12 | 0.011 | 0.013 | 0.401 |
| Insomnia | rs830716 | 16 | 12323509 | *-* | C | G | NA | 0.045 | 0.007 | 8.68E-12 | -0.003 | 0.011 | 0.771 |
| Insomnia | rs9931543 | 16 | 56128782 | *-* | T | C | NA | 0.048 | 0.007 | 1.11E-12 | -0.002 | 0.011 | 0.869 |
| Insomnia | rs11650304 | 17 | 46035001 | *-* | C | G | NA | 0.067 | 0.012 | 1.23E-08 | 0.031 | 0.019 | 0.106 |
| Insomnia | rs34490907 | 17 | 26933741 | *-* | C | G | NA | 0.054 | 0.009 | 1.76E-08 | 0.010 | 0.017 | 0.549 |
| Insomnia | rs4643373 | 17 | 47123423 | *-* | T | C | NA | 0.041 | 0.007 | 1.58E-10 | 0.037 | 0.010 | 0.000 |
| Insomnia | rs62068188 | 17 | 2400876 | *-* | T | C | NA | 0.049 | 0.008 | 1.18E-09 | -0.015 | 0.049 | 0.761 |
| Insomnia | rs7214267 | 17 | 43157709 | *-* | A | G | NA | -0.044 | 0.006 | 5.09E-13 | -0.003 | 0.009 | 0.770 |
| Insomnia | rs8076183 | 17 | 61024696 | *-* | T | C | NA | -0.038 | 0.006 | 2.75E-10 | 0.016 | 0.009 | 0.069 |
| Insomnia | rs9889282 | 17 | 50259142 | *-* | A | C | NA | -0.042 | 0.006 | 4.70E-12 | -0.009 | 0.010 | 0.387 |
| Insomnia | rs10502966 | 18 | 50748499 | *-* | A | G | NA | -0.039 | 0.006 | 8.54E-11 | -0.016 | 0.010 | 0.091 |
| Insomnia | rs12454003 | 18 | 26315799 | *-* | C | G | NA | -0.035 | 0.006 | 4.94E-09 | 0.004 | 0.010 | 0.705 |
| Insomnia | rs12605642 | 18 | 31313965 | *-* | T | G | NA | 0.035 | 0.006 | 2.13E-09 | -0.011 | 0.009 | 0.228 |
| Insomnia | rs60565673 | 18 | 52906830 | *-* | T | G | NA | -0.043 | 0.006 | 1.59E-12 | -0.012 | 0.009 | 0.194 |
| Insomnia | rs9964420 | 18 | 56824041 | *-* | A | C | NA | 0.035 | 0.007 | 4.54E-08 | -0.011 | 0.010 | 0.276 |
| Insomnia | rs12983032 | 19 | 5073447 | *-* | A | G | NA | -0.043 | 0.006 | 1.07E-11 | -0.025 | 0.010 | 0.013 |
| Insomnia | rs429358 | 19 | 45411941 | *-* | T | C | NA | 0.046 | 0.008 | 2.13E-08 | -0.036 | 0.013 | 0.003 |
| Insomnia | rs6510033 | 19 | 30710785 | *-* | A | G | NA | -0.037 | 0.007 | 4.66E-08 | -0.020 | 0.012 | 0.099 |
| Insomnia | rs908668 | 19 | 56134038 | *-* | T | C | NA | 0.050 | 0.007 | 1.41E-11 | 0.008 | 0.011 | 0.489 |
| Insomnia | rs2867690 | 20 | 41972028 | *-* | T | C | NA | 0.042 | 0.008 | 3.70E-08 | 0.020 | 0.013 | 0.117 |
| Insomnia | rs6019663 | 20 | 47774512 | *-* | T | C | NA | 0.040 | 0.007 | 6.47E-10 | -0.001 | 0.010 | 0.903 |
| Insomnia | rs6119267 | 20 | 31163914 | *-* | C | G | NA | -0.060 | 0.006 | 2.32E-20 | -0.007 | 0.010 | 0.476 |
| Insomnia | rs742760 | 20 | 50985290 | *-* | A | T | NA | 0.043 | 0.008 | 2.48E-08 | -0.013 | 0.013 | 0.315 |
| Insomnia | rs76145129 | 20 | 62670427 | *-* | T | G | NA | -0.050 | 0.009 | 2.73E-08 | -0.023 | 0.016 | 0.150 |
| Insomnia | rs910187 | 20 | 45841052 | *-* | A | G | NA | -0.035 | 0.006 | 1.63E-08 | -0.019 | 0.035 | 0.597 |
| Insomnia | rs2838787 | 21 | 46539725 | *-* | A | G | NA | -0.036 | 0.006 | 7.65E-09 | -0.007 | 0.011 | 0.501 |
| Insomnia | rs11090039 | 22 | 41496800 | *-* | A | G | NA | 0.039 | 0.007 | 1.82E-09 | 0.018 | 0.011 | 0.118 |
| Insomnia | rs9931543 | 16 | 56128782 | *-* | T | C | NA | 0.048 | 0.007 | 1.11E-12 | -0.002 | 0.011 | 0.869 |
| Insomnia | rs11650304 | 17 | 46035001 | *-* | C | G | NA | 0.067 | 0.012 | 1.23E-08 | 0.031 | 0.019 | 0.106 |
| Insomnia | rs34490907 | 17 | 26933741 | *-* | C | G | NA | 0.054 | 0.009 | 1.76E-08 | 0.010 | 0.017 | 0.549 |
| Insomnia | rs4643373 | 17 | 47123423 | *-* | T | C | NA | 0.041 | 0.007 | 1.58E-10 | 0.037 | 0.010 | 0.000 |
| Insomnia | rs62068188 | 17 | 2400876 | *-* | T | C | NA | 0.049 | 0.008 | 1.18E-09 | -0.015 | 0.049 | 0.761 |
| Insomnia | rs7214267 | 17 | 43157709 | *-* | A | G | NA | -0.044 | 0.006 | 5.09E-13 | -0.003 | 0.009 | 0.770 |
| Insomnia | rs8076183 | 17 | 61024696 | *-* | T | C | NA | -0.038 | 0.006 | 2.75E-10 | 0.016 | 0.009 | 0.069 |
| Insomnia | rs9889282 | 17 | 50259142 | *-* | A | C | NA | -0.042 | 0.006 | 4.70E-12 | -0.009 | 0.010 | 0.387 |
| Insomnia | rs10502966 | 18 | 50748499 | *-* | A | G | NA | -0.039 | 0.006 | 8.54E-11 | -0.016 | 0.010 | 0.091 |
| Insomnia | rs12454003 | 18 | 26315799 | *-* | C | G | NA | -0.035 | 0.006 | 4.94E-09 | 0.004 | 0.010 | 0.705 |
| Insomnia | rs12605642 | 18 | 31313965 | *-* | T | G | NA | 0.035 | 0.006 | 2.13E-09 | -0.011 | 0.009 | 0.228 |
| Insomnia | rs60565673 | 18 | 52906830 | *-* | T | G | NA | -0.043 | 0.006 | 1.59E-12 | -0.012 | 0.009 | 0.194 |
| Insomnia | rs9964420 | 18 | 56824041 | *-* | A | C | NA | 0.035 | 0.007 | 4.54E-08 | -0.011 | 0.010 | 0.276 |
| Insomnia | rs12983032 | 19 | 5073447 | *-* | A | G | NA | -0.043 | 0.006 | 1.07E-11 | -0.025 | 0.010 | 0.013 |
| Insomnia | rs429358 | 19 | 45411941 | *-* | T | C | NA | 0.046 | 0.008 | 2.13E-08 | -0.036 | 0.013 | 0.003 |
| Insomnia | rs6510033 | 19 | 30710785 | *-* | A | G | NA | -0.037 | 0.007 | 4.66E-08 | -0.020 | 0.012 | 0.099 |
| Insomnia | rs908668 | 19 | 56134038 | *-* | T | C | NA | 0.050 | 0.007 | 1.41E-11 | 0.008 | 0.011 | 0.489 |
| Insomnia | rs2867690 | 20 | 41972028 | *-* | T | C | NA | 0.042 | 0.008 | 3.70E-08 | 0.020 | 0.013 | 0.117 |
| Insomnia | rs6019663 | 20 | 47774512 | *-* | T | C | NA | 0.040 | 0.007 | 6.47E-10 | -0.001 | 0.010 | 0.903 |
| Insomnia | rs6119267 | 20 | 31163914 | *-* | C | G | NA | -0.060 | 0.006 | 2.32E-20 | -0.007 | 0.010 | 0.476 |
| Insomnia | rs742760 | 20 | 50985290 | *-* | A | T | NA | 0.043 | 0.008 | 2.48E-08 | -0.013 | 0.013 | 0.315 |
| Insomnia | rs76145129 | 20 | 62670427 | *-* | T | G | NA | -0.050 | 0.009 | 2.73E-08 | -0.023 | 0.016 | 0.150 |
| Insomnia | rs910187 | 20 | 45841052 | *-* | A | G | NA | -0.035 | 0.006 | 1.63E-08 | -0.019 | 0.035 | 0.597 |
| Insomnia | rs2838787 | 21 | 46539725 | *-* | A | G | NA | -0.036 | 0.006 | 7.65E-09 | -0.007 | 0.011 | 0.501 |
| Insomnia | rs11090039 | 22 | 41496800 | *-* | A | G | NA | 0.039 | 0.007 | 1.82E-09 | 0.018 | 0.011 | 0.118 |
| Apnea | rs11205802 | 1 | 39699114 | *MACF1* | T | C | 0.61 | -0.020 | 0.004 | 7.30E-10 | 0.007 | 0.010 | 0.504 |
| Apnea | rs543874 | 1 | 177889480 | *FAM5B,SEC16B* | G | A | 0.21 | -0.027 | 0.005 | 1.00E-15 | 0.012 | 0.011 | 0.257 |
| Apnea | rs13021737 | 2 | 632348 | *FAM150B,TMEM18* | G | A | 0.84 | -0.022 | 0.005 | 7.88E-22 | 0.053 | 0.012 | 0.000 |
| Apnea | rs72904209 | 2 | 157046432 | *NR4A2* | T | C | 0.88 | -0.036 | 0.006 | 1.31E-14 | 0.009 | 0.015 | 0.553 |
| Apnea | rs1403848 | 3 | 77609655 | *ROBO2* | C | A | 0.41 | 0.016 | 0.004 | 2.27E-10 | 0.012 | 0.009 | 0.208 |
| Apnea | rs1554654 | 3 | 44044344 | *ABHD5,TOPAZ1* | T | C | 0.51 | -0.018 | 0.004 | 8.12E-10 | -0.018 | 0.009 | 0.049 |
| Apnea | rs10075809 | 5 | 122703026 | *CEP120* | C | A | 0.30 | 0.016 | 0.004 | 2.91E-11 | 0.015 | 0.010 | 0.111 |
| Apnea | rs7005777 | 8 | 78233600 | *PEX2* | T | G | 0.74 | 0.018 | 0.004 | 3.68E-08 | -0.014 | 0.011 | 0.201 |
| Apnea | rs79932406 | 8 | 71607667 | *XKR9* | T | G | 0.39 | 0.016 | 0.004 | 2.21E-08 | -0.011 | 0.009 | 0.244 |
| Apnea | rs12683343 | 9 | 128012939 | *HSPA5,GAPVD1* | G | A | 0.51 | 0.015 | 0.004 | 1.62E-08 | -0.007 | 0.010 | 0.485 |
| Apnea | rs8176749 | 9 | 136131188 | *ABO* | T | C | 0.10 | -0.032 | 0.007 | 1.23E-09 | 0.051 | 0.015 | 0.001 |
| Apnea | rs1444789 | 10 | 9064361 | *GATA3* | T | C | 0.79 | -0.037 | 0.005 | 2.40E-13 | -0.019 | 0.011 | 0.082 |
| Apnea | rs11821161 | 11 | 88888262 | *GRM5,TYR* | T | C | 0.58 | -0.015 | 0.004 | 1.15E-08 | -0.020 | 0.009 | 0.027 |
| Apnea | rs12805133 | 11 | 66483265 | *SPTBN2* | G | A | 0.44 | 0.019 | 0.004 | 1.44E-09 | 0.011 | 0.009 | 0.238 |
| Apnea | rs6265 | 11 | 27679916 | *BDNF* | T | C | 0.18 | -0.022 | 0.005 | 1.29E-36 | -0.034 | 0.012 | 0.005 |
| Apnea | rs7107532 | 11 | 28480924 | *METTL15* | G | A | 0.48 | -0.027 | 0.004 | 2.27E-12 | -0.006 | 0.009 | 0.537 |
| Apnea | rs2277339 | 12 | 57146069 | *PRIM1* | T | G | 0.87 | 0.045 | 0.006 | 5.19E-14 | -0.013 | 0.014 | 0.323 |
| Apnea | rs2958153 | 12 | 57081517 | *PTGES3* | G | A | 0.74 | -0.031 | 0.004 | 1.29E-12 | 0.014 | 0.011 | 0.198 |
| Apnea | rs7138383 | 12 | 103724090 | *C12orf42* | G | A | 0.69 | 0.017 | 0.004 | 4.67E-08 | 0.014 | 0.010 | 0.168 |
| Apnea | rs9783497 | 12 | 65830349 | *MSRB3* | G | A | 0.67 | -0.040 | 0.004 | 1.23E-25 | -0.008 | 0.010 | 0.443 |
| Apnea | rs9526702 | 13 | 51475452 | *DLEU7,RNASEH2B* | G | A | 0.88 | 0.024 | 0.006 | 4.55E-08 | -0.001 | 0.015 | 0.924 |
| Apnea | rs11634019 | 15 | 76634680 | *ISL2* | T | C | 0.73 | 0.027 | 0.004 | 4.44E-10 | 0.007 | 0.011 | 0.530 |
| Apnea | rs11075985 | 16 | 53805207 | *FTO* | C | A | 0.57 | -0.017 | 0.004 | 1.68E-23 | -0.051 | 0.009 | 0.000 |
| Apnea | rs1136070 | 16 | 1751935 | *HN1L* | T | C | 0.16 | 0.037 | 0.007 | 4.06E-09 | 0.025 | 0.015 | 0.091 |
| Apnea | rs1436047 | 16 | 60618247 | *RP11-457D20.2* | G | A | 0.40 | -0.025 | 0.004 | 1.19E-10 | -0.001 | 0.009 | 0.948 |
| Apnea | rs879620 | 16 | 4015729 | *ADCY9* | T | C | 0.55 | -0.016 | 0.004 | 1.04E-09 | 0.025 | 0.010 | 0.010 |
| Apnea | rs12603115 | 17 | 46248994 | *SKAP1* | T | C | 0.53 | -0.018 | 0.004 | 1.23E-10 | -0.013 | 0.009 | 0.168 |
| Apnea | rs227731 | 17 | 54773238 | *NOG,C17orf67* | T | G | 0.52 | -0.026 | 0.004 | 2.40E-11 | -0.007 | 0.009 | 0.428 |
| Apnea | rs6567160 | 18 | 57829135 | *PMAIP1,MC4R* | T | C | 0.77 | 0.024 | 0.005 | 1.19E-31 | -0.046 | 0.011 | 0.000 |
| Apnea | rs35445111 | 19 | 32172047 | *TSHZ3,ZNF507* | G | A | 0.11 | -0.048 | 0.007 | 2.62E-12 | 0.009 | 0.015 | 0.558 |
| Apnea | rs17794954 | 20 | 50966307 | *ZFP64,TSHZ2* | T | C | 0.16 | -0.022 | 0.005 | 3.11E-14 | 0.012 | 0.013 | 0.324 |
| Apnea | rs6038517 | 20 | 6458205 | *FERMT1,BMP2* | G | A | 0.26 | 0.017 | 0.005 | 1.44E-08 | 0.006 | 0.010 | 0.569 |
| Apnea | rs6113592 | 20 | 22229505 | *PAX1,FOXA2* | G | A | 0.44 | -0.020 | 0.004 | 5.51E-11 | -0.007 | 0.010 | 0.507 |
| Apnea | rs2735309 | 21 | 40715313 | *HMGN1* | T | C | 0.33 | -0.021 | 0.004 | 9.10E-09 | -0.006 | 0.010 | 0.515 |
| Snoring | rs10835317 | 11 | 28389701 | *-* | G | A | 0.46 | -0.006 | 0.001 | 2.3E-08 | -0.010 | 0.010 | 0.316 |
| Snoring | rs10844664 | 12 | 33781709 | *-* | G | C | 0.37 | 0.006 | 0.001 | 2.1E-08 | -0.015 | 0.010 | 0.124 |
| Snoring | rs10878269 | 12 | 65791463 | *-* | T | C | 0.34 | 0.009 | 0.001 | 4.9E-18 | 0.009 | 0.010 | 0.365 |
| Snoring | rs12119849 | 1 | 96878072 | *-* | A | G | 0.08 | 0.011 | 0.002 | 3.9E-09 | -0.016 | 0.019 | 0.409 |
| Snoring | rs12429765 | 13 | 40745860 | *-* | G | A | 0.45 | -0.007 | 0.001 | 9.7E-12 | -0.003 | 0.009 | 0.786 |
| Snoring | rs13251292 | 8 | 71474355 | *-* | G | A | 0.37 | 0.007 | 0.001 | 1.5E-10 | -0.008 | 0.010 | 0.436 |
| Snoring | rs1374895 | 3 | 77615539 | *-* | T | C | 0.54 | -0.006 | 0.001 | 1.5E-08 | -0.011 | 0.009 | 0.223 |
| Snoring | rs180110 | 17 | 67930613 | *-* | A | G | 0.59 | 0.006 | 0.001 | 6.9E-10 | 0.014 | 0.009 | 0.131 |
| Snoring | rs183066 | 9 | 97517432 | *-* | T | C | 0.83 | 0.006 | 0.001 | 3.6E-08 | -0.001 | 0.033 | 0.970 |
| Snoring | rs227727 | 17 | 54776955 | *-* | T | A | 0.48 | 0.006 | 0.001 | 2.2E-09 | 0.008 | 0.009 | 0.411 |
| Snoring | rs2664303 | 14 | 99750520 | *-* | T | C | 0.39 | -0.007 | 0.001 | 3.1E-12 | -0.019 | 0.010 | 0.065 |
| Snoring | rs3862266 | 1 | 50822435 | *-* | A | G | 0.29 | 0.007 | 0.001 | 1.2E-10 | 0.002 | 0.010 | 0.851 |
| Snoring | rs4961747 | 9 | 16744269 | *-* | A | G | 0.19 | -0.007 | 0.001 | 2.6E-08 | 0.035 | 0.014 | 0.011 |
| Snoring | rs4976269 | 5 | 134452597 | *-* | A | G | 0.31 | -0.007 | 0.001 | 4.5E-11 | 0.011 | 0.010 | 0.291 |
| Snoring | rs6099273 | 20 | 55347828 | *-* | T | C | 0.23 | 0.007 | 0.001 | 3.8E-08 | -0.002 | 0.011 | 0.892 |
| Snoring | rs6117259 | 20 | 6420731 | *-* | C | T | 0.18 | 0.008 | 0.001 | 6.4E-09 | -0.012 | 0.012 | 0.343 |
| Snoring | rs61597598 | 2 | 156996626 | *-* | A | G | 0.13 | 0.011 | 0.002 | 1.1E-12 | -0.001 | 0.013 | 0.938 |
| Snoring | rs725861 | 10 | 9063776 | *-* | G | A | 0.21 | 0.009 | 0.001 | 9.7E-13 | 0.018 | 0.011 | 0.100 |
| Snoring | rs7829639 | 8 | 78215352 | *-* | G | A | 0.73 | 0.007 | 0.001 | 2.4E-10 | -0.011 | 0.011 | 0.324 |
| Snoring | rs8108822 | 19 | 32183171 | *-* | T | C | 0.15 | -0.011 | 0.002 | 7.7E-11 | 0.005 | 0.014 | 0.701 |
| Snoring | rs853241 | 3 | 64305763 | *-* | T | C | 0.34 | 0.007 | 0.001 | 1.1E-08 | -0.007 | 0.010 | 0.458 |
| Snoring | rs9521988 | 13 | 111559742 | *-* | A | G | 0.33 | -0.006 | 0.001 | 3.8E-08 | 0.012 | 0.010 | 0.199 |
| Snoring | rs9912001 | 17 | 7445372 | *-* | T | C | 0.33 | 0.007 | 0.001 | 8.8E-10 | 0.008 | 0.010 | 0.445 |

Chr, chromosome; EA, effect allele; EAF, effect allele frequency; NEA, non-effect allele; PAD, peripheral artery disease; SE, standard error; SNP, single nucleotide polymorphism.

### **Supplementary Table 3. Primary instrumental variables for peripheral artery disease and their associations with sleep-related traits**

| **Trait** | **SNP** | **Chr** | **Position** | **Nearby gene** | **EA** | **NEA** | **EAF** | **PAD** | | | **Sleep-related trait** | | |
| --- | --- | --- | --- | --- | --- | --- | --- | --- | --- | --- | --- | --- | --- |
|  |  |  |  |  |  |  |  | **Beta** | **SE** | **P** | **Beta** | **SE** | **P** |
| Napping | rs10851907 | 15 | 78915864 | *CHRNA3* | A | G | 0.41 | 0.058 | 0.005 | 1.49E-13 | 0.003 | 0.002 | 0.097 |
| Napping | rs11066301 | 12 | 112871372 | *PTPN11* | G | A | 0.43 | 0.058 | 0.010 | 2.96E-11 | -0.002 | 0.002 | 0.190 |
| Napping | rs118039278 | 6 | 160985526 | *LPA* | A | G | 0.08 | 0.231 | 0.016 | 1.57E-43 | 0.000 | 0.003 | 0.980 |
| Napping | rs138294113 | 19 | 11191729 | *(LDLR)* | C | T | 0.88 | 0.086 | 0.014 | 1.20E-10 | -0.004 | 0.002 | 0.150 |
| Napping | rs1537372 | 9 | 22103183 | *CDKN2B-AS1/9p21* | T | G | 0.43 | 0.113 | 0.009 | 4.32E-39 | 0.004 | 0.002 | 0.013 |
| Napping | rs1975514 | 13 | 110828891 | *COL4A1* | C | T | 0.37 | 0.049 | 0.005 | 8.32E-10 | 0.000 | 0.002 | 0.880 |
| Napping | rs2107595 | 7 | 19049388 | *(HDAC9)* | A | G | 0.15 | 0.077 | 0.014 | 2.49E-11 | 0.000 | 0.002 | 0.960 |
| Napping | rs3130968 | 6 | 31065071 | *(HLA-B)* | T | C | 0.17 | 0.068 | 0.010 | 3.16E-10 | -0.004 | 0.002 | 0.043 |
| Napping | rs322 | 8 | 19819217 | *LPL* | A | C | 0.73 | 0.058 | 0.010 | 2.53E-09 | 0.000 | 0.002 | 0.830 |
| Napping | rs4722172 | 7 | 22786532 | *(IL6)* | G | A | 0.22 | 0.077 | 0.014 | 3.65E-11 | 0.004 | 0.002 | 0.044 |
| Napping | rs4842266 | 12 | 79951566 | *RP11-359M6.3* | G | A | 0.30 | 0.058 | 0.010 | 1.01E-09 | 0.004 | 0.002 | 0.010 |
| Napping | rs505922 | 9 | 136149229 | *ABO* | C | T | 0.32 | 0.058 | 0.010 | 7.10E-11 | -0.002 | 0.002 | 0.310 |
| Napping | rs55784307 | 14 | 70501364 | *SMOC1* | A | C | 0.19 | 0.058 | 0.010 | 2.93E-08 | -0.001 | 0.002 | 0.630 |
| Napping | rs566125 | 11 | 102710471 | *MMP3* | T | C | 0.13 | 0.077 | 0.014 | 4.37E-09 | -0.003 | 0.002 | 0.180 |
| Napping | rs6025 | 1 | 169519049 | *F5* | T | C | 0.02 | 0.182 | 0.026 | 1.63E-12 | -0.011 | 0.005 | 0.029 |
| Napping | rs62084752 | 17 | 66089393 | *LOC732538* | C | G | 0.21 | 0.068 | 0.010 | 1.58E-10 | -0.001 | 0.002 | 0.530 |
| Napping | rs7476 | 11 | 46342834 | *CREB3L1* | C | A | 0.31 | 0.058 | 0.010 | 8.33E-10 | -0.006 | 0.002 | 0.001 |
| Napping | rs7528419 | 1 | 109817192 | *CELSR2/SORT1* | A | G | 0.78 | 0.068 | 0.010 | 2.54E-11 | 0.004 | 0.002 | 0.046 |
| Napping | rs7903146 | 10 | 114758349 | *TCF7L2* | T | C | 0.29 | 0.058 | 0.010 | 3.76E-11 | -0.003 | 0.002 | 0.077 |
| Sleep duration | rs6025 | 1 | 169519049 | *F5* | T | C | 0.02 | 0.182 | 0.026 | 1.63E-12 | -0.609 | 0.454 | 0.190 |
| Sleep duration | rs7528419 | 1 | 109817192 | *CELSR2/SORT1* | A | G | 0.78 | 0.068 | 0.010 | 2.54E-11 | 0.037 | 0.163 | 0.830 |
| Sleep duration | rs118039278 | 6 | 160985526 | *LPA* | A | G | 0.08 | 0.231 | 0.016 | 1.57E-43 | 0.085 | 0.252 | 0.760 |
| Sleep duration | rs3130968 | 6 | 31065071 | *(HLA-B)* | T | C | 0.17 | 0.068 | 0.010 | 3.16E-10 | -0.293 | 0.181 | 0.098 |
| Sleep duration | rs2107595 | 7 | 19049388 | *(HDAC9)* | A | G | 0.15 | 0.077 | 0.014 | 2.49E-11 | -0.065 | 0.189 | 0.750 |
| Sleep duration | rs4722172 | 7 | 22786532 | *(IL6)* | G | A | 0.22 | 0.077 | 0.014 | 3.65E-11 | 0.077 | 0.165 | 0.680 |
| Sleep duration | rs322 | 8 | 19819217 | *LPL* | A | C | 0.73 | 0.058 | 0.010 | 2.53E-09 | 0.091 | 0.153 | 0.520 |
| Sleep duration | rs1537372 | 9 | 22103183 | *CDKN2B-AS1/9p21* | T | G | 0.43 | 0.113 | 0.009 | 4.32E-39 | 0.032 | 0.138 | 0.870 |
| Sleep duration | rs505922 | 9 | 136149229 | *ABO* | C | T | 0.32 | 0.058 | 0.010 | 7.10E-11 | -0.524 | 0.146 | 0.000 |
| Sleep duration | rs7903146 | 10 | 114758349 | *TCF7L2* | T | C | 0.29 | 0.058 | 0.010 | 3.76E-11 | 0.120 | 0.150 | 0.440 |
| Sleep duration | rs566125 | 11 | 102710471 | *MMP3* | T | C | 0.13 | 0.077 | 0.014 | 4.37E-09 | -0.191 | 0.204 | 0.340 |
| Sleep duration | rs7476 | 11 | 46342834 | *CREB3L1* | C | A | 0.31 | 0.058 | 0.010 | 8.33E-10 | -0.496 | 0.147 | 0.001 |
| Sleep duration | rs11066301 | 12 | 112871372 | *PTPN11* | G | A | 0.43 | 0.058 | 0.010 | 2.96E-11 | 0.008 | 0.137 | 0.950 |
| Sleep duration | rs4842266 | 12 | 79951566 | *RP11-359M6.3* | G | A | 0.30 | 0.058 | 0.010 | 1.01E-09 | -0.234 | 0.148 | 0.110 |
| Sleep duration | rs1975514 | 13 | 110828891 | *COL4A1* | C | T | 0.37 | 0.049 | 0.005 | 8.32E-10 | -0.131 | 0.141 | 0.350 |
| Sleep duration | rs55784307 | 14 | 70501364 | *SMOC1* | A | C | 0.19 | 0.058 | 0.010 | 2.93E-08 | -0.133 | 0.175 | 0.490 |
| Sleep duration | rs10851907 | 15 | 78915864 | *CHRNA3* | A | G | 0.41 | 0.058 | 0.005 | 1.49E-13 | 0.004 | 0.139 | 0.990 |
| Sleep duration | rs62084752 | 17 | 66089393 | *LOC732538* | C | G | 0.21 | 0.068 | 0.010 | 1.58E-10 | 0.066 | 0.166 | 0.670 |
| Sleep duration | rs138294113 | 19 | 11191729 | *(LDLR)* | C | T | 0.88 | 0.086 | 0.014 | 1.20E-10 | 0.349 | 0.210 | 0.098 |
| Sleep_long | rs10851907 | 15 | 78915864 | *CHRNA3* | A | G | 0.41 | 0.058 | 0.005 | 1.49E-13 | -0.003 | 0.007 | 0.620 |
| Sleep_long | rs11066301 | 12 | 112871372 | *PTPN11* | G | A | 0.43 | 0.058 | 0.010 | 2.96E-11 | 0.004 | 0.007 | 0.550 |
| Sleep_long | rs118039278 | 6 | 160985526 | *LPA* | A | G | 0.08 | 0.231 | 0.016 | 1.57E-43 | 0.004 | 0.012 | 0.720 |
| Sleep_long | rs138294113 | 19 | 11191729 | *(LDLR)* | C | T | 0.88 | 0.086 | 0.014 | 1.20E-10 | 0.001 | 0.010 | 0.880 |
| Sleep_long | rs1537372 | 9 | 22103183 | *CDKN2B-AS1/9p21* | T | G | 0.43 | 0.113 | 0.009 | 4.32E-39 | 0.010 | 0.007 | 0.120 |
| Sleep_long | rs1975514 | 13 | 110828891 | *COL4A1* | C | T | 0.37 | 0.049 | 0.005 | 8.32E-10 | 0.000 | 0.007 | 0.950 |
| Sleep_long | rs2107595 | 7 | 19049388 | *(HDAC9)* | A | G | 0.15 | 0.077 | 0.014 | 2.49E-11 | 0.015 | 0.009 | 0.100 |
| Sleep_long | rs3130968 | 6 | 31065071 | *(HLA-B)* | T | C | 0.17 | 0.068 | 0.010 | 3.16E-10 | -0.001 | 0.009 | 0.880 |
| Sleep_long | rs322 | 8 | 19819217 | *LPL* | A | C | 0.73 | 0.058 | 0.010 | 2.53E-09 | -0.012 | 0.007 | 0.098 |
| Sleep_long | rs4722172 | 7 | 22786532 | *(IL6)* | G | A | 0.22 | 0.077 | 0.014 | 3.65E-11 | 0.009 | 0.008 | 0.250 |
| Sleep_long | rs4842266 | 12 | 79951566 | *RP11-359M6.3* | G | A | 0.30 | 0.058 | 0.010 | 1.01E-09 | -0.005 | 0.007 | 0.490 |
| Sleep_long | rs505922 | 9 | 136149229 | *ABO* | C | T | 0.32 | 0.058 | 0.010 | 7.10E-11 | -0.006 | 0.007 | 0.370 |
| Sleep_long | rs55784307 | 14 | 70501364 | *SMOC1* | A | C | 0.19 | 0.058 | 0.010 | 2.93E-08 | -0.009 | 0.008 | 0.280 |
| Sleep_long | rs566125 | 11 | 102710471 | *MMP3* | T | C | 0.13 | 0.077 | 0.014 | 4.37E-09 | -0.007 | 0.010 | 0.450 |
| Sleep_long | rs6025 | 1 | 169519049 | *F5* | T | C | 0.02 | 0.182 | 0.026 | 1.63E-12 | -0.031 | 0.022 | 0.150 |
| Sleep_long | rs62084752 | 17 | 66089393 | *LOC732538* | C | G | 0.21 | 0.068 | 0.010 | 1.58E-10 | 0.007 | 0.008 | 0.390 |
| Sleep_long | rs7476 | 11 | 46342834 | *CREB3L1* | C | A | 0.31 | 0.058 | 0.010 | 8.33E-10 | -0.007 | 0.007 | 0.290 |
| Sleep_long | rs7528419 | 1 | 109817192 | *CELSR2/SORT1* | A | G | 0.78 | 0.068 | 0.010 | 2.54E-11 | 0.006 | 0.008 | 0.450 |
| Sleep_long | rs7903146 | 10 | 114758349 | *TCF7L2* | T | C | 0.29 | 0.058 | 0.010 | 3.76E-11 | -0.010 | 0.007 | 0.150 |
| Sleep_short | rs10851907 | 15 | 78915864 | *CHRNA3* | A | G | 0.41 | 0.058 | 0.005 | 1.49E-13 | 0.001 | 0.005 | 0.780 |
| Sleep_short | rs11066301 | 12 | 112871372 | *PTPN11* | G | A | 0.43 | 0.058 | 0.010 | 2.96E-11 | 0.001 | 0.005 | 0.810 |
| Sleep_short | rs118039278 | 6 | 160985526 | *LPA* | A | G | 0.08 | 0.231 | 0.016 | 1.57E-43 | -0.002 | 0.009 | 0.850 |
| Sleep_short | rs138294113 | 19 | 11191729 | *(LDLR)* | C | T | 0.88 | 0.086 | 0.014 | 1.20E-10 | -0.006 | 0.008 | 0.450 |
| Sleep_short | rs1537372 | 9 | 22103183 | *CDKN2B-AS1/9p21* | T | G | 0.43 | 0.113 | 0.009 | 4.32E-39 | 0.005 | 0.005 | 0.310 |
| Sleep_short | rs1975514 | 13 | 110828891 | *COL4A1* | C | T | 0.37 | 0.049 | 0.005 | 8.32E-10 | 0.005 | 0.005 | 0.340 |
| Sleep_short | rs2107595 | 7 | 19049388 | *(HDAC9)* | A | G | 0.15 | 0.077 | 0.014 | 2.49E-11 | 0.019 | 0.007 | 0.008 |
| Sleep_short | rs3130968 | 6 | 31065071 | *(HLA-B)* | T | C | 0.17 | 0.068 | 0.010 | 3.16E-10 | 0.009 | 0.007 | 0.160 |
| Sleep_short | rs322 | 8 | 19819217 | *LPL* | A | C | 0.73 | 0.058 | 0.010 | 2.53E-09 | -0.007 | 0.006 | 0.200 |
| Sleep_short | rs4722172 | 7 | 22786532 | *(IL6)* | G | A | 0.22 | 0.077 | 0.014 | 3.65E-11 | 0.001 | 0.006 | 0.870 |
| Sleep_short | rs4842266 | 12 | 79951566 | *RP11-359M6.3* | G | A | 0.30 | 0.058 | 0.010 | 1.01E-09 | 0.006 | 0.005 | 0.260 |
| Sleep_short | rs505922 | 9 | 136149229 | *ABO* | C | T | 0.32 | 0.058 | 0.010 | 7.10E-11 | 0.014 | 0.005 | 0.008 |
| Sleep_short | rs55784307 | 14 | 70501364 | *SMOC1* | A | C | 0.19 | 0.058 | 0.010 | 2.93E-08 | 0.000 | 0.006 | 0.980 |
| Sleep_short | rs566125 | 11 | 102710471 | *MMP3* | T | C | 0.13 | 0.077 | 0.014 | 4.37E-09 | 0.003 | 0.008 | 0.690 |
| Sleep_short | rs6025 | 1 | 169519049 | *F5* | T | C | 0.02 | 0.182 | 0.026 | 1.63E-12 | 0.016 | 0.017 | 0.350 |
| Sleep_short | rs62084752 | 17 | 66089393 | *LOC732538* | C | G | 0.21 | 0.068 | 0.010 | 1.58E-10 | 0.005 | 0.006 | 0.360 |
| Sleep_short | rs7476 | 11 | 46342834 | *CREB3L1* | C | A | 0.31 | 0.058 | 0.010 | 8.33E-10 | 0.017 | 0.005 | 0.002 |
| Sleep_short | rs7528419 | 1 | 109817192 | *CELSR2/SORT1* | A | G | 0.78 | 0.068 | 0.010 | 2.54E-11 | 0.006 | 0.006 | 0.290 |
| Sleep_short | rs7903146 | 10 | 114758349 | *TCF7L2* | T | C | 0.29 | 0.058 | 0.010 | 3.76E-11 | -0.004 | 0.006 | 0.460 |
| Insomnia | rs6025 | 1 | 169519049 | *F5* | T | C | 0.77 | 0.182 | 0.026 | 1.63E-12 | -0.010 | 0.017 | 0.541 |
| Insomnia | rs7528419 | 1 | 109817192 | *CELSR2/SORT1* | A | G | 0.03 | 0.068 | 0.010 | 2.54E-11 | 0.012 | 0.006 | 0.044 |
| Insomnia | rs118039278 | 6 | 160985526 | *LPA* | A | G | 0.07 | 0.231 | 0.016 | 1.57E-43 | -0.005 | 0.010 | 0.601 |
| Insomnia | rs3130968 | 6 | 31065071 | *(HLA-B)* | T | C | 0.14 | 0.068 | 0.010 | 3.16E-10 | -0.006 | 0.007 | 0.370 |
| Insomnia | rs2107595 | 7 | 19049388 | *(HDAC9)* | A | G | 0.19 | 0.077 | 0.014 | 2.49E-11 | 0.013 | 0.007 | 0.075 |
| Insomnia | rs4722172 | 7 | 22786532 | *(IL6)* | G | A | 0.20 | 0.077 | 0.014 | 3.65E-11 | 0.002 | 0.006 | 0.782 |
| Insomnia | rs322 | 8 | 19819217 | *LPL* | A | C | 0.71 | 0.058 | 0.010 | 2.53E-09 | -0.005 | 0.006 | 0.421 |
| Insomnia | rs1537372 | 9 | 22103183 | *CDKN2B-AS1/9p21* | T | G | 0.33 | 0.113 | 0.009 | 4.32E-39 | -0.003 | 0.005 | 0.615 |
| Insomnia | rs505922 | 9 | 136149229 | *ABO* | C | T | 0.42 | 0.058 | 0.010 | 7.10E-11 | 0.009 | 0.005 | 0.087 |
| Insomnia | rs7903146 | 10 | 114758349 | *TCF7L2* | T | C | 0.29 | 0.058 | 0.010 | 3.76E-11 | -0.002 | 0.006 | 0.674 |
| Insomnia | rs566125 | 11 | 102710471 | *MMP3* | T | C | 0.13 | 0.077 | 0.014 | 4.37E-09 | 0.014 | 0.008 | 0.067 |
| Insomnia | rs7476 | 11 | 46342834 | *CREB3L1* | C | A | 0.36 | 0.058 | 0.010 | 8.33E-10 | 0.010 | 0.006 | 0.060 |
| Insomnia | rs11066301 | 12 | 112871372 | *PTPN11* | G | A | 0.41 | 0.058 | 0.010 | 2.96E-11 | -0.005 | 0.005 | 0.360 |
| Insomnia | rs4842266 | 12 | 79951566 | *RP11-359M6.3* | G | A | 0.39 | 0.058 | 0.010 | 1.01E-09 | -0.001 | 0.006 | 0.896 |
| Insomnia | rs1975514 | 13 | 110828891 | *COL4A1* | C | T | 0.36 | 0.049 | 0.005 | 8.32E-10 | 0.001 | 0.005 | 0.870 |
| Insomnia | rs55784307 | 14 | 70501364 | *SMOC1* | A | C | 0.18 | 0.058 | 0.010 | 2.93E-08 | -0.001 | 0.007 | 0.946 |
| Insomnia | rs10851907 | 15 | 78915864 | *CHRNA3* | A | G | 0.41 | 0.058 | 0.005 | 1.49E-13 | -0.006 | 0.005 | 0.263 |
| Insomnia | rs62084752 | 17 | 66089393 | *LOC732538* | C | G | 0.22 | 0.068 | 0.010 | 1.58E-10 | 0.016 | 0.006 | 0.010 |
| Insomnia | rs138294113 | 19 | 11191729 | *(LDLR)* | C | T | 0.88 | 0.086 | 0.014 | 1.20E-10 | 0.014 | 0.008 | 0.084 |

Chr, chromosome; EA, effect allele; EAF, effect allele frequency; NEA, non-effect allele; PAD, peripheral artery disease; SE, standard error; SNP, single nucleotide polymorphism.

**Supplementary Table 4. Complementary instrumental variables for peripheral artery disease and their associations with sleep-related** **traits**

| **Trait** | **SNP** | **Chr** | **Position** | **Nearby gene** | **EA** | **NEA** | **EAF** | **PAD** | | | **Sleep-related trait** | | |
| --- | --- | --- | --- | --- | --- | --- | --- | --- | --- | --- | --- | --- | --- |
|  |  |  |  |  |  |  |  | **Beta** | **SE** | **P** | **Beta** | **SE** | **P** |
| Sleep duration | rs6025 | 1 | 169519049 | *F5* | T | C | 0.02 | 0.184 | 0.028 | 8.81E-11 | -0.526 | 0.243 | 0.029 |
| Sleep duration | rs7528419 | 1 | 109817192 | *CELSR2* | A | G | 0.78 | 0.062 | 0.011 | 7.12E-09 | 0.185 | 0.087 | 0.046 |
| Sleep duration | rs6841581 | 4 | 148401190 | *EDNRA* | A | G | 0.14 | 0.074 | 0.012 | 1.92E-09 | 0.208 | 0.105 | 0.050 |
| Sleep duration | rs186696265 | 6 | 161111700 | *-* | T | C | 0.01 | 0.347 | 0.043 | 6.62E-16 | 0.584 | 0.302 | 0.061 |
| Sleep duration | rs3094087 | 6 | 31061561 | *-* | C | T | 0.17 | 0.074 | 0.013 | 5.03E-09 | -0.200 | 0.097 | 0.046 |
| Sleep duration | rs55730499 | 6 | 161005610 | *LPA* | T | C | 0.08 | 0.195 | 0.018 | 1.87E-26 | 0.025 | 0.134 | 0.910 |
| Sleep duration | rs2107595 | 7 | 19049388 | *-* | A | G | 0.15 | 0.071 | 0.012 | 4.77E-09 | -0.004 | 0.101 | 0.960 |
| Sleep duration | rs1537372 | 9 | 22103183 | *CDKN2B-AS1* | T | G | 0.43 | 0.107 | 0.010 | 1.96E-28 | 0.184 | 0.074 | 0.013 |
| Sleep duration | rs10786400 | 10 | 99926859 | *R3HCC1L* | A | G | 0.68 | 0.060 | 0.011 | 1.30E-08 | -0.027 | 0.078 | 0.780 |
| Sleep duration | rs7903146 | 10 | 114758349 | *TCF7L2* | T | C | 0.29 | 0.058 | 0.010 | 1.81E-09 | -0.140 | 0.080 | 0.077 |
| Sleep duration | rs3740973 | 11 | 46422237 | *AMBRA1* | A | G | 0.29 | 0.064 | 0.010 | 5.82E-10 | -0.277 | 0.080 | 0.001 |
| Sleep duration | rs626750 | 11 | 102720945 | *-* | A | G | 0.17 | 0.072 | 0.012 | 4.11E-10 | -0.042 | 0.097 | 0.660 |
| Sleep duration | rs11066301 | 12 | 112871372 | *PTPN11* | G | A | 0.43 | 0.053 | 0.010 | 4.34E-08 | -0.098 | 0.074 | 0.190 |
| Sleep duration | rs4842266 | 12 | 79951566 | *RP11-359M6.3* | G | A | 0.30 | 0.059 | 0.011 | 3.60E-08 | 0.206 | 0.079 | 0.010 |
| Sleep duration | rs227419 | 14 | 70459370 | *SMOC1* | C | T | 0.57 | 0.054 | 0.009 | 5.15E-09 | 0.042 | 0.074 | 0.570 |
| Sleep duration | rs667282 | 15 | 78863472 | *CHRNA5* | T | C | 0.77 | 0.064 | 0.011 | 8.84E-10 | 0.213 | 0.087 | 0.014 |
| Sleep duration | rs17817497 | 16 | 53815435 | *FTO* | C | T | 0.39 | 0.062 | 0.010 | 1.38E-10 | -0.347 | 0.075 | 0.000 |
| Sleep duration | rs244420 | 16 | 69657996 | *NFAT5* | T | G | 0.59 | 0.063 | 0.010 | 5.76E-11 | 0.238 | 0.074 | 0.001 |
| Sleep_short | rs10786400 | 10 | 99926859 | *R3HCC1L* | A | G | 0.68 | 0.060 | 0.011 | 1.30E-08 | 0.010 | 0.005 | 0.070 |
| Sleep_short | rs11066301 | 12 | 112871372 | *PTPN11* | G | A | 0.43 | 0.053 | 0.010 | 4.34E-08 | 0.001 | 0.005 | 0.810 |
| Sleep_short | rs1537372 | 9 | 22103183 | *CDKN2B-AS1* | T | G | 0.43 | 0.107 | 0.010 | 1.96E-28 | 0.005 | 0.005 | 0.310 |
| Sleep_short | rs17817497 | 16 | 53815435 | *FTO* | C | T | 0.39 | 0.062 | 0.010 | 1.38E-10 | 0.016 | 0.005 | 0.002 |
| Sleep_short | rs186696265 | 6 | 161111700 | *-* | T | C | 0.01 | 0.347 | 0.043 | 6.62E-16 | 0.011 | 0.021 | 0.590 |
| Sleep_short | rs2107595 | 7 | 19049388 | *-* | A | G | 0.15 | 0.071 | 0.012 | 4.77E-09 | 0.019 | 0.007 | 0.008 |
| Sleep_short | rs227419 | 14 | 70459370 | *SMOC1* | C | T | 0.57 | 0.054 | 0.009 | 5.15E-09 | -0.004 | 0.005 | 0.490 |
| Sleep_short | rs244420 | 16 | 69657996 | *NFAT5* | T | G | 0.59 | 0.063 | 0.010 | 5.76E-11 | -0.006 | 0.005 | 0.210 |
| Sleep_short | rs3094087 | 6 | 31061561 | *-* | C | T | 0.17 | 0.074 | 0.013 | 5.03E-09 | 0.009 | 0.007 | 0.150 |
| Sleep_short | rs3740973 | 11 | 46422237 | *AMBRA1* | A | G | 0.29 | 0.064 | 0.010 | 5.82E-10 | 0.016 | 0.006 | 0.004 |
| Sleep_short | rs4842266 | 12 | 79951566 | *RP11-359M6.3* | G | A | 0.30 | 0.059 | 0.011 | 3.60E-08 | 0.006 | 0.005 | 0.260 |
| Sleep_short | rs55730499 | 6 | 161005610 | *LPA* | T | C | 0.08 | 0.195 | 0.018 | 1.87E-26 | -0.002 | 0.009 | 0.860 |
| Sleep_short | rs6025 | 1 | 169519049 | *F5* | T | C | 0.02 | 0.184 | 0.028 | 8.81E-11 | 0.016 | 0.017 | 0.350 |
| Sleep_short | rs626750 | 11 | 102720945 | *-* | A | G | 0.17 | 0.072 | 0.012 | 4.11E-10 | -0.001 | 0.007 | 0.970 |
| Sleep_short | rs667282 | 15 | 78863472 | *CHRNA5* | T | C | 0.77 | 0.064 | 0.011 | 8.84E-10 | 0.006 | 0.006 | 0.310 |
| Sleep_short | rs6841581 | 4 | 148401190 | *EDNRA* | A | G | 0.14 | 0.074 | 0.012 | 1.92E-09 | -0.002 | 0.007 | 0.790 |
| Sleep_short | rs7528419 | 1 | 109817192 | *CELSR2* | A | G | 0.78 | 0.062 | 0.011 | 7.12E-09 | 0.006 | 0.006 | 0.290 |
| Sleep_short | rs7903146 | 10 | 114758349 | *TCF7L2* | T | C | 0.29 | 0.058 | 0.010 | 1.81E-09 | -0.004 | 0.006 | 0.460 |
| Sleep_long | rs10786400 | 10 | 99926859 | *R3HCC1L* | A | G | 0.68 | 0.060 | 0.011 | 1.30E-08 | -0.001 | 0.007 | 0.930 |
| Sleep_long | rs11066301 | 12 | 112871372 | *PTPN11* | G | A | 0.43 | 0.053 | 0.010 | 4.34E-08 | 0.004 | 0.007 | 0.550 |
| Sleep_long | rs1537372 | 9 | 22103183 | *CDKN2B-AS1* | T | G | 0.43 | 0.107 | 0.010 | 1.96E-28 | 0.010 | 0.007 | 0.120 |
| Sleep_long | rs17817497 | 16 | 53815435 | *FTO* | C | T | 0.39 | 0.062 | 0.010 | 1.38E-10 | -0.032 | 0.007 | 0.000 |
| Sleep_long | rs186696265 | 6 | 161111700 | *-* | T | C | 0.02 | 0.347 | 0.043 | 6.62E-16 | 0.083 | 0.027 | 0.002 |
| Sleep_long | rs2107595 | 7 | 19049388 | *-* | A | G | 0.15 | 0.071 | 0.012 | 4.77E-09 | 0.015 | 0.009 | 0.100 |
| Sleep_long | rs227419 | 14 | 70459370 | *SMOC1* | C | T | 0.58 | 0.054 | 0.009 | 5.15E-09 | -0.002 | 0.007 | 0.750 |
| Sleep_long | rs244420 | 16 | 69657996 | *NFAT5* | T | G | 0.59 | 0.063 | 0.010 | 5.76E-11 | 0.021 | 0.007 | 0.002 |
| Sleep_long | rs3094087 | 6 | 31061561 | *-* | C | T | 0.17 | 0.074 | 0.013 | 5.03E-09 | -0.001 | 0.009 | 0.880 |
| Sleep_long | rs3740973 | 11 | 46422237 | *AMBRA1* | A | G | 0.29 | 0.064 | 0.010 | 5.82E-10 | -0.007 | 0.007 | 0.330 |
| Sleep_long | rs4842266 | 12 | 79951566 | *RP11-359M6.3* | G | A | 0.30 | 0.059 | 0.011 | 3.60E-08 | -0.005 | 0.007 | 0.490 |
| Sleep_long | rs55730499 | 6 | 161005610 | *LPA* | T | C | 0.08 | 0.195 | 0.018 | 1.87E-26 | 0.006 | 0.012 | 0.640 |
| Sleep_long | rs6025 | 1 | 169519049 | *F5* | T | C | 0.02 | 0.184 | 0.028 | 8.81E-11 | -0.031 | 0.022 | 0.150 |
| Sleep_long | rs626750 | 11 | 102720945 | *-* | A | G | 0.17 | 0.072 | 0.012 | 4.11E-10 | -0.009 | 0.009 | 0.270 |
| Sleep_long | rs667282 | 15 | 78863472 | *CHRNA5* | T | C | 0.77 | 0.064 | 0.011 | 8.84E-10 | 0.015 | 0.008 | 0.058 |
| Sleep_long | rs6841581 | 4 | 148401190 | *EDNRA* | A | G | 0.14 | 0.074 | 0.012 | 1.92E-09 | 0.010 | 0.009 | 0.310 |
| Sleep_long | rs7528419 | 1 | 109817192 | *CELSR2* | A | G | 0.78 | 0.062 | 0.011 | 7.12E-09 | 0.006 | 0.008 | 0.450 |
| Sleep_long | rs7903146 | 10 | 114758349 | *TCF7L2* | T | C | 0.29 | 0.058 | 0.010 | 1.81E-09 | -0.010 | 0.007 | 0.150 |
| Napping | rs10786400 | 10 | 99926859 | *R3HCC1L* | A | G | 0.68 | 0.060 | 0.011 | 1.30E-08 | -0.001 | 0.002 | 0.780 |
| Napping | rs11066301 | 12 | 112871372 | *PTPN11* | G | A | 0.43 | 0.053 | 0.010 | 4.34E-08 | -0.002 | 0.002 | 0.190 |
| Napping | rs1537372 | 9 | 22103183 | *CDKN2B-AS1* | T | G | 0.43 | 0.107 | 0.010 | 1.96E-28 | 0.004 | 0.002 | 0.013 |
| Napping | rs17817497 | 16 | 53815435 | *FTO* | C | T | 0.39 | 0.062 | 0.010 | 1.38E-10 | -0.008 | 0.002 | 0.000 |
| Napping | rs186696265 | 6 | 161111700 | *-* | T | C | 0.01 | 0.347 | 0.043 | 6.62E-16 | 0.013 | 0.007 | 0.061 |
| Napping | rs2107595 | 7 | 19049388 | *-* | A | G | 0.15 | 0.071 | 0.012 | 4.77E-09 | 0.000 | 0.002 | 0.960 |
| Napping | rs227419 | 14 | 70459370 | *SMOC1* | C | T | 0.57 | 0.054 | 0.009 | 5.15E-09 | 0.001 | 0.002 | 0.570 |
| Napping | rs244420 | 16 | 69657996 | *NFAT5* | T | G | 0.59 | 0.063 | 0.010 | 5.76E-11 | 0.005 | 0.002 | 0.001 |
| Napping | rs3094087 | 6 | 31061561 | *-* | C | T | 0.17 | 0.074 | 0.013 | 5.03E-09 | -0.004 | 0.002 | 0.046 |
| Napping | rs3740973 | 11 | 46422237 | *AMBRA1* | A | G | 0.29 | 0.064 | 0.010 | 5.82E-10 | -0.006 | 0.002 | 0.001 |
| Napping | rs4842266 | 12 | 79951566 | *RP11-359M6.3* | G | A | 0.30 | 0.059 | 0.011 | 3.60E-08 | 0.004 | 0.002 | 0.010 |
| Napping | rs55730499 | 6 | 161005610 | *LPA* | T | C | 0.08 | 0.195 | 0.018 | 1.87E-26 | 0.001 | 0.003 | 0.910 |
| Napping | rs6025 | 1 | 169519049 | *F5* | T | C | 0.02 | 0.184 | 0.028 | 8.81E-11 | -0.011 | 0.005 | 0.029 |
| Napping | rs626750 | 11 | 102720945 | *-* | A | G | 0.17 | 0.072 | 0.012 | 4.11E-10 | -0.001 | 0.002 | 0.660 |
| Napping | rs667282 | 15 | 78863472 | *CHRNA5* | T | C | 0.77 | 0.064 | 0.011 | 8.84E-10 | 0.005 | 0.002 | 0.014 |
| Napping | rs6841581 | 4 | 148401190 | *EDNRA* | A | G | 0.14 | 0.074 | 0.012 | 1.92E-09 | 0.005 | 0.002 | 0.050 |
| Napping | rs7528419 | 1 | 109817192 | *CELSR2* | A | G | 0.78 | 0.062 | 0.011 | 7.12E-09 | 0.004 | 0.002 | 0.046 |
| Napping | rs7903146 | 10 | 114758349 | *TCF7L2* | T | C | 0.29 | 0.058 | 0.010 | 1.81E-09 | -0.003 | 0.002 | 0.077 |
| Insomnia | rs6025 | 1 | 169519049 | *F5* | T | C | 0.77 | 0.184 | 0.028 | 8.81E-11 | -0.010 | 0.017 | 0.541 |
| Insomnia | rs7528419 | 1 | 109817192 | *CELSR2* | A | G | 0.03 | 0.062 | 0.011 | 7.12E-09 | 0.012 | 0.006 | 0.044 |
| Insomnia | rs6841581 | 4 | 148401190 | *EDNRA* | A | G | 0.18 | 0.074 | 0.012 | 1.92E-09 | 0.008 | 0.007 | 0.251 |
| Insomnia | rs186696265 | 6 | 161111700 | *-* | T | C | 0.14 | 0.347 | 0.043 | 6.62E-16 | 0.003 | 0.021 | 0.891 |
| Insomnia | rs3094087 | 6 | 31061561 | *-* | C | T | 0.07 | 0.074 | 0.013 | 5.03E-09 | -0.006 | 0.007 | 0.382 |
| Insomnia | rs55730499 | 6 | 161005610 | *LPA* | T | C | 0.01 | 0.195 | 0.018 | 1.87E-26 | -0.003 | 0.009 | 0.736 |
| Insomnia | rs2107595 | 7 | 19049388 | *-* | A | G | 0.19 | 0.071 | 0.012 | 4.77E-09 | 0.013 | 0.007 | 0.075 |
| Insomnia | rs1537372 | 9 | 22103183 | *CDKN2B-AS1* | T | G | 0.42 | 0.107 | 0.010 | 1.96E-28 | -0.003 | 0.005 | 0.615 |
| Insomnia | rs10786400 | 10 | 99926859 | *R3HCC1L* | A | G | 0.29 | 0.060 | 0.011 | 1.30E-08 | 0.006 | 0.005 | 0.312 |
| Insomnia | rs7903146 | 10 | 114758349 | *TCF7L2* | T | C | 0.70 | 0.058 | 0.010 | 1.81E-09 | -0.002 | 0.006 | 0.674 |
| Insomnia | rs3740973 | 11 | 46422237 | *AMBRA1* | A | G | 0.37 | 0.064 | 0.010 | 5.82E-10 | 0.005 | 0.006 | 0.381 |
| Insomnia | rs626750 | 11 | 102720945 | *-* | A | G | 0.20 | 0.072 | 0.012 | 4.11E-10 | 0.007 | 0.007 | 0.319 |
| Insomnia | rs11066301 | 12 | 112871372 | *PTPN11* | G | A | 0.41 | 0.053 | 0.010 | 4.34E-08 | -0.005 | 0.005 | 0.360 |
| Insomnia | rs4842266 | 12 | 79951566 | *RP11-359M6.3* | G | A | 0.41 | 0.059 | 0.011 | 3.60E-08 | -0.001 | 0.006 | 0.896 |
| Insomnia | rs227419 | 14 | 70459370 | *SMOC1* | C | T | 0.59 | 0.054 | 0.009 | 5.15E-09 | 0.005 | 0.005 | 0.311 |
| Insomnia | rs667282 | 15 | 78863472 | *CHRNA5* | T | C | 0.74 | 0.064 | 0.011 | 8.84E-10 | 0.001 | 0.006 | 0.896 |
| Insomnia | rs17817497 | 16 | 53815435 | *FTO* | C | T | 0.37 | 0.062 | 0.010 | 1.38E-10 | 0.002 | 0.005 | 0.761 |
| Insomnia | rs244420 | 16 | 69657996 | *NFAT5* | T | G | 0.62 | 0.063 | 0.010 | 5.76E-11 | 0.004 | 0.005 | 0.475 |

Chr, chromosome; EA, effect allele; EAF, effect allele frequency; NEA, non-effect allele; PAD, peripheral artery disease; SE, standard error; SNP, single nucleotide polymorphism.

### **Supplementary Table 5. Associations of sleep quality traits with risk of peripheral artery disease in Swedish adults (the SIMPLER study)**

| **Sleep traits** | **Cases** | **Total** | **Person-years** | **Model 1** | | **Model 2** | | **Model 3** | |
| --- | --- | --- | --- | --- | --- | --- | --- | --- | --- |
|  |  |  |  | **OR** | ***P*** | **OR** | ***P*** | **OR** | ***P*** |
| **Difficulty falling asleep (n = 53 416)** | | | | | | | | | |
| Never | 274 | 11 249 | 103 103 | 1.00 | Ref | 1.00 | Ref | 1.00 | Ref |
| Seldom | 725 | 30 148 | 279 030 | 1.01 (0.88, 1.16) | 0.923 | 1.02 (0.88, 1.17) | 0.798 | 1.02 (0.88, 1.17) | 0.823 |
| Often, mostly, always | 282 | 10 168 | 90 836 | 1.21 (1.02, 1.44) | 0.031 | 1.13 (0.95, 1.35) | 0.155 | 1.07 (0.90, 1.27) | 0.441 |
| **Repeatedly waking up with difficulty falling asleep (n = 53 416)** | | | | | | | | | |
| Never | 213 | 8 452 | 77 139 | 1.00 | Ref | 1.00 | Ref | 1.00 | Ref |
| Seldom | 653 | 27 859 | 258 137 | 0.91 (0.78, 1.06) | 0.232 | 0.98 (0.84, 1.14) | 0.769 | 0.96 (0.82, 1.12) | 0.575 |
| Often, mostly, always | 370 | 13 951 | 127 058 | 1.05 (0.89, 1.25) | 0.540 | 1.12 (0.94, 1.33) | 0.200 | 1.04 (0.88, 1.24) | 0.624 |
| **Premature awakening (n = 53 416)** | | | | | | | | | |
| Never | 221 | 8 161 | 74 582 | 1.00 | Ref | 1.00 | Ref | 1.00 | Ref |
| Seldom | 642 | 26 764 | 248 197 | 0.87 (0.75, 1.01) | 0.073 | 0.92 (0.79, 1.07) | 0.300 | 0.94 (0.81, 1.09) | 0.413 |
| Often, mostly, always | 366 | 15 101 | 137 724 | 0.88 (0.74, 1.04) | 0.141 | 0.93 (0.79, 1.1) | 0.406 | 0.91 (0.77, 1.08) | 0.271 |
| **Disturbed or restless sleep (n = 53 416)** | | | | | | | | | |
| Never | 285 | 10 747 | 96 510 | 1.00 | Ref | 1.00 | Ref | 1.00 | Ref |
| Seldom | 641 | 26 644 | 247 339 | 0.96 (0.84, 1.11) | 0.581 | 1.00 (0.87, 1.15) | 0.950 | 1.00 (0.87, 1.14) | 0.952 |
| Often, mostly, always | 298 | 12 177 | 112 313 | 1.05 (0.89, 1.23) | 0.591 | 1.07 (0.91, 1.26) | 0.419 | 1.00 (0.85, 1.18) | 0.987 |
| **Sleep apnea (n = 53 416)** | | | | | | | | | |
| Never | 886 | 36 952 | 339 109 | 1.00 | Ref | 1.00 | Ref | 1.00 | Ref |
| Seldom | 204 | 8 362 | 77 442 | 1.04 (0.89, 1.22) | 0.601 | 1.00 (0.86, 1.17) | 0.990 | 0.97 (0.83, 1.13) | 0.668 |
| Often, mostly, always | 102 | 3 853 | 36 030 | 1.19 (0.96, 1.46) | 0.104 | 1.08 (0.88, 1.33) | 0.444 | 1.00 (0.81, 1.23) | 0.998 |
| **Disturbing snoring (n = 53 416)** | | | | | | | | | |
| Never | 431 | 17 746 | 157 872 | 1.00 | Ref | 1.00 | Ref | 1.00 | Ref |
| Seldom | 491 | 20 640 | 192 190 | 1.02 (0.89, 1.16) | 0.817 | 1.01 (0.89, 1.16) | 0.862 | 1.01 (0.89, 1.16) | 0.852 |
| Often, mostly, always | 302 | 11 893 | 112 696 | 1.15 (0.98, 1.34) | 0.078 | 1.05 (0.90, 1.23) | 0.494 | 1.03 (0.88, 1.20) | 0.722 |

Missing information was 2.5% for difficulty falling asleep, 4.8% for repeatedly waking up with difficulty falling asleep, 5.0% for premature awakening, 6.0% for disturbed or restless sleep, 6.8 % for sleep apnea, and 4.7% for disturbing snoring. Individuals with missing information were classified in a separate missing group. Model 1 adjusted for age and sex. Model 2 adjusted for age, sex, education level, smoking status, physical activity, and diet quality. Model 3 adjusted for age, sex, education level, smoking status, physical activity, diet quality, and potential mediators including body mass index and baseline history of hypertension, hypercholesteremia and diabetes.

### **Supplementary Table 6. Demographic features of participants in MVP**

| **Characteristic** | **Overall,**  **N = 135,740** | **5 to 7,**  **N = 48,482** | **7 to 8,**  **N = 37,240** | **8 to 10,**  **N = 50,018** |
| --- | --- | --- | --- | --- |
| Age, mean (SD) | 67 (11) | 64 (11) | 67 (10) | 69 (10) |
| Sex |  |  |  |  |
| Female | 10,283 (7.6%) | 4,442 (9.2%) | 2,605 (7.0%) | 3,236 (6.5%) |
| Male | 125,457 (92%) | 44,040 (91%) | 34,635 (93%) | 46,782 (94%) |
| Body Mass Index (BMI), mean (SD) | 30.6 (6.0) | 31.0 (6.2) | 30.3 (5.7) | 30.3 (5.9) |
| Hypertension | 106,877 (79%) | 37,741 (78%) | 28,757 (77%) | 40,379 (81%) |
| Hemoglobin A1c, mean (SD) | 6.17 (1.10) | 6.19 (1.16) | 6.12 (1.03) | 6.18 (1.09) |
| Smoke |  |  |  |  |
| Current | 27,191 (20%) | 11,714 (24%) | 6,595 (18%) | 8,882 (18%) |
| Former | 86,223 (64%) | 28,881 (60%) | 24,149 (65%) | 33,193 (66%) |
| Never | 22,326 (16%) | 7,887 (16%) | 6,496 (17%) | 7,943 (16%) |
| Statin | 93,106 (69%) | 32,452 (67%) | 24,951 (67%) | 35,703 (71%) |
| Education |  |  |  |  |
| Less than high school | 4,869 (3.6%) | 1,855 (3.8%) | 1,089 (2.9%) | 1,925 (3.8%) |
| High school diploma/GED | 30,599 (23%) | 11,353 (23%) | 7,765 (21%) | 11,481 (23%) |
| Some college credit, but no degree | 41,679 (31%) | 15,960 (33%) | 11,013 (30%) | 14,706 (29%) |
| Associates degree | 17,328 (13%) | 6,695 (14%) | 4,810 (13%) | 5,823 (12%) |
| Bachelor's degree | 24,476 (18%) | 7,720 (16%) | 7,266 (20%) | 9,490 (19%) |
| Master's degree | 12,409 (9.1%) | 3,686 (7.6%) | 3,951 (11%) | 4,772 (9.5%) |
| Professional or Doctorate degree | 4,380 (3.2%) | 1,213 (2.5%) | 1,346 (3.6%) | 1,821 (3.6%) |
| Activity |  |  |  |  |
| Very good | 12,887 (9.5%) | 3,235 (6.7%) | 4,322 (12%) | 5,330 (11%) |
| Fairly good | 35,853 (26%) | 10,849 (22%) | 11,317 (30%) | 13,687 (27%) |
| Satisfactory | 47,433 (35%) | 17,044 (35%) | 13,275 (36%) | 17,114 (34%) |
| Fairly poor | 32,672 (24%) | 14,111 (29%) | 7,224 (19%) | 11,337 (23%) |
| Very poor | 6,895 (5.1%) | 3,243 (6.7%) | 1,102 (3.0%) | 2,550 (5.1%) |
| PAD | 24,283 (18%) | 8,816 (18%) | 6,121 (16%) | 9,346 (19%) |

### **Supplementary Table 7. Baseline characteristics of 452,028 participants by sleep duration in UK Biobank**

|  | **Sleep duration in hours per night** | | | | **Total** |
| --- | --- | --- | --- | --- | --- |
| **No. of individual and characteristics** | **<5** | **≥5 & <7** | **≥7 & <8** | **≥8** |  |
| No. of Individuals | 3802 | 104 566 | 177 596 | 166 064 | 452 028 |
| Age, mean±SD, years | 57.1±7.6 | 56.7±7.8 | 56.0±8.0 | 57.5±8.1 | 56.7±8.0 |
| Sleep duration, median±IQR, hours | 4±0 | 6±0 | 7±0 | 8±0 | 7±1 |
| Male, % | 42.5 | 46.7 | 47.2 | 43.4 | 45.7 |
| Body mass index, mean±SD, kg/m^2^ | 28.9±5.8 | 27.9±5.0 | 27.1±4.6 | 27.3±4.7 | 27.4±4.7 |
| With college/university degree, % | 15.0 | 28.2 | 36.7 | 30.5 | 32.3 |
| Hypertension, % | 39.4 | 30.3 | 25.2 | 28.9 | 27.9 |
| Hypercholesterolemia, % | 19.5 | 14.0 | 11.6 | 14.3 | 13.2 |
| Diabetes, % | 8.4 | 5.3 | 3.9 | 5.4 | 4.8 |
| Smoking status, % |  |  |  |  |  |
| Never smoker | 48.4 | 51.9 | 55.9 | 54.3 | 54.3 |
| Past smoker | 32.7 | 35.8 | 34.7 | 36.1 | 35.4 |
| Current smoker | 18.9 | 12.3 | 9.5 | 9.7 | 10.3 |
| Physical activity, % |  |  |  |  |  |
| Low | 18.2 | 16.0 | 14.8 | 14.7 | 15.1 |
| Moderate | 24.4 | 31.1 | 34.5 | 33.6 | 33.3 |
| High | 30.0 | 32.6 | 33.4 | 33.1 | 33.1 |
| Cardiometabolic healthy diet score, mean±SD | 17.3±3.4 | 17.6±3.4 | 17.8±3.4 | 17.6±3.4 | 17.7±3.4 |
| Daytime napping, % | 43.1 | 41.1 | 38.7 | 49.1 | 43.1 |
| Polygenic risk score, mean±SD | -0.3±0.2 | -0.3±0.2 | -0.3±0.2 | -0.3±0.2 | -0.3±0.2 |

IQR, interquartile range; SD, standard deviation.

Missing information was around 0.31% for body mass index, 0.93% for education, 0.84% for smoking status, 0.33% for diet, 18.5% for physical activity.

### **Supplementary Table 8. Association of genetic predisposition to sleep apnea and snoring with peripheral artery disease risk**

| **Exposure** | **SNPs** | **Method** | **OR** | **95% CI** | ***p*** |
| --- | --- | --- | --- | --- | --- |
| Sleep apnea | 34 | IVW-random effects | 1.05 | 0.78, 1.40 | 0.754 |
|  |  | Weighted median | 1.21 | 0.96, 1.53 | 0.113 |
|  |  | MR-Egger | 0.66 | 0.27, 1.59 | 0.358 |
|  |  | MR-PRESSO | 1.14 | 0.94, 1.39 | 0.201 |
|  |  | Cochrane’s Q = 130 (*p* <0.001); MR-Egger intercept = 0.012 (*p* = 0.282) | | | |
| Snoring | 23 | IVW-random effects | 1.03 | 0.52, 2.06 | 0.927 |
|  |  | Weighted median | 1.10 | 0.44, 2.76 | 0.844 |
|  |  | MR-Egger | 1.29 | 0.03, 51.89 | 0.895 |
|  |  | MR-PRESSO | NA | NA | NA |
|  |  | Cochrane’s Q = 29 (*p* = *0.158*); MR-Egger intercept = -0.002 (*p* = 0.907) | | | |

CI, confidence interval; IVW, inverse-variance weighted; SNPs, single nucleotide polymorphisms.

### **Supplementary Table 9. Heterogeneity, pleiotropy, and outliers in the Mendelian randomization analyses**

| **Trait** | **used SNPs** | **Cochrane's Q** | **Intercept** | ***p* for intercept** | **Outliers** |
| --- | --- | --- | --- | --- | --- |
| **Sleep → PAD** |  |  |  |  |  |
| Sleep duration | 73 | 183 | -0.006 | 0.438 | 2 |
| Short sleep | 27 | 37 | -0.005 | 0.549 | 2 |
| Long sleep | 7 | 29 | -0.063 | 0.044 | 3 |
| Insomnia | 207 | 353 | 0.004 | 0.372 | 3 |
| Daytime napping | 102 | 183 | 0.004 | 0.417 | 2 |
| **PAD → Sleep** |  |  |  |  |  |
| Sleep duration | 19 | 35 | -0.003 | 0.179 | 2 |
| Short sleep | 19 | 27 | 0.006 | 0.204 | 0 |
| Long sleep | 19 | 19 | -0.006 | 0.231 | 0 |
| Insomnia | 19 | 29 | 0.004 | 0.357 | 0 |
| Daytime napping | 19 | 55 | -0.001 | 0.646 | 2 |

SNPs, single nucleotide polymorphism.

### **Supplementary Table 10. Associations of peripheral artery disease with sleep-related traits in reverse Mendelian randomization analysis based on SNPs from the VA Million Veteran Program**

| **Outcome** | **MR method** | **Effect estimate** | | |  | **Test of pleiotropy** | |
| --- | --- | --- | --- | --- | --- | --- | --- |
|  |  | **OR/Beta** | **95% CI** | ***p*** |  | **Test** |  |
| Sleep duration | IVW-random effects | 0.23 | -0.97, 1.43 | 0.708 |  | Cochran Q value | 89 |
|  | Weighted median | 0.52 | -0.41, 1.45 | 0.271 |  | MR-Egger intercept (*p*) | 0.586 |
|  | MR-Egger | 0.99 | -1.97, 3.96 | 0.520 |  | Outliers | 3 |
|  | MR-PRESSO | 0.59 | -0.38, 1.56 | 0.254 |  | Distortion test (*p*) | 0.640 |
|  | IVW-random effects | 1.06 | 1.01, 1.11 | 0.015 |  | Cochran Q value | 29 |
| Short sleep | Weighted median | 1.04 | 0.99, 1.10 | 0.161 |  | MR-Egger intercept (*p*) | 0.354 |
|  | MR-Egger | 1.01 | 0.90, 1.13 | 0.897 |  | Outliers | 0 |
|  | MR-PRESSO | NA | NA | NA |  | Distortion test (*p*) | NA |
|  | IVW-random effects | 1.03 | 0.95, 1.12 | 0.491 |  | Cochran Q value | 59 |
| Long sleep | Weighted median | 1.05 | 0.98, 1.13 | 0.161 |  | MR-Egger intercept (*p*) | 0.272 |
|  | MR-Egger | 1.15 | 0.93, 1.42 | 0.204 |  | Outliers | 3 |
|  | MR-PRESSO | 1.02 | 0.96, 1.08 | 0.496 |  | Distortion test (*p*) | 0.397 |
| Insomnia | IVW-random effects | 1.02 | 0.99, 1.06 | 0.222 |  | Cochran Q value | 14 |
|  | Weighted median | 0.99 | 0.94, 1.05 | 0.807 |  | MR-Egger intercept (*p*) | 0.189 |
|  | MR-Egger | 0.96 | 0.88, 1.05 | 0.442 |  | Outliers | 0 |
|  | MR-PRESSO | NA | NA | NA |  | Distortion test (*p*) | NA |
|  | IVW-random effects | 1.00 | 0.98, 1.03 | 0.708 |  | Cochran Q value | 89 |
| Daytime napping | Weighted median | 1.01 | 0.99, 1.03 | 0.264 |  | MR-Egger intercept (*p*) | 0.586 |
|  | MR-Egger | 1.02 | 0.96, 1.09 | 0.520 |  | Outliers | 3 |
|  | MR-PRESSO | 1.01 | 0.99, 1.03 | 0.254 |  | Distortion test (*p*) | 0.637 |

CI indicates confidence interval; NA, not available; OR, odds ratio; SNPs, single-nucleotide polymorphisms; UKBB, UK Biobank.

### **Supplementary Figure 1. Study population of the SIMPLER study**

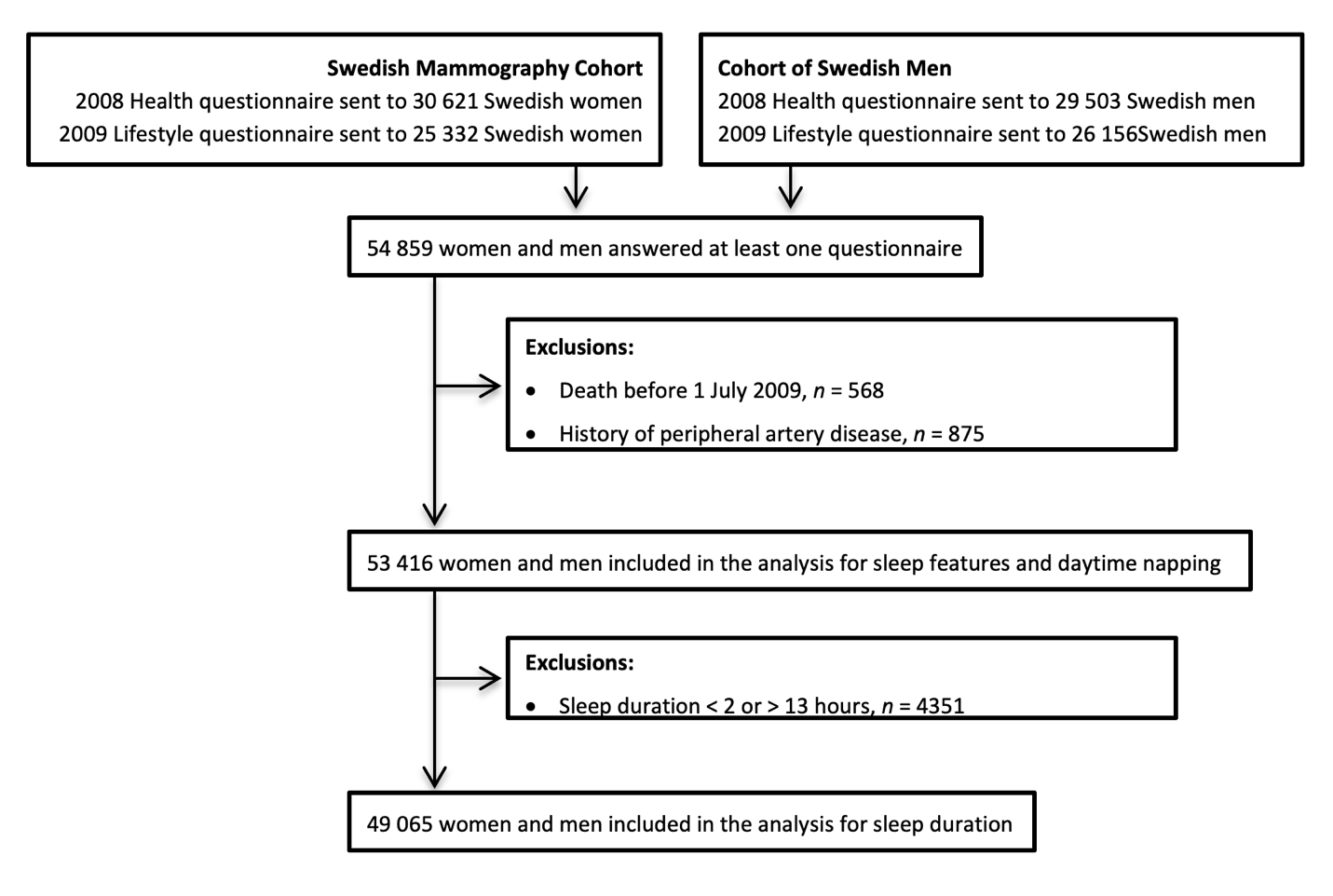
